# ArcticAI: A Deep Learning Platform for Rapid and Accurate Histological Assessment of Intraoperative Tumor Margins

**DOI:** 10.1101/2022.05.06.22274781

**Authors:** Joshua Levy, Matthew Davis, Rachael Chacko, Michael Davis, Lucy Fu, Tarushii Goel, Akash Pamal, Irfan Nafi, Abhinav Angirekula, Brock Christensen, Matthew Hayden, Louis Vaickus, Matthew LeBoeuf

## Abstract

Successful treatment of solid cancers relies on complete surgical excision of the tumor either for definitive treatment or before adjuvant therapy. Radial sectioning of the resected tumor and surrounding tissue is the most common form of intra-operative and post-operative margin assessment. However, this technique samples only a tiny fraction of the available tissue and therefore may result in incomplete excision of the tumor, increasing the risk of recurrence and distant metastasis and decreasing survival. Repeat procedures, chemotherapy, and other resulting treatments pose significant morbidity, mortality, and fiscal costs for our healthcare system. Mohs Micrographic Surgery (MMS) is used for the removal of basal cell and squamous cell carcinoma utilizing frozen sections for real-time margin assessment while assessing 100% of the peripheral and deep margins, resulting in a recurrence rate of less than one percent. Real-time assessment in many tumor types is constrained by tissue size and complexity and the time to process tissue and evaluate slides while a patient is under general anesthesia. In this study, we developed an artificial intelligence (AI) platform, *ArcticAI,* which augments the surgical workflow to improve efficiency by reducing rate-limiting steps in tissue preprocessing and histological assessment through automated mapping and orientation of tumor to the surgical specimen. Using basal cell carcinoma (BCC) as a model system, the results demonstrate that ArcticAI can provide effective grossing recommendations, accurately identify tumor on histological sections, map tumor back onto the surgical resection map, and automate pathology report generation resulting in seamless communication between the surgical pathology laboratory and surgeon. AI-augmented-surgical excision workflows may make real-time margin assessment for the excision of more complex and challenging tumor types more accessible, leading to more streamlined and accurate tumor removal while increasing healthcare delivery efficiency.

## Introduction

Complete surgical resection is first line treatment for many solid tumors, which typically requires excision of the clinically evident tumor and the rim of surrounding normal tissue followed by closure with subsequent post-operative histologic analysis of tissue margins (POMA). In the pathology laboratory, the specimen is most commonly grossed in a breadloafed or radial fashion, embedded, sectioned, stained, and read by the pathologist. While POMA of breadloafed sections is the current standard, removal and histologic analysis of the margin in this manner has three major pitfalls: 1) post-operative identification of positive margins (tumor identified at the tissue edge), necessitating a repeat procedure, 2) false negative or “missed” margins, where the tumor is present at a portion of the margin not evaluated due to sampling error, and 3) excessive tissue is removed to limit the possibility of pitfalls 1 and 2 as above, which can result in the removal of critical structures. Standard excisions and POMA for the treatment of skin cancer reveals a combined positive margin or tumor recurrence rate of at least 20% ^1–4^. Either of these outcomes (i.e., post-operative positive margins or false negative margins) requires additional surgery, radiation, chemotherapy, or some combination thereof, resulting in patient morbidity, mortality, and a significant cost to our healthcare system ^5–10^. These pitfalls have been addressed in some settings through the use of intraoperative frozen sections or analyzing a larger percentage of tissue margins, which, in comparison to standard excisions and POMA, can reduce positive margin or recurrence rates to less than 1-2% in certain surgical subspecialties ^3,11–13^.

Successful intraoperative treatment of solid tumors requires the combined efforts of multiple highly trained individuals. Tumor removal is performed by the surgeon; cryofreezing and sectioning of the tissue and then staining by the histotechnologist; and histologic analysis by the pathologist. In the current surgical workflow, the surgeon, histotechnologist, and pathologist are often separated by time and space. For example, communication of histological findings between pathologist and surgeon may occur over the phone. This separation presents an obstacle to evaluating intraoperative frozen sections (**Figure 1**). Prior studies have shown that breadloaf grossing of tissue sections results in analysis of approximately 1-2% of the margin ^6^. Increasing the percentage of tissue margins analyzed requires either: 1) more tissue blocks and sections, or 2) an alternative grossing method. These approaches require more time and/or expertise on the part of both the histotechnician and pathologist.

**Figure 1:**
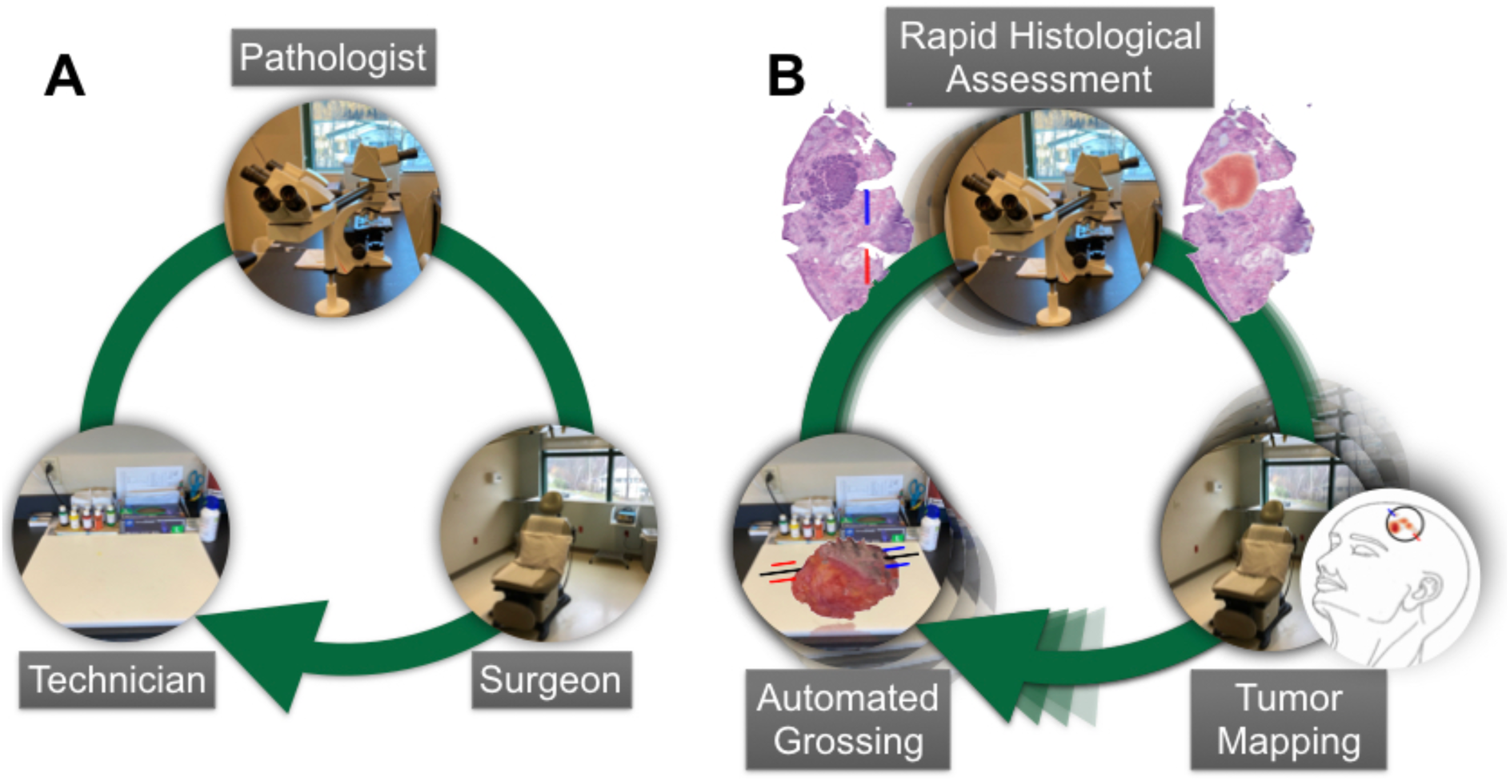
Intraoperative surgical excision setting and potential use cases for integrating artificial intelligence: **A)** Surgeon removes tumor in the operating room, tissue is prepared in the gross room, margins are assessed by pathologist in slide room for margin assessment and findings are mapped back to orientation of surgical site. **B)** 3D modeling for automated tissue grossing, computer vision and graph neural networks for margin assessment, and morphing techniques orient histological findings to a surgical tumor map to inform surgeon where to cut additional tumor

Mohs Micrographic Surgery (MMS) is used for the treatment of skin cancers of the head, neck, and special sites ^14,15^. Tumor removal is performed under local anesthesia with real-time margin assessment using frozen tissue sections that are cut by a histotechnician in an on-site laboratory. Tissue is grossed in a manner that allows the peripheral and deep margin to sit in the same plane, allowing analysis of 100% of tissue margins. The Mohs Micrographic Surgeon performs tumor removal, histologic analysis, and the creation of a surgical tumor map to inform additional tumor removal if necessary. As compared to standard excisions and POMA (≥20% recurrence, as aforementioned), MMS results in a significantly lower tumor recurrence rate (less than 1-2%, as aforementioned) while minimizing the size of the surgical defect and sparing normal surrounding tissue. The advantages conferred by MMS are largely possible due to the size and location of the tumors being excised. These characteristics facilitate the use of local anesthesia and allow the entire margin to be efficiently processed and analyzed. There are numerous obstacles to the application of real-time 100% margin analysis in other surgical practices, including: 1) time under general anesthesia, 2) tissue specimens of prohibitive size and complexity, 3) availability of expert pathologists, and 4) clear mapping of histological findings back to the resection site. Additionally, suboptimal preparation of frozen sections can impact the location of positive margins and is often cited as precluding real-time margin assessment in higher risk settings. Highly trained histotechnicians are required to create quality tissue sections but are in short supply ^16^. Thus, investing in methods that can improve the speed of specimen preparation, ensure high-quality tissue sections, and promote rapid and accurate histological assessment of tissue margins are of paramount importance.

Emerging artificial intelligence (AI) technologies have demonstrated the capacity to model complex medical processes and may soon fundamentally transform healthcare delivery through incorporation of non-autonomous diagnostic decision making. These technological advancements have been propelled through the advent of artificial neural networks (ANN) including deep learning methodologies ^17^. ANN are inspired by central nervous system processes and represent data input to the algorithm through a collection of nodes, where, given the appropriate activation energy, the signal from these nodes may be passed or shared to a hidden set of nodes organized into multiple processing layers which represent an object through multiple layers of abstraction. For instance, ANN have been widely applied to tasks in digital pathology ^18–26^, from simulating application of chemical staining reagents ^27–34^, to predicting prognostic molecular information from digitized representations of tissue slides (Whole Slide Images; WSI), and predicting the origin of tumors with unknown primary site ^35^. Recent ANN methods have been proposed for margin assessment across multiple surgical subspecialties though have only focused on identifying tumor ^36–39^ while ignoring other issues that are critical to MMS, including: 1) assessing 100% tissue margins intraoperatively, 2) tissue preparation, 3) tissue section quality, 4) mapping findings to surgical tumor site, and 5) operational efficiency.

We have designed and developed a non-autonomous artificial intelligence driven platform (*ArcticAI)* that can expedite tissue preparation, histological inspection, and tumor mapping to improve solid tumor removal **(Figure 2)** using MMS for removal of basal cell carcinoma as a model system. *ArcticAI* places the surgeon, histotechnician, and pathologist in the same virtual space to: 1) reduce the amount of time a histotechnician takes to process tissue and generate pathology reports through 3D modeling techniques and smart grossing recommendations (e.g., reporting of tissue size and where to ink), 2) improve the efficiency of pathologic analysis through a collection of sophisticated graph neural networks to map tumor and artifacts on whole slide images (WSI) acquired from serial tissue sections, and 3) automatically generate a descriptive and visual pathology report easily interpreted by the surgeon either in real-time or post-operatively.

**Figure 2:**
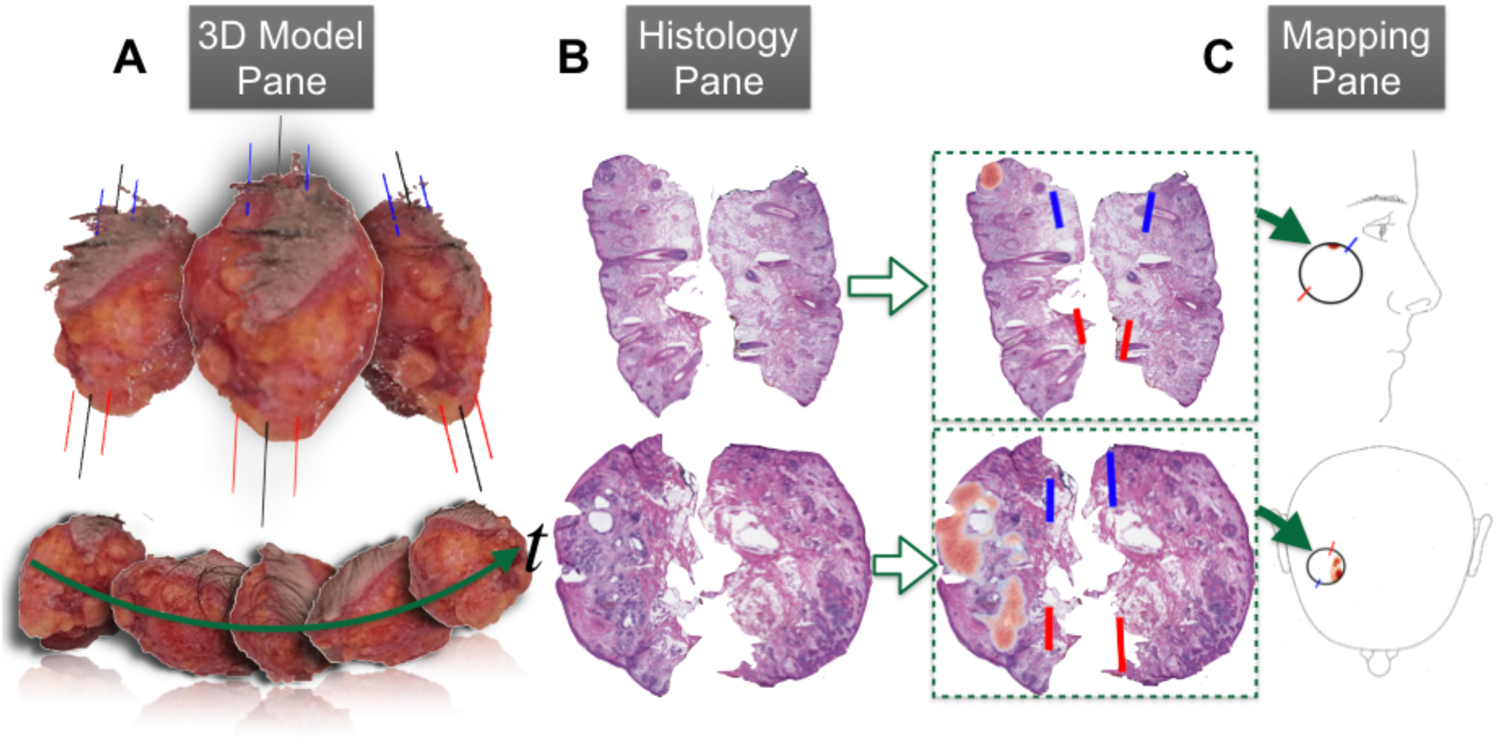
Workflow overview: **A)** Gross tissue measurements and inking recommendations are made using the *3D Model Pane*, which reconstructs 3D models of tissue from video of tissue rotating around a turntable set up, **B)** Rapid margin assessment is accomplished through the *Histology Pane*, which localizes holes/tears (completeness), tumor, and calculates spatial statistics on ink for orientation (blue is 12 o’clock, red is 6 o’clock), and **C)** *Mapping Pane* maps margin assessment results to surgical specimen through morphing to user defined surgical tumor map and by leveraging the orientation calculated in the *Histology Pane* to reorient the results to a format understandable by the surgeon

## Results

### Data Collection and Study Population

After Institutional Review Board approval, we assessed specimens from 194 patients undergoing tumor excision in the Mohs Micrographic Surgery (MMS) setting for the treatment basal cell carcinoma (BCC). Tissue from 16 patients (17 specimens) were used for tissue grossing algorithms, while tissue from the remaining 178 patients were used for histological assessment and tumor mapping algorithms. All specimens first underwent accessioning and gross measurement. For the tissue grossing algorithm, the gross specimen was placed on a turntable and imaged using low resolution video capture. The remaining cases underwent grossing, inking, processing, cryoembedding (frozen section), sectioning, and staining with hematoxylin and eosin (H&E). From these 178 cases, 351 slides corresponding to 1,065 serial sections and 1,537 tissue pieces were scanned at 20X resolution using the Leica Aperio AT2 scanner and stored as Whole Slide Images (WSI) in either SVS or TIFF file format. A total of 3,754,730 image patches (256×256 pixels) were extracted from the WSI for further analysis. Using the ASAP annotation software (https://computationalpathologygroup.github.io/ASAP/, v1.9), all cases were annotated for tumor (BCC), benign structures, inflammatory aggregates, holes/tears, ink color and location, and major compartments (epithelium, dermis, fat, etc.). The data was then divided into training/validation sets (65% of cases) responsible for algorithm training and finetuning and a held-out test set (35% of cases) (**Table 1**). Follicles and individual nuclei were annotated in a subset of the training/validation slides as annotation of these smaller structures on all training/validation slides was intractable. BCC subtypes seen in clinical practice were reflected in the training/validation and test sets.

### Results Overview

In the following subsections, the impact of an AI-augmented digital assessment on the surgical workflow will be demonstrated through description of expected display outputs and results from: 1) tumor removal and specimen preparation, 2) histological assessment, and 3) tumor mapping. Finally, execution time is recorded for rapid and parallel histological assessment.

Following tumor removal, a specimen is sent to the pathology laboratory for accessioning, grossing, inking, sectioning, and staining. Tissue grossing and inking decisions are made by the histotechnician depending on the size and shape of the tissue specimen. These decisions are not standardized and require a high level of training and rigorous documentation. To determine if tissue characteristics could be autogenerated, seventeen surgical specimens were collected, and the superior pole was delineated by either tissue ink or placement of a suture. Reconstructions combined multiple views of the tissue through captured smartphone videos for one revolution around a turn table setup (**Figure 3A**). These tissue images were cropped using a tissue detection algorithm into serial images and photogrammetry techniques were applied to reconstruct a 3D model of the excised tissue (**Figure 3B; Supplementary File 1**).

**Figure 3:**
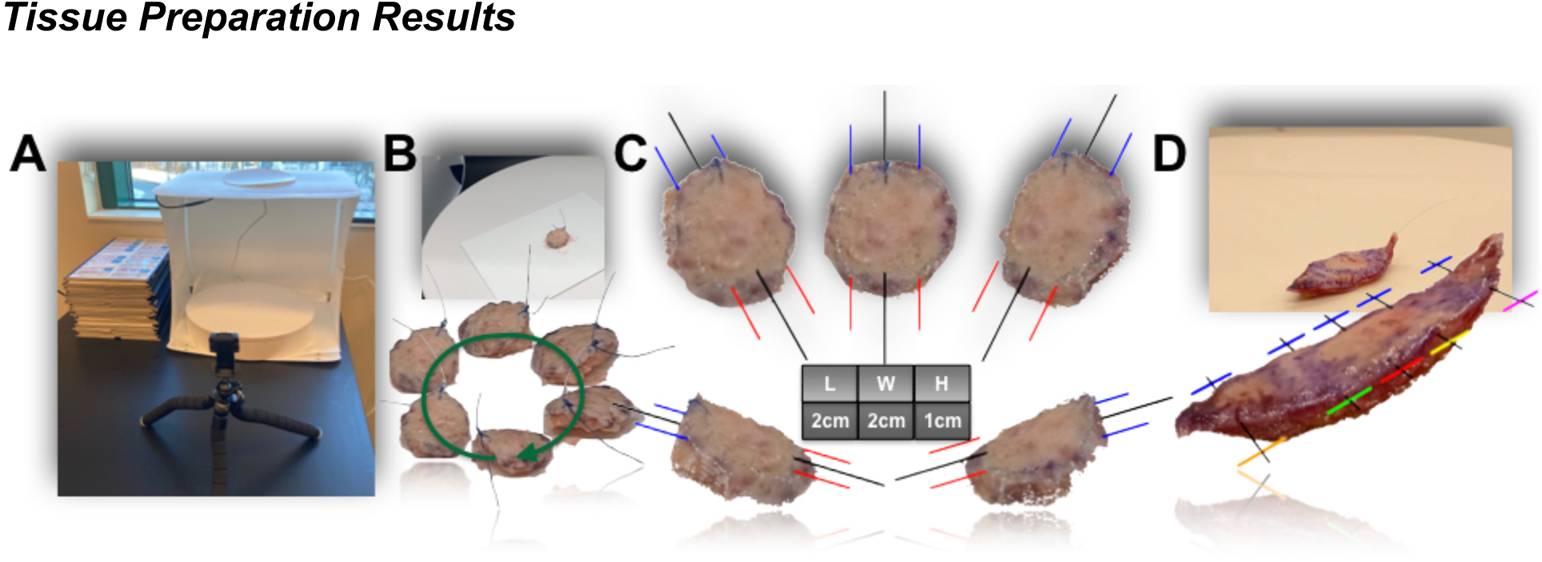
Tissue Grossing Measurement and Recommendations via the *3D Model Pane*: **A)** Image of turntable setup, where phone camera is placed on mount to record gross specimen revolve around table; **B)** still frame from phone video of rotating tissue specimen; also depicted on the bottom are automated segmentations of the tissue and suture using the *3D Model* preprocessing subroutine, images were selected from a representative set of still frames to demonstrate multiple object viewpoints; **C)** these viewpoints are integrated together for 3D reconstruction of the gross specimen, pictured here are screenshots while using the interactive *3D Model Pane* to rotate the specimen to various orientations; inking recommendations are deposited via the addition of red/blue and black lines, which denote inking of 12 o’clock (blue) near the suture (which has been removed) and 6 o’clock (red) after bisecting the tissue (black);; length, width and height measurements are automatically reported on the scale of centimeters while operating the display; **D)** tissue grossing and ink recommendations for radial sectioning of a wide local excision specimen

#### Tissue Size Measurements

Subsequently, additional filtration techniques were applied to the model (i.e., suture was automatically segmented and removed from the 3D model), and ellipse shape detectors ^40^ were used to measure the width of the turn table to normalize the dimensions of the 3D model to actual tissue proportions (**Figure 3C**). Automated tissue measurements of the reconstructed tissue varied by 0.25cm on average as compared to manual measurements for each tissue dimension **(Tables 2-3)**.

#### Inking Recommendations

Taking into account: 1) autogenerated tissue size, 2) preferred grossing approach (MMS versus radial) as dictated by surgeon/pathologist, and 3) size of a glass slide, automated grossing and inking measurements/recommendations were generated to maximize the amount of margin per tissue block/slide. This resulted in lines being placed through the 3D model recapitulating expert domain knowledge, using suture/ink locations for guidance. For MMS specimens, the algorithm placed 3D black lines, which identified the location of grossing cuts, and blue and red lines, denoting 12 and 6 o’clock ink placement, respectively (**Figure 3C**). **Figure 3D** demonstrates tissue grossing and ink recommendations for radial sectioning of a wide local excision specimen. Grossing cuts are identified by black lines through the body and two tips of the specimen based on tissue size. Inking recommendations include unique color combinations for each tissue piece allowing multiple pieces to be put into a single cassette, resulting in fewer tissue blocks for the histotechnician to section.

### Histological Assessment Results

#### Tissue Completeness Assessment

Effective histologic analysis of tumor margins relies on high-quality tissue sections that are devoid of holes or tears. In the absence of a complete tissue section, it is not possible to definitively declare a margin free of tumor. To address this, a ‘Tissue Completeness’ algorithm was developed using a combination of convolutional and graph convolutional neural networks (CNN-GNN) ^41^. The algorithm was trained and validated on 381 annotated tissue sections using PyTorch to segment holes/tears (tissue artifacts) in the tissue ^41,42^. The tissue completeness algorithm was trained to delineate between the following macro-architectural features: 1) holes/tears, 2) epidermis, 3) dermis, and 4) subcutaneous fat. The algorithm successfully identified tissue defects in our test set with an AUC of 0.84 (**Table 3**). An example output of the ‘Completeness’ algorithm is shown in **Figure 4A-B**, where sporadically placed holes/tears are highlighted by the algorithm, while regions of fat or significant gaps introduced by hair follicles or less structured dermis are ignored. A few instances where holes were missed or overcalled are shown in **Supplementary Figure 1**.

**Figure 4:**
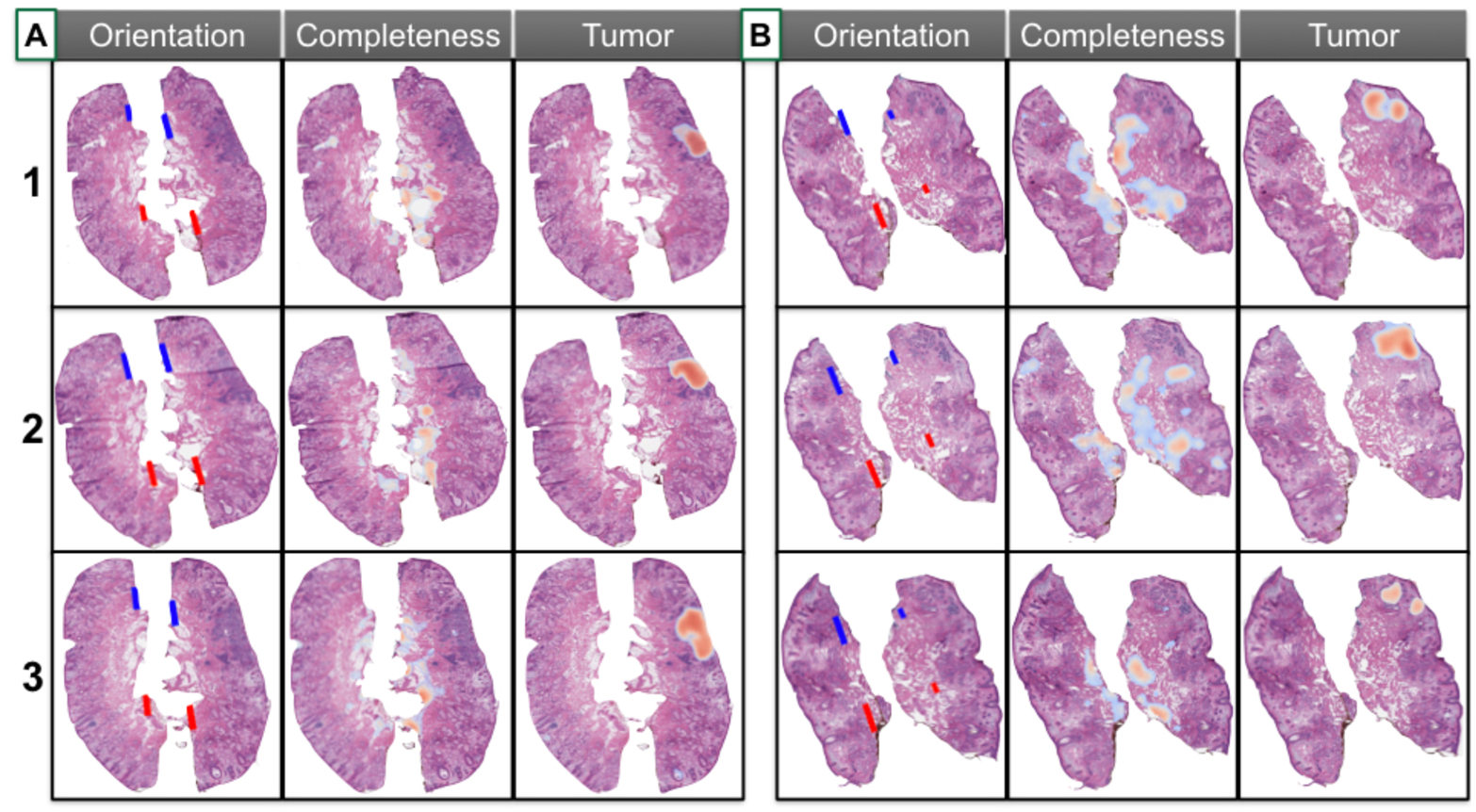
Margin assessment via the *Histology Pane* for two test cases, A and B for three serial sections (1,2, and 3). Results are plotted on top of each WSI for assessments of orientation, tissue completeness, and tumor localization. Screenshots of WSI from the *Histology Pane*. For tissue orientation, blue and red lines are drawn over center of mass positions to define 12 o’clock and 6 o’clock respectively for each tissue section. High resolution completeness and tumor results represented by thresholded heatmaps (where patches removed from display if failing to surpass probability threshold), where red indicates whether part of tissue is incomplete or positive margins, and blue indicates lower yet non-negligible probability of incompleteness/tumor.

#### Tumor Localization

To evaluate for the presence or absence of tumor, a CNN-GNN was trained and validated on 1,065 tissue sections containing a variety of basal cell carcinoma histologic subtypes reflective of clinical practice **(Table 1).** Annotation subgroups included: tumor, benign skin structures (hair follicles), and cell populations that may be confused with or a harbinger of tumor (inflammation). Using a small subset of WSIs, an image detection model was trained to identify and remove hair follicles from tumor-predicted regions that were conflated with follicles (**Table 2, Supplementary Figure 2, Supplementary Table 1**). Inflammation was explicitly modeled to avoid calling tumor in pockets of inflammation. On a test set on 121 held out slides, the CNN-GNN obtained an AUC of 0.97 for the task of tumor localization across sections (**Table 3**). Example displays of the tumor detection output across serial sections of two test set cases is shown in **Figure 4A-B**. We have included example displays of hair follicle and inflammation-predicted regions on held-out slides in the supplementary materials (**Supplementary Figures 2-3**), which further demonstrate how exclusion of these regions can inform tumor localization.

#### Nuclei Detection and Classification

To rule out rare tumor cells in regions predicted to be inflammatory aggregates, a cell detection neural network (Detectron2) was trained to provide high-resolution tumor maps designating precisely which cells correspond to the BCC annotation subgroup ^43,44^. Annotation subgroups to train the cell detection algorithm included: ‘BCC’, ‘hair follicle’, ‘inflammatory’, ‘fibroblast’, and ‘epidermal keratinocyte’. The model was trained and validated on 32,763 cell annotations (polygonal contours). Initial cell assignments were further refined by training a GNN on cell graphs (node is cell and spatial proximity to neighboring cells form edges). Results demonstrate the ability to accurately localize cells (Dice=0.86) in a small internal test set, while predicting with high accuracy the corresponding cell type (F1-Score=0.86) (**Supplementary Table 1; Supplementary Figure 4**).

#### Tissue Orientation

Histologic ink location is critical to tissue orientation and subsequent tumor mapping. In this study, MMS specimens were inked blue (12 o’clock), and red (6 o’clock) which was reflected in the surgeon’s hand-drawn diagrams. To automate ink detection, tissue edges were segmented using a Sobel filter with morphological dilation and opening operations. Then, a sensitivity analysis over thresholds in Hue, Saturation, Value (HSV) color space yielded optimal color thresholds to detect inks, which were paired with a connected component analysis to identify contiguous regions and remove spurious applications of ink within the tissue edges (i.e., where ink is erroneously applied / seeps) ^45^. Subsequently, a line was plotted between detected blue and red ink on tissue sections and stored for use later to calculate the relative orientation to lines drawn either with surgeon annotated inks on histological slides as comparison or to inks drawn in the surgical tumor map for mapping histological results back to the specimen (**Figures 4–5**). On a subset of held out test slides, the relative angular difference between red and blue ink line was measured and compared to the relative angular difference to the blue-red ink line calculated from the surgeon annotation of inks on the WSI (center of mass calculations for detected/annotated inks). Findings indicate that 95% of tissue sections were oriented correctly (i.e., less than 45° difference between annotated/predicted lines) with an average relative angular difference of 4% (**Table 3, Supplementary Figure 5A,C**). Sections without correct orientation demonstrated relative lack or spurious applications of ink, highlighting the importance of proper tissue inking (**Supplementary Figure 5B**)^46^.

**Figure 5:**
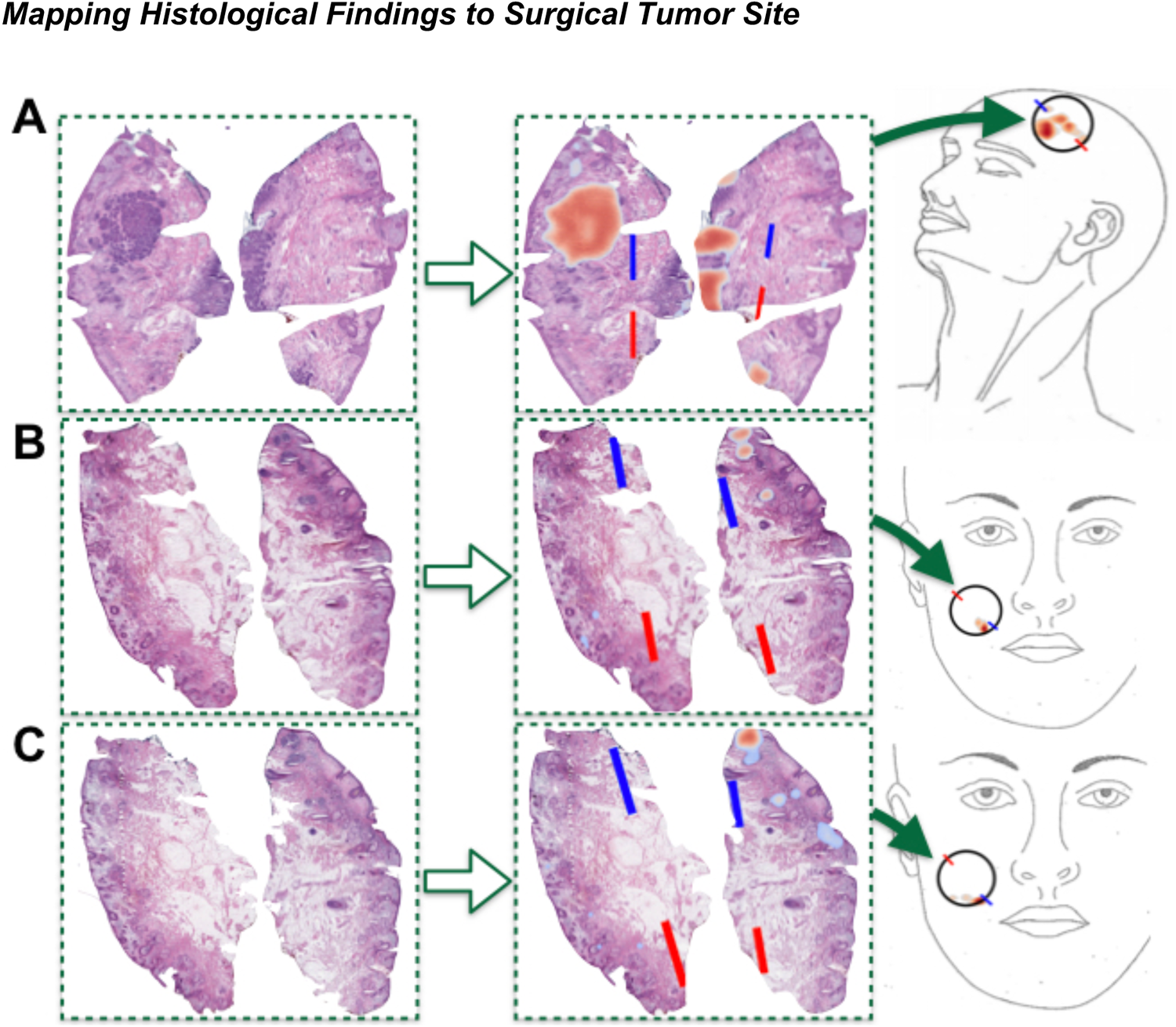
Mapping margin assessment results to surgical tumor map via *Mapping Pane*: **A-C)** Representation of workflow using three separate sections, the first **(A)** from one case, and the second two **(B-C)** are serial sections in another case; first, margins are assessed via *Histology Pane* and tissue orientation and tumor localization results are plotted over the WSI; then, results are mapped to surgical tumor diagram selected by the user (**A** features top of scalp, while case **B-C** are of front of face), where circle is drawn by user to represent surgical site anatomic location and an arbitrary orientation is defined via user drawing of blue/red lines. Note how tumor results are morphed and rotated to match circle interior and orientation in surgical tumor maps, where density map in surgical tumor map represents tumor at user defined threshold. For **B-C**, note how tumor is automatically rotated close to 180 degrees to preserve orientation of margin on surgical map

#### Tumor Mapping

Accurate tumor mapping is critical to inform additional tumor removal if needed. Tumor mapping relies on anatomic identification of surgical site, accurate tissue size measurement, tissue orientation, and tumor identification. To build a tumor map, a template is selected by the surgeon based on the anatomical surgical site with blue and red lines used to indicate 12 o’clock and 6 o’clock, respectively (**Supplementary Figure 6**). The tissue sections are then fit to the tumor map through an algorithm that morphs the WSI histologic tissue sections into the shape of the map using an optimal transport method and rotating the map such that orientation (blue-red ink line) predicted using the inks aligns with the blue and red surgical map lines (**Figure 5A-C** illustrates how histological results can mapped to any arbitrary orientation**, Supplementary File 2** demonstrates concordance between hand-drawn and digital tumor maps) ^47–49^. Both tumor and tissue completeness are mapped and can be viewed separately or overlapping. For each tumor removal stage, information across multiple serial sections is integrated together or can be viewed separately after registering the histological findings to the surgical tumor map. To determine the accuracy of automated tumor mapping, 28 test set cases (selected at random from cases with known positive margins) were used to compare platform generated tumor maps to the surgeon hand drawn maps. Maps corresponding to first stage removals were assessed as most second stage removals were clear at the margin. This showed 99.2% (95% CI: 91.5%-99.9%) correspondence between the surgeon and algorithm generated maps, respectively (**Supplementary Figure 7, Table 2**).

### Margin Assessment Speed

Local anesthesia is used during MMS. For broad applicability of this approach for tumors in patients where general anesthesia is required, the platform must perform with efficiency and speed. The platform workflow ^50^ includes an initial preprocessing step to separate tissue sections and then parallel execution of all subroutines (**Supplementary Figure 8A**) across separated sections within the WSI (**Table 4**). Overall, margin assessment using the *ArcticAI* platform across the entire test set (n=41 cases, 121 slides) had an average execution time of 72 seconds per slide and 78 (95% CI: [66 - 88]) seconds per case (i.e., many slides/sections per case), consisting of 48 seconds for preprocessing and 24 seconds for parallel performance of image stitching, CNN-GNN analysis, and tissue orientation (**Supplementary Figure 8B**). Execution of the platform in series would take five to seven times longer than parallelized, 494 (95% CI: [367 - 553]) seconds per case.

## Discussion

Tumor excision with real-time intraoperative 100% margin assessment results in low recurrence rate and efficient delivery of surgical care in MMS. Real-time margin analysis or increased percentage of margin analysis has the potential to decrease true positive or false negative margins across surgical oncology procedures. However, broad applicability of real-time total margin analysis is relatively limited outside the MMS setting. There are several logistical constraints that contribute to this including separation of multiple experts in time and space, inefficient manual laboratory processes, and labor-intensive pathologic analysis of histologic specimens. In this study a rapid tissue margin assessment tool was designed and tested to address the rate limiting steps in the current surgical tumor removal workflow.

The *ArcticAI* platform performs automated tissue measurements aimed at improving laboratory workflow through efficient grossing and inking recommendations. These recommendations aim at maximizing the amount of tissue per block while decreasing the number of tissue blocks to be cut by the histotechnician. Additionally, unique predetermined ink combinations allow the tissue sections to be reconstructed and mapped to the 3D tissue model. These features have the potential to standardize grossing and inking in the surgical pathology laboratory thereby decreasing the time of processing and required level of expertise. Histotechnicians are highly trained and currently in high demand in our healthcare system. Increasing the efficiency of the histotechnician as well as decreasing the training required to expertly process and section a tissue excision specimen are two solutions to address the demand for more histotechnicians^51^. In addition to the importance of the ‘Completeness’ algorithm for tumor margin analysis, *ArcticAI* can also be used as a training tool to assess the competency of histotechnicians either in training or as part of an annual review of performance. Taken together, the platform provides significant support in training, standardization, and workflow efficiency for histotechnicians.

The current study resulted in an AUC of 0.97, which is anticipated to increase with subsequent and expanded training sets. Identification of specific histologic BCC subtypes or normal structures that present particular challenges for the CNN-GNN will help to identify surgical cases that will be most impactful for further improvement of the algorithm. Interestingly, this study elucidates the importance of the relationship between tumor and additional cell populations including surrounding inflammation as an indicator of the presence of tumor. Further delineation of the tumor niche may provide additional important information to delineate tumor from surrounding benign tissue or structures such as hair follicles which can have similar structures and nuclear morphologies to BCC. The creation of CNN-GNN for the margin analysis of other solid tumors will require the identification of both varied histologic tumor types and tissue specific cell populations or tissue structures that aid in tumor identification.

MMS is possible because the surgeon removes the tumor and reads the slides in a laboratory in close proximity to the operating room. In other surgical settings, performing frozen section margin analysis requires an onsite pathologist to read the slides and relay the results back to the surgeon in the operating room. In these settings the laboratory and operating room are separated by time and space. This prevents the use of frozen sections in many healthcare settings, particularly smaller rural hospitals, where caseload or demand may not support the presence of an on-demand highly trained expert pathologist. By creating an algorithm that enables rapid and accurate identification of tumor combined with a virtual platform allowing for remote whole slide imaging and result viewing, the *ArcticAI* platform obviates the pathologist being in physical proximity to the operating suite. This allows a pathologist with expertise in one particular tumor type or organ system to maintain a high case load while providing highly specialized pathologic care to healthcare settings that might not otherwise have such access. Integration of the ‘Completeness’ algorithm will identify low quality or incomplete sections prior to pathologic analysis and allow the histotechnician to create additional sections as needed, prior to final pathologist review. This will minimize recuts and allow rapid sign out and reporting. Integration of the vast amount of data present in a pathology report through automation will decrease the amount of work on the back end and also provide both written and visual outputs that can be used either in real-time or post-operatively. Excessive charting and documentation result in pathologist burnout, limiting the amount of manual documentation will both increase productivity and decrease administrative burden ^52–54^.

For skin cancer of the head and neck, which is more challenging to assess than the model system featured in this work (BCC) due to increased tissue size and complexity, decreased positive margin and recurrence rates have been shown with real-time complete margin analysis ^55,55–62^. As many healthcare systems are functioning with decreased staffing and disruption of supply chains, delivery of efficient surgical care is critical for patient access and maximizing hospital resources. Limiting positive margins, tumor recurrence, or the need for adjuvant treatments will decrease the burden on the surgical and medical system ^63–65^. Access to remote pathologists using AI augmented platforms will allow both hospitals that may otherwise not be able to offer a surgical service to do so and increase the productivity of remote pathologists. Providing histotechnicians with a platform that automates their tedious tasks and makes grossing/inking recommendations that are reflected in the pathology report will allow them to focus their time and energy on embedding and sectioning the tissue, thereby increasing the number of specimens processed. Taken together, the use of technology in the delivery of surgical care will not just provide better outcomes for the patients, but also improve the efficiency of surgical care delivery in an unprecedented time of resource shortages (e.g., access to care).

Limitations of this study include that it was performed at a single site. Whole slide images in the training, validation, and test set were generated in a single laboratory with a standardized sectioning and staining protocol. Therefore, the next steps will include creation of an external test set from outside MMS units. Finalized tumor detection and completeness algorithms will likely require an input of whole slide images from multiple laboratories. Alternatively, sites aiming to use the platform could adopt a standardized workflow including reagents and process similar to those used to generate the tissue sections incorporated into the algorithms in this study. Obstacle to the usage of the platform include the availability whole slide scanners, as these are currently costly, and have large file uploads requiring a robust computing infrastructure or workstation capable of handling high throughput assessment. With time, the cost of scanners will decrease, and computing power will increase using graphics processing units (GPUs). Timely tissue processing and analysis is critical to seamless integration of the platform into the surgical workflow. The timing featured in this study considers parallel execution of workflow elements in optimal computing infrastructure. However, many high-performance computing environments are bottlenecked by the time it takes to submit and start simultaneous compute jobs as well as communication bottlenecks which may be workflow specific. In future studies, all aspects of the surgical workflow and platform including: 1) tissue transport and processing 2) slide scanning 3) image upload and processing of the ArcticAI platform and 4) pathologist review, via a simulated clinical trial will be timed to provide practical time estimates.

## Conclusion

Complete surgical removal of solid tumors remains most patients’ best chance at achieving a cure. In this study, MMS removal of BCC is used as a model system to highlight the integration of artificial intelligence and machine learning into the surgical workflow to address critical bottlenecks that might otherwise prevent real-time and/or complete tumor margin analysis. This model has the ability to improve surgical care delivery through technology driven standardization and automation as one approach to solve the significant labor and resource shortages and mismatches in the current system. This can be accomplished by: 1) improving the efficiency of the individuals and processes within the system and 2) increasing the number of individuals capable of performing a critical task. Nonetheless, adopting a digital aid requires stakeholder buy-in and a readiness for changing established practices, which carries significant barriers for entry. Dissemination and implementation of such technologies requires educational alignment and qualitative assessment of stakeholder interests and values. In order for such technology to be adopted, it will need to demonstrate significant improvement in efficiency over traditional methods while meeting the needs of surgeons, pathologists, and histotechnicians.

## Methods

### Technology Overview

*ArcticAI* is an AI-based software platform for the rapid assessment of tumor margins. The functions of *ArcticAI* are encapsulated in several modules including:

*Tissue grossing via the 3D Model Pane* (**Figure 2A**): When tissue arrives at the pathology laboratory, it undergoes accessioning, description, measurement, grossing, inking, processing, embedding, sectioning, and staining prior to pathologic analysis. To expedite this process, we have prototyped a mobile application that takes multiple images / video of the tissue and synthesizes them to form a 3D model of the tissue. This allows the system to: 1) determine tissue size (e.g., length, width, height) and orientation automatically and add this data to the pathology report, 2) create optimal grossing guides for the histotechnician, and 3) create optimum inking diagrams for the specimen (e.g., blue ink indicates piece is at 12 o’clock; ink used to establish a “coordinate system” for tissue “map”).

*Histological Assessment via the Histology Pane* (**Figure 2B**): Following tissue processing, slides are scanned to generate high resolution WSIs which are uploaded into the ArcticAI platform where they are assessed for 1) tissue orientation by detecting inking patterns, 2) tissue quality assessment (e.g. holes and tears from processing and sectioning), and 3) presence or absence of tumor, where 4) tumor confounders (e.g., identification of hair follicles) and 5) nuclei are classified to provide further clarification of histological findings (e.g., residual tumor within large pockets of inflammation).

*Mapping of Results to Surgical Specimen via the Mapping Pane* (**Figure 2C**) Outputs of data from the aforementioned algorithms, notably tissue inking/orientation and the presence or absence of tumor, are used to automatically transpose tumor predictions onto hand-drawn surgical maps. Automated mapping has the advantage of providing the precise location of remaining tumor to inform the surgeon if and where additional tumor needs to be removed. A pathology text report with information on tissue preprocessing is automatically generated and piped to the patient’s electronic health record. This information is communicated back to the surgeon, and tumor mapping results (graphics which resemble surgeon drawn tumor maps) are exported to the EHR system to update the automatically generated pathology report.

*Workflow automation:* Intraoperative resection with 100% margin analysis typically involves the inspection of 6-10 serial tissue sections and can take upwards of 30 minutes per patient under general/local anesthesia. *ArcticAI* was optimized to reduce histological inspection and tumor mapping time using a sophisticated workflow engine that can be executed in both high performance computing environments and local workstations using Toil and Singularity. The pipeline is additionally capable of processing multiple tissue sections across multiple whole slide images in parallel.

*Web Application:* Histotechnicians, pathologists, and surgeons can interact with the results in real-time as an interactive/exportable pathology report through a dynamic web application which contains the following panes: 1) Case upload and execution (*Selection Pane*), 2) 3D specimen modeling and pathology report generation (*3D Model Pane*), 3) histological findings and quality report (*Histology Pane*), 4) tumor mapping and orientation to surgical specimen (*Mapping Pane*).

### ArcticAI Software Framework

The aforementioned functionality of *ArcticAI* is accomplished through a self-contained software framework, comprised of:

1. A *pip-installable* Python package (*arctic_ai*) which contains an Application Programming Interface (API) and command line interface (*CLI*) that are organized into a collection of modules:

a. 3D tissue modeling via photogrammetry (*arctic_ai.model_3d*)
b. Tissue Preprocessing (*arctic_ai.preprocessing*)
c. Histological Findings

i. Tissue Quality Prediction (arctic_ai.cnn_embeddings, generate_graph, gnn_prediction, set to macro_map mode)
ii. Tumor Margin Assessment (arctic_ai.cnn_embeddings, generate_graph, gnn_prediction, set to tumor_map mode)
iii. Ink detection and spatial statistics for tissue orientation (arctic_ai.ink_detection)
d. Tumor Confounder Identification

i. Follicle detection (arctic_ai.follicle_detection)
ii. Cell classification (arctic_ai.nuclei_detection)
e. Tumor and quality mapping onto surgical specimen (*arctic_ai.tumor_map*)
f. Image stitching (*arctic_ai.image_stitch*)
2. A collection of docker and singularity containers that host various subcomponents of the software to enable interoperability and ease installation/dependency conflicts through self-contained linux subkernels.
3. Toil job scheduling tool and workflow engine for massive parallelization across local and cloud computing clusters (*arctic_ai.workflow*).
4. A dockerized dynamic web framework that can be hosted online and interacts with the aforementioned software elements and results output through *Plotly Dash*, which contain the following panes:

a. Patient selection and workflow job submission (*Selection Pane*)
b. 3D tissue model, size and orientation measurements, smart grossing recommendations and report generation (*3D Model Pane*)
c. Histological findings–tumor and tissue quality (e.g., holes and tears), optional nuclei/follicle detection results, and detected inks placed atop slide images (*Histology Pane*)
d. Mapping of tumor and/or hole/tear results back to original specimen via computer generated surgical maps (*Mapping Pane*)

In the following sections, we will elaborate on the functionality of each of the ArcticAI modules, with reference to supplementary methods if necessary.

### Patient Selection Pane

A log-in pane allows for the selection of a patient/case. A database containing the patient and file paths to existing results data are searched and if results do not exist for the patient, the user is prompted to upload data for the *3D Model* and *Histology* panes, whichever may exist. Upon uploading, jobs are deployed to a high-performance computing cluster or within a GPU-capable device for parallel execution, which dynamically updates the database as results become available. If results exist, the *3D Model, Histology,* and *Mapping* panes become available for navigation. Here, the user is also instructed to supply the number of sections and tissue pieces per WSI based on their placement prior to image scanning.

### 3D Tissue Modeling and Grossing Recommendations in *3D Model Pane*

Three-dimensional tissue modeling prior to histological assessment provides smart grossing recommendations while automating the report of tissue size and orientation ^66^. We utilized photogrammetry techniques which triangulate image features across multiple viewpoints/images to 3D coordinates in order to generate a 3D model of the tissue. We developed a low-cost photogrammetry studio using a phone camera placed at a fixed distance away from a turntable. Immediately after resection, the tissue is placed on a turntable, from which a video of the tissue is recorded on a smart phone as it revolves around the table for one revolution. The video is then uploaded to the *ArcticAI* web app interface. Three-dimensional modeling is accomplished using:

1. **Tissue Localization:** First, the area of the turntable is approximated using RANSAC-based ellipse finding algorithm, which defines a static search area for tissue across the video frames. Then, image segmentation is performed on each video frame, which separates tissue from background using intensity thresholding, a connected component analysis for image labeling and an object size filter (**Supplementary Figure 9A**) with background removal using the *grabcut* algorithm ^67,68^. Under diverse imaging conditions, intensity thresholding can return many objects; however, only the gross specimen should follow an elliptical pattern as it completes a revolution. As such, RANSAC ellipse fitting and various fit statistics are again used to remove non-specimen objects through consideration of gross specimen’s temporal trajectory. Alternatively, segmentation neural networks, which return pixelwise coordinates of the tissue location, can also accomplish this task given training data. Here, only one-tenth the number of segmented still frames are selected for inclusion in the reconstruction algorithms to reduce the compute time. This limits reconstruction quality, but the number of frames used for reconstruction can be varied based on speed/accuracy preferences.
2. **Feature Matching:** Is accomplished with image matching (e.g., SIFT, SURF, ORB, deep feature matching ^69,70^), which can find correspondent features across different viewpoints. We utilized colmap’s SIFT implementation, which was accelerated using graphics processing units ^71–73^.
3. **3D Reconstruction:** Three-dimensional scene reconstruction using colmap’s structure from motion (SFM) framework after image pairing (i.e., match features between images from similar perspective), registration, and triangulation of pixel coordinates in a 3D cartesian coordinate system, which yields a sparse point cloud ^74^, after which a dense point cloud is generated using a Multi-View Stereo (MVS) framework via depth estimation.
4. **Distance Calibration:** Distance calibration (i.e., conversion of pixel distance to physical distance) by measuring the diameter of the turntable and fitting an ellipse (RANSAC) to the edges of the turntable, where edges were detected using a *scharr* filter (**Supplementary Figure 9B**) ^40,75,76^.
5. **Measuring orientation:** Since the 3D model is oriented randomly upon creation, the 3D model is reoriented such that the flat surface at the tissue bottom is fixed in the downward (“negative-z” direction) position and the tissue is translated to the (0,0,0) cartesian coordinate system. First, a k-nearest neighbor’s outlier detection subroutine is used to refine the point cloud. Calculation of the bottom tissue surface is accomplished through RANSAC plane fitting, where the normal plane vector is used to calculate a rotation matrix ^77^. Finally, the tissue’s 12 o’clock is calculated through segmentation of the tissue suture as a point of reference, which is used to rotate the tissue such that 12 o’clock aligns with the “positive-y” direction (**Supplementary Figure 9C**). In the absence of the suture or potential slight misalignment, the web application features a slider to allow minor rotational adjustments.
6. **Measuring Tissue Size:** Measurements of tissue size (e.g., length, width, height) are captured by calculating the maximal x-y-z extents of the tissue respectively after tissue orientation (**Supplementary Figure 9D**).
7. **Further Model Refinement:** The output 3D model retains the original color and texture of the excised tissue. The model is further refined using a Radius Neighbor’s regression algorithm, which interpolates color and texture from adjacent points while estimating the z-coordinates from a closely spaced x-y grid. Alternatively, Poisson mesh reconstruction after estimation of triangle normals and/or Delauney triangulation and alpha hull construction present alternative refinement approaches (**Supplementary Figure 9E**) ^78^.

It should be noted that the *3D Modeling* step does not model or image deep margins since the bottom of the tissue sits on the turntable, though this modeling step is entirely separate from the histological findings (which do model deep margins) and mapping those results to the surgical tumor map but may be integrated with the other two modules in future iterations.

*Grossing Recommendations and Size Report in 3D Model Pane.* The 3D tissue model is displayed using an interactive web application using the *dash_vtk* package along with exportable technical readouts on the tissue size measurements (*3D Model Pane*) ^79,80^. ArcticAI features two grossing recommendation tools, one for Mohs and another for traditional excisions with breadloafing. For the Mohs configuration, a 3D line is drawn from 12 o’clock to 6 o’clock in the web application. The 12 o’clock portion of the line is colored blue while the 6 o’clock portion is colored red. If the tissue is to be bisected, two pairs of blue-red lines are drawn parallel to a black line, which is drawn in the middle of the orientation lines. For breadloafing, the surgical excision is arranged such that the Burow’s triangles/cones (i.e., superior/inferior or lateral/medial triangular excisions adjacent to resection used as a skin graft to repair surgical defects) point in the forward/backward (“positive/negative-y”) direction. Colored lines are placed across the specimen in the side-to-side direction at regular 0.5 to 1-centimeter increments (or set by the user; based on distance to the center) to represent placement of the breadloaf section cuts. Lines to the left of the specimen are colored blue to maintain orientation, while lines to the right are colored red, yellow, green, purple and orange to denote unique sections. The tissue can be inked in accordance with grossing recommendations.

### Whole Slide Image Preprocessing

After uploading tissue image sections in Whole Slide Image (WSI) format (TIFF/SVS file format, unsigned 8-bit color), slide images are prepared for both tumor and hole/tear prediction subroutines. First, tissue mask is created using a collection of image filters via the *PathFlowAI* package ^81^. The tissue mask is generated using the following subroutine:

1. Using an intensity threshold filter, where objects of too high intensity are removed/set to white and filtering out large gray objects which may be artifactual (e.g., image scanner text, background black pen, etc.).
2. Morphology (binary closing) and blurring operations to smooth out the mask.
3. Removal of small objects and small holes.

*Patch Extraction and Assignment of Tissue Piece and Section Identifier.* WSI are typically partitioned into patches/subimages because they are too large to predict on using modern high performance computing resources with limited GPU memory. Therefore, subimages (256-pixel by 256-pixel) were extracted from the source image. Patches were extracted given that they had a significant overlap with the tissue mask as defined by a set threshold of tissue present. Patches were appended with patch metadata (e.g., x-y coordinate in WSI). The patch metadata also contains which serial section the patch belongs to (multiple serial sections per WSI). Oftentimes, each of the tissue sections were bisected or cut into four quadrants and inked separately. We refer to the resulting fragments as tissue pieces (one or more pieces per section). Each piece was placed separately in the WSI and physically close to other pieces in the same section, though there were instances where pieces either overlapped or highly separated which made it difficult to properly tag a patch with the relevant section. Tagging patches with the section they belong to is essential for tumor mapping, such that each section can be isolated from the others and then mapped by itself to the surgical tumor map after predicting the histological findings. As many WSI may be extracted per excision stage/depth, and multiple stages may be extracted during the excision procedure, the naming convention for each section denotes the depth in the specimen. Inadequate separation of sections and/or tissue pieces by failing to tag patches with the correct section identifier may degrade the performance of the tissue completeness, orientation and mapping algorithms because patches will be extracted from the space between the two conjoined section, which may contain excess whitespace and distort the ink and shape statistics. However, estimating which sections certain tissue pieces belong to is non-trivial since neighboring pieces may be conjoined, which resembles a section with fewer pieces that may fail to map well. To this end, the *preprocess* module features a robust automated section/piece splitting algorithm which can assign patches to the appropriate tissue piece/section using the following subroutine (**Supplementary Figure 10A**):

1. Tissue patches are connected based on spatial proximity, building a radius nearest neighbors graph. Sections comprised of patches are established using a connected component analysis that finds and labels contiguous sets of patches. Sections are assigned based on a large neighborhood of patches, large enough (within a 4096-pixel radius) to connect patches between neighboring tissue pieces within the section but small enough to not incorporate adjacent sections. At this point, it is difficult to delineate pieces from the section especially if they are conjoined, because what constitutes a contiguous element is defined at a large distance, and it is assumed that tissue pieces are the tissue sections, which now must be further subdivided. Then, pieces are initially broken based on connectivity at a smaller distance (within a 512-pixel radius), small enough to break apart pieces when they are separable in a section but not small enough to separate conjoined pieces. This will generate in most cases multiple pieces per section.
2. For each tissue section, if the number of tissue pieces for the section matches expectations (input parameter), then piece/section assignment for that section is complete. If the number of pieces per section does not match expectations, then the algorithm would divide largest candidate piece into the expected section pieces using Spectral Clustering, a technique that divides conjoined sections into separate areas by regions of weak connectivity between conjoined pieces. Repeat this step until optimal results are achieved.

The initial set of tissue subimages that remain after the above procedure serve as input to the tumor prediction algorithm *i.e., tumor_map* configuration), which predicts presence of tumor on a patch-by-patch basis. This configuration defines areas on a section where tissue is present from which to predict location of tumor but purposefully omits candidate holes and tears to avoid predicting those regions as benign. Thus, this set of patches is insufficient to predict where tissue is absent or incomplete (i.e., tissue quality, location of holes and tears that determine whether the section should be assessed) since patches are, by definition, absent.

Patches correspondent to candidate holes and tears are extracted for each section piece using an *alpha shape* object finding algorithm which outlines the section piece in a way that is both tightly fit to the piece while also bridging tears that are connected to the exterior of the object and thus are not normally estimated using traditional hole finding algorithms (**Supplementary Figure 10B**) ^78^. These patches are added to the *tumor_map* patches to form the *macro_map* configuration for tissue quality / tissue completeness assessment. If tissue is incomplete, it is inadvisable to assess margins. All tissue piece subimage patches, tissue masks and their corresponding metadata are written to NPY format (numpy array ^82^) and serialized into pickle format respectively for storage.

### Feature Extraction using Convolutional Neural Networks

We trained *ResNet-50* convolutional neural network (CNN) models to extract predictors from the tissue subimages to be used in our prediction workflow ^83^. First, convolutional neural networks were trained and internally validated on a subset of image patches (n=122 WSI; 1,988,841 patches) for the following prediction tasks, with a batch size of 32 patches, learning rate of 1e-4, modulated with a cosine annealing learning rate scheduler for 100 training epochs:

1. *Tumor CNN:* Tumor localization, where regions of tumor were delineated from benign structures and inflammation. If a patch contained both malignant and inflammatory cells, the patch was labeled as having contained tumor.
2. *Completeness CNN:* Delineation of macro-architectural subcompartments, including: 1) hole/tear, 2) fat, which if not explicitly annotated, could closely resemble hole/tears, 3) epidermis and 4) dermis. Here, patches of dermis containing wispy white patterns were removed from the training/validation set to avoid conflating regions of dermis with hole/tear.

After training, the two ResNet50 CNN models were used to extract *embeddings* of image features from the penultimate layer of the model as images passed through the neural networks. CNNs were organized into multiple processing layers, each of which represent objects/images at increasing levels of abstraction (i.e., each input register corresponds to a more complex image feature at a deeper layer). Whereas the final CNN layer is used to output the probability of the presence of a specific tissue architecture, the penultimate layers output a rich feature set (*embeddings*) which can be used as a generic representation of the image features and if plotted could demonstrate how specific images cluster together with dimensionality more expressive than the output layer alone. The trained CNN models, *Tumor CNN* and *Completeness CNN*, are configured to output 2048-dimensional *embeddings* for all *tumor_map* and *macro_map* patches respectively for a given tissue section. CNN models were configured using the *PyTorch* package (v1.8.0) using Python v3.7 ^84^.

### Graph Neural Networks for Final Histological Assessment

WSI contain significant white space and the placement of tissue on a slide is relatively arbitrary. The dimensions of WSI are typically very large which necessitates dividing the tissue into smaller subimages (tiles). However, prediction using a CNN assumes that neighboring image patches are unassociated, which undervalues their spatial context within the surrounding tissue architecture. Graphs represent image patches and their spatial dependencies as nodes and edges, capturing both feature and spatial information. As such, graph-based neural networks (GNN) have emerged as premier methods for histological assessment. Using GNN’s, predictions are invariant to the positioning and orientation of the tissue and are enhanced by the incorporation of spatial information encoded in the edges. We fit two GNN models corresponding to the following prediction tasks, with a batch size of 16 WSI graphs, learning rate of 1e-2, modulated with a cosine annealing learning rate scheduler for 1,500 training epochs:

1. *Tumor GNN:* Analogous to *Tumor CNN*.
2. *Completeness GNN:* Analogous to *Completeness CNN*. However, all patches are included in this analysis, including wispy dermis which is now contextualized by the surrounding dermis and not subject to conflation with holes/tears.

Graphs were defined using a radius neighbors algorithm, which connected patches (nodes) to their immediate neighbors (edges) using their positional x-y coordinates ^41^. Attributes of the graph nodes were set to the *embeddings* extracted by the relevant CNN. Node attributes (CNN features) were shared/passed to adjacent patches using three graph attention layers of dimensionality 32, 32, and 64. The graph convolution layers were interspersed with *DropEdge* and *Dropout* layers, which randomly pruned patch-wise connections and graph-learned node features during training (to enhance model robustness to noise). After running the graph convolutional layers, these features were piped to a prediction output layer that would return predicted probabilities (and their logits) of each class for the respective tasks. GNN models were configured using the *PyTorch-Geometric* package (v1.7.1) using Python v3.7 ^42^.

### Ink Detection and Calculation of Spatial Statistics for Tissue Orientation

The orientation of the WSI tissue section with respect to the original specimen / surgical tumor map was inferred using spatial statistics / tissue orientation algorithms. These algorithms were developed to automatically identify ink colors and orient a WSI tissue section based on a collection of applied inks: blue and red, though subroutines exist in *ArcticAI* to additionally calculate: yellow, green, orange, purple, and black. First, tissue edges were segmented using a Sobel filter with morphological dilation and opening operations. Then, sensitivity analysis over thresholds in Hue, Saturation, Value (HSV) color space yielded optimal color thresholds to detect inks, which were paired with a connected component analysis to identify contiguous regions and remove spurious applications of ink within the tissue edges (i.e., where ink is erroneously applied / seeps). Alternatively, semantic segmentation algorithms based on annotations, and conditional random field ^85^ can further improve ink detection. After detecting ink, orientation of the WSI section piece is inferred through calculation of the center of mass of the x-y coordinates of each detected ink color (using either the mean, median, or trimmed mean of each pixel coordinate). In our practice setting blue defines 12 o’clock, red defines 6 o’clock, in accordance with the 3D model of the Mohs specimen and with the surgical tumor map. The line between the blue and red ink defines tissue orientation relative to blue and red locations defined in the surgeon’s hand-drawn tumor map, where the relative angular difference between the blue-red lines from the histology and blue-red lines from the tumor map dictate the relative rotation required for the histology results to match the same angle of the tumor map.

### Image Stitching

Input WSI are prepared for viewing using a subroutine which converts the images of individual sections, extracted using the preprocessing workflow, to a “Deep Zoom Image” (DZI) format, a pyramidal file format which interfaces with *openseadragon*, a WSI viewer. The aforementioned rapid histological assessment steps (Preprocessing, *CNN-GNN*, *Ink Detection*) return positional predictions for their respective coordinates. These positional predictions are piped and prepared for display through a dynamic json export of the histological results and imported into an *openseadragon* SVG overlay component for viewing across the slide ^86^.

### Removal of Potential Tumor Confounders and Identifying Residual Tumor in Regions of Predominant Inflammation

Two R101-FPN neural network models using the computer vision framework *detectron2* (for panoptic segmentation) were trained for the task of localizing follicles across a slide and residual tumor within pockets of inflammation as an added layer of auditing. Panoptic segmentation models can detect objects in an image and their image class through proposals of bounding boxes using neural network detected features while simultaneously segmenting the object using a segmentation architecture which operates dynamically on the proposed regions ^43,44^.

First, 672 follicles were annotated across 16 WSI from the *ArcticAI* training/validation sets, where 595 non-overlapping 1024 pixel by 1024-pixel subimages were extracted and assigned to training and validation sets whether they belonged to WSI of the training or validation set for the CNN-GNN algorithms. A panoptic segmentation network was fit to the data at a starting learning rate of 1e-3 and was trained for 1000 epochs. The trained neural network was applied to patches suspected to contain tumor in the test set WSI to eliminate patches with significant confounding. This was done using an adjustment scheme in which tumor scores were reduced more for patches with greater proximity to the follicle based on the overlap between the predicted follicle and three concentric circles (128, 256, 512-pixel radii respectively) around each patch. The percentage overlap with the follicles and each circle multiplied by a circle specific penalty (higher for the 128-pixel circle and lower for the 512-pixel radius circle), normalized to a scale between zero and one was used to determine the tumor prediction probability to dock from the original score.

Inflammatory patches were assessed at the level of individual nuclei via the creation of a workflow to detect, classify, and segment nuclei. Using the nuclei annotations that were manually annotated by four pathologists using the Automated Slide Analysis Platform (ASAP), we extracted 795 patches of size 128 by 128, correspondent to approximately 32,763 nuclei, across three whole slide images (WSI) from the training/validation set. The model was trained to detect and delineate the following cell types: 1) fibroblasts, 2) hair follicles, 3) inflammation, 4) malignant basal cells, and 5) benign epidermal keratinocytes. For our train-validation split, 80% of these patches were randomly chosen for our training dataset and the algorithm was validated using the remaining 20% of the image patches prior to prediction on the test WSI. We reported the predictive statistics for detection accuracy on an internal validation set using the Dice score (related to Intersection over Union). Predicted cell types were refined from the detected nuclei using a CNN (ResNet50 architecture) and GNN model using the same training/validation sets. F1-score statistics were recorded as a final measure of fit across the test set cells, separately comparing the detection, CNN and GNN models for their prediction accuracy, bootstrapping on the patch and slide level since the nuclei are nested within patches.

Here, the validation set was not used for optimal hyperparameter scanning nor model early stopping criterion, so we evaluated results on this held out validation set prior to application across test slides. We timed both algorithms through evaluation across test set WSI and similarly recorded the uncertainty through non-parametric bootstrapping while accounting for clustering on the WSI level.

### Compilation of Histological Assessment Results into *Histology Pane*

The results from the histological assessment models (*Tumor CNN-GNN, Completeness CNN-GNN*) are passed to an OpenSeadragon plugin that features the DZI image correspondent to the selected case, resection site/stage and section depth. This plugin operates within a *plotly dash* environment that interfaces the WSI viewer with the results data ^87^. The user has the option of selecting whether to display a heatmap containing the tumor or completeness (holes/tears) prediction results over the slide using the SVG plugin as aforementioned. A slider controls the minimal prediction probability for inclusion in the heatmap to filter out irrelevant regions. The patches’ color intensity (blue to red and opacity) is determined by their prediction scores. The predictions from image patches can be optionally refined using an interpolation method which uses a custom prediction propagation GNN to yield refined predicted probabilities that exist between the original patches (e.g., if we had four patches in locations (1024,256), (1024,512), (1024,768), (1024, 1024), we could infer information at tile position (1024, 640) by leveraging information from all four tiles, though primarily from adjacent tiles) ^88^. The SVG display can be toggled on and off. The tissue orientation may also be toggled on and off, where red and blue lines may be automatically placed on the slide based on detected inking patterns and associated spatial statistics.

### Mapping Tumor and Completeness Results to Surgical Specimen using the Mapping Pane

Results (tumor/completeness) from user selected tissue sections for each case can be mapped to the surgical specimen that is featured on a hand-drawn surgical tumor map at arbitrary locations using an interactive image display. First, the user selects from a set of prepopulated surgical map templates representing various anatomical positions (e.g., back of hand, neck). After selecting the position template, the user draws a black ellipse on the image template representing the removal site. The user also defines tissue orientation by drawing blue and red inks at the circle’s edge to define 12 o’clock and 6 o’clock respectively, which is correspondent to inking patterns recommended/selected from the *3D Model* and *Histology* panes. Tissue sections comprised of 1-2 tissue pieces are represented by a 2D point cloud or a collection of points, where each point is tagged with positional x-y coordinates within the WSI, the tumor/hole/tear predicted probabilities and ink locations. These points are “morphed” or registered to locations in the interior of the circle using an optimizer for optimal transport, which minimizes the cost or effort required to match the points to the interior of the circle while maintaining the relative positioning of the coordinates comprising the histological section. The distributional difference between the histological section and surgical mapping ellipse is estimated using the sliced Wasserstein (“Earth Movers”) distance and minimized using gradient descent via *pytorch* and *python optimal transport (POT)* libraries ^47–49,84^. In sum, this methodology morphs the arbitrary shapes of the histological specimen, which are dependent on serial sectioning of the gross specimen, to the elliptical shape drawn by the Mohs surgeon. The relative positioning of ink is preserved during this transformation and the angular difference between the ink after tissue morphing and that defined via the *Mapping pane* are used as a final rotational adjustment to match the surgical tumor map. Finally, the histological section results are placed in the circle on the *Mapping pane* in the correct orientation, where a kernel density contour map defined over the tumor/completeness results is placed to highlight tumor/holes/tears. Like the *Histology pane*, the density map can be thresholded with arbitrary probability cutoffs by the user to yield specific tumor locations and the finalized map can be exported to the pathology report.

### Workflow Specification

All aforementioned *ArcticAI* jobs execute using a Toil job executor ^50^, which can run jobs in parallel either locally (on a GPU-capable machine) or in an HPC environment using a Slurm or alternative job submission system. Here, we will enumerate which components execute in parallel based on their respective workflows, where we have denoted which set of subcomponents execute in series or parallel.

1. *3D Model Pane* (**series**):

a. Tissue preprocessing (series)
b. 3D Reconstruction (series)
c. Final tissue filtering (series)
2. *Histology Pane (**series**)*:

a. Tissue preprocessing and section assignment (series)
b. Parallel components, where final subworkflow time is assessed by the subcomponent and tissue section which took the longest time to execute (below subpoints are **parallel**)

i. CNN-GNN subcomponents for tumor/completeness prediction, comprised of CNN embedding creation, graph generation and GNN prediction (parallel)
ii. Ink detection and orientation (parallel)
iii. Image stitching (parallel)
c. Based on CNN-GNN results (**parallel**)

i. Follicle detection (parallel)
ii. Nuclei detection (parallel)

Finally, we have included a *Histology* workflow diagram which illustrates how results from various workflow components feed into subsequent steps (**Supplementary Figure 8**). It should be noted that after tissue preprocessing, tissue pieces/sections all execute in parallel, from which the aforementioned subworkflows also execute in parallel.

### Experimental Objectives

#### Tissue Grossing Measurement Concordance

Length, width and height measurements of the 3D reconstruction of resected tissue was compared to hand measurements of the original specimen using median absolute deviation and spearman correlation statistics. These statistics were also recalculated under the assumption that the calculated tissue dimensions were off on all 3 dimensions by a proportional constant (i.e., improper calibration of the video with distance measurements).

#### Concordance of Algorithm to Hand-Drawn Maps from Surgeon

To assess the accuracy of the Arctic grossing, completeness, and tumor detection algorithms and mapping of histological findings to the surgical tumor map, we included the following comparisons, assessing accurate: 1) calculation of tissue size, 2) analysis of tissue quality or completeness of tissue as judged by the localization of holes and tissue tears, common to frozen specimens, 3) localization of tumor in WSI, 4) orientation of tissue section, 5) mapping of tumor to surgical tumor map, and 6) a prediction of whether and where additional tumor removal is required.

In comparison to previous studies which examine the diagnostic significance of positive margins on post-operative BCC sections, we assessed whether pathologists would manually map tumors similarly in digital versus analog mediums by comparing hand-drawn tumor maps to digital ones. After establishing concordance between histological findings via pathologist annotations and BCC predictions, we established the concordance between the automated tumor map with the intraoperative hand generated map that was produced by the Mohs Micrographic Surgeon at the time of surgery. If there were discrepancies between surgeon-generated and ArcticAI-generated tumor maps, both the original glass slides and WSI were manually reviewed.

A receiver operating characteristic curve (ROC; sensitivity analysis) was performed to establish predictive probability cutoffs which result in high sensitivity to minimize the potential for false negatives. Results are reported with a 1000-sample non-parametric bootstrap 95% confidence interval with bootstrapping performed on the WSI level to capture clustering on the slide level (i.e., variation in performance statistics between and across slides).

Separately, concordance between hand-drawn and digital tumor maps was calculated based on the proportion of cases the surgeon subjectively rated as equivalent (orientation of map and position of tumor) to the original map. Uncertainty in this proportion was assessed through calculation of the 95% credible interval (CI; like the confidence interval) of a Beta posterior distribution (*Beta*(*a* = 0.5 + 28, *b* = 0.5 + 0)), updated through a beta-binomial conjugate prior, with a Jeffrey’s prior (*Beta*(0.5,0.5)) and Binomial likelihood (*Binomial*(*n* = 28, *p* = 1.0); 28 cases with positive margin, 28 successful trials; three cases had clear margins).

#### Execution Time

To demonstrate the timely execution of the ArcticAI system, the following steps in the process were precisely timed: 1) image preprocessing; 2) tissue quality and tumor CNN-GNN prediction; and 3) tumor mapping and pathology report output. We report median times across slides to account for outliers, with 1000-sample nonparametric bootstrap 95% confidence intervals. Details on the calculation of timing given optimal parallelization can be found in the methods section, section “Workflow Specification”. Test cases were evaluated using four compute nodes in the Dartmouth Discovery computing cluster which shared between them 13 Nvidia v100 GPUs (32 Gb memory each), 272 CPUs, and 1.9 TB RAM.

## Supporting information

Supplementary Discussion

## Data Availability

The datasets presented in this article are not readily available because of participant privacy concerns. Requests to access the datasets should be directed to.

## Acknowledgements

We would like to thank John Kim, Adnan Murtaza, Sagar Gupta, Sachin Satishkumar and Aryan Kumawat from the EDIT Machine Learning Summer program for initial explorations of 3D modeling of common everyday objects using Meshroom. We would also like to acknowledge the help of the Dartmouth Hitchcock Department of Dermatology Mohs surgical staff, for data collection of gross tissue specimen. JL is funded in part under NIH subaward of grant number P20GM104416.

