## Supplementary material for "ArcticAI: A Deep Learning Platform for Rapid and Accurate Histological Assessment of Intraoperative Tumor Margins": Tables

**Table 1: Case Count:** Number of cases, slides, tissue sections, pieces and annotations comprising the training/validation and test sets for the margin assessment (completeness, tumor localization) and ink orientation algorithms, where number of WSI are also broken down by BCC histological subtype and clear margins (in the test set as controls). Note that some of the WSI featured in the BCC subtype breakdown are double counted, as they represent several histological subtypes

| | Tumor and Holes/Tears | | | Test Set: Inking Patterns |
|---|---|---|---|---|
| | **Total** | **Training/Validation** | **Test** | **Tissue Orientation** |
| **Cases** | 178 | 76 | 41 | 31 |
| **Slides** | 351 | 122 | 121 | 108 |
| **Sections** | 1065 | 381 | 360 | 324 |
| **Tissue Pieces** | 1537 | 472 | 559 | 506 |
| **Number Annotations** | 16,128 | 11,343 | 2,535 | 2,250 |

| | **Training/Validation** | **Test** |
|---|---|---|
| **Superficial** | 36 | 21 |
| **Nodular** | 102 | 58 |
| **Micronodular** | 11 | 6 |
| **Infiltrative** | 20 | 23 |
| **Sclerosing** | 6 | 0 |
| **Microcystic** | 2 | 2 |
| **Squamatized** | 0 | 3 |
| **Nodular/Micronodular** | 0 | 1 |
| **No tumor** | 0 | 45 |

**Table 2: Tissue size measurements (length, width, height)**, measured empirically (left) and predicted with the automated tissue grossing tool, in centimeters (cm). Also reported was whether the excision type was a radial section for breadloafing or circumferential excision for the assessment of peripheral margins.

| **Specimen** | **Excision Type** | **Measured (cm)** | | | **Predicted (cm)** | | |
|---|---|---|---|---|---|---|---|
| | | L | W | H | L | W | H |
| **0748** | Radial | 2.4 | 1.3 | 0.6 | 2.38 | 1.23 | 0.79 |
| **0749** | Radial | 4.5 | 4 | 3.3 | 4.86 | 3.61 | 2.61 |
| **0750** | Peripheral | 2.8 | 2 | 2.2 | 2.43 | 2.17 | 1.73 |
| **0751** | Radial | 4.5 | 2 | 2.5 | 4.64 | 2.52 | 2 |
| **0752** | Peripheral | 4.5 | 3.5 | 3 | 4.87 | 3.49 | 2.98 |
| **0753** | Peripheral | 2 | 2 | 0.6 | 2.02 | 1.98 | 1.05 |

| | | | | | | | |
|---|---|---|---|---|---|---|---|
| **0754** | Peripheral | 3.5 | 2.5 | 2 | 3.97 | 3.3 | 1.9 |
| **0756** | Radial | 2 | 1 | 0.7 | 1.68 | 0.88 | 0.45 |
| **0796** | Radial | 3 | 1.5 | 0.5 | 3.84 | 1.98 | 0.78 |
| **0798** | Radial | 3 | 1.5 | 0.5 | 2.47 | 1.27 | 0.85 |
| **6693** | Peripheral | 1.5 | 1.3 | 0.7 | 1.54 | 1.28 | 0.8 |
| **6694** | Peripheral | 2.9 | 2.4 | 1.1 | 2.86 | 2.2 | 1.09 |
| **6696** | Peripheral | 2 | 1.5 | 0.6 | 2.23 | 2.06 | 1.14 |
| **6697** | Radial | 5.1 | 1.7 | 0.6 | 3.46 | 1.26 | 0.79 |
| **6698** | Radial | 2.8 | 1.3 | 0.7 | 3.96 | 2.03 | 1.34 |
| **6699** | Radial | 1.7 | 1 | 0.5 | 1.91 | 1.13 | 0.79 |
| **6700** | Peripheral | 2.1 | 1.3 | 1 | 3.59 | 2.05 | 1.53 |

**Table 3: Model performance and concordance with pathologist/surgeon annotations for the gross measurements, tissue orientation/mapping, completeness, and margin assessment tasks.** Macro-AUC represents reporting of AUC statistic on slide level and averaging across slides, giving each slide equal weight, while normal AUC statistic is calculated for subimages across all slides. 95% confidence intervals were acquired using 1000-sample non-parametric bootstrap, where bootstrapping was done on the WSI level to account for variation in concordance across the cases.

| Task | Evaluation Metric | Subtask | Estimate | 2.5% CI | 97.5% CI |
|---|---|---|---|---|---|
| **Gross Measurements** | Median Absolute Deviation (cm) | L | 0.36 | 0.21 | 0.53 |
| | | W | 0.23 | 0.12 | 0.52 |
| | | H | 0.29 | 0.19 | 0.50 |
| | | Overall | 0.29 | 0.2 | 0.47 |
| | Median Absolute Deviation* (cm) | L | 0.17 | 0.14 | 0.33 |
| | | W | 0.12 | 0.06 | 0.24 |
| | | H | 0.14 | 0.09 | 0.30 |
| | | Overall | 0.14 | 0.12 | 0.25 |
| | Median Proportional Change (%) | L | 12.4 | 7.9 | 17.7 |
| | | W | 13.0 | 8.3 | 32.0 |
| | | H | 31.7 | 20.0 | 58.0 |
| | | Overall | 19.0 | 12.6 | 33.2 |
| | Median Proportional Change* (%) | L | 7.0 | 3.6 | 13.4 |
| | | W | 9.3 | 2.7 | 15.1 |
| | | H | 16.8 | 12.3 | 26.8 |
| | | Overall | 11 | 7.9 | 15.9 |
| | Correlation* | L | 0.957 | 0.827 | 0.993 |
| | | W | 0.961 | 0.854 | 0.994 |
| | | H | 0.822 | 0.448 | 0.973 |

| | | | | | |
|---|---|---|---|---|---|
| | | Overall | 0.911 | 0.766 | 0.968 |
| **Tissue Orientation** | Median Absolute Deviation (degrees) | | 4.883 | 4.04 | 5.451 |
| | Proportion Correct Orientation (≤45° difference) | | 94.7% | 92.3% | 96.6% |
| **Tissue Completeness** | AUC | | 0.839 | 0.825 | 0.855 |
| | Macro-AUC | | 0.851 | 0.839 | 0.863 |
| **Margin Assessment** | AUC | Original Slides | 0.967 | 0.960 | 0.979 |
| | | Follicle Removal | 0.965 | 0.959 | 0.975 |
| | Macro-AUC | Original Slides | 0.962 | 0.954 | 0.970 |
| | | Follicle Removal | 0.957 | 0.949 | 0.964 |
| **Tumor Mapping** | Accuracy | | 0.992 | 0.915 | 0.999 |

* adjusted for proportional constant from improperly calibrated turntable dimensions (ellipse major axis dimensions in pixels to physical measurement of turntable major axis in video)

**Table 4: Average execution time for workflow subcomponents.** Final times for a case were given by the maximum compute time across sections for the case after preprocessing. After preprocessing a WSI, the CNN-GNN, Tissue Orientation and Image Stitching tasks execute in parallel, as do all sections in the specimen, where the section that takes the longest serves as the bottleneck. Within the CNN-GNN tasks, the CNN-GNN (broken into serial CNN, graph generation, and GNN subcomponents) for tissue completeness and tumor run in parallel. 95% confidence intervals for median time statistics were estimated via 1000 non-parametric bootstrapped resamplings

| **Task** | **Subtask** | **Median(s)** | **2.5% CI** | **97.5% CI** |
|---|---|---|---|---|
| **Tissue Preprocessing** | | 47.68 | 44.21 | 51.22 |
| **CNN-GNN** | **Total** | 8.84 | 7.05 | 11.4 |
| | **Completeness CNN** | 4.31 | 4.12 | 4.5 |
| | **Tumor CNN** | 5.58 | 5.28 | 5.9 |
| | **Completeness Graph Generation** | 0.32 | 0.3 | 0.35 |
| | **Tumor Graph Generation** | 0.34 | 0.33 | 0.36 |
| | **Tumor GNN** | 0.48 | 0.45 | 0.55 |
| | **Completeness GNN** | 0.43 | 0.41 | 0.49 |
| **Tissue Orientation** | | 20.96 | 18.52 | 24.15 |
| **Image Stitching** | | 24.39 | 22.22 | 28.81 |
| **Total per WSI** | **Parallel** | 71.57 | 66.4 | 79.23 |

| | | | | |
|---|---|---|---|---|
| **Total per Case** | **Parallel** | 78.49 | 65.74 | 87.73 |
| | **Series** | 493.82 | 367.49 | 553.44 |
