## Supplementary Figures and Tables for "ArcticAI: A Deep Learning Platform for Rapid and Accurate Histological Assessment of Intraoperative Tumor Margins"

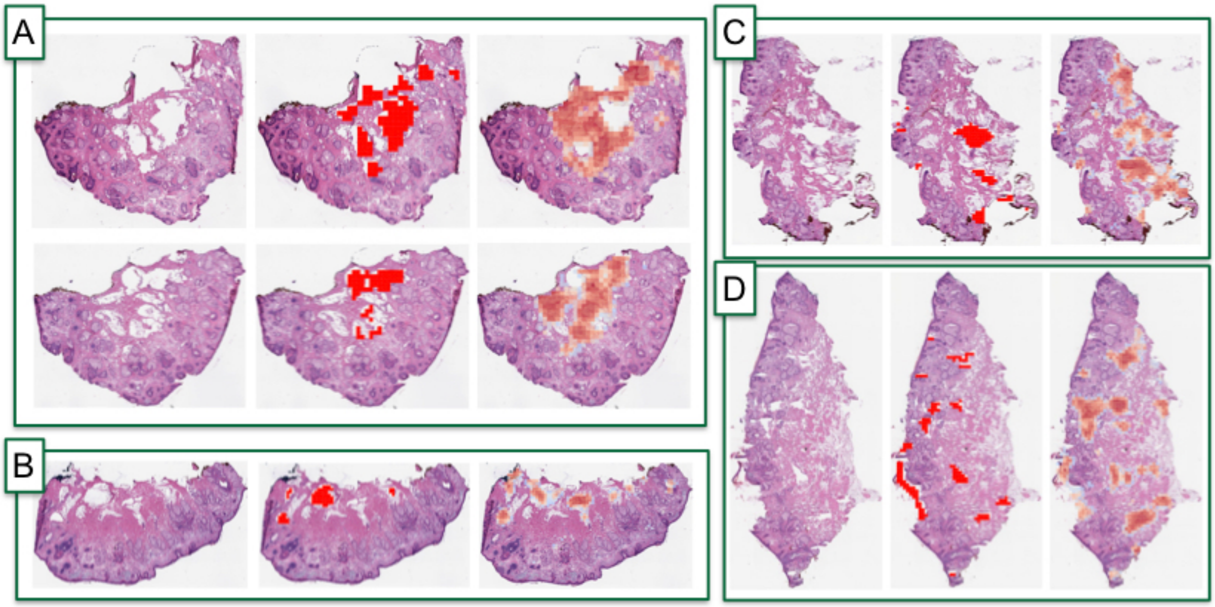


**Supplementary Figure 1: Examples of holes and tears missed/overcalled:** A-D) represent sections randomly selected from four separate cases; left image is original WSI; middle image annotated holes/tears from the pathologist are denoted in red; right image overlaid is a heatmap where degree of red for an image subarray indicates predicted probability of incompleteness by Completeness GNN

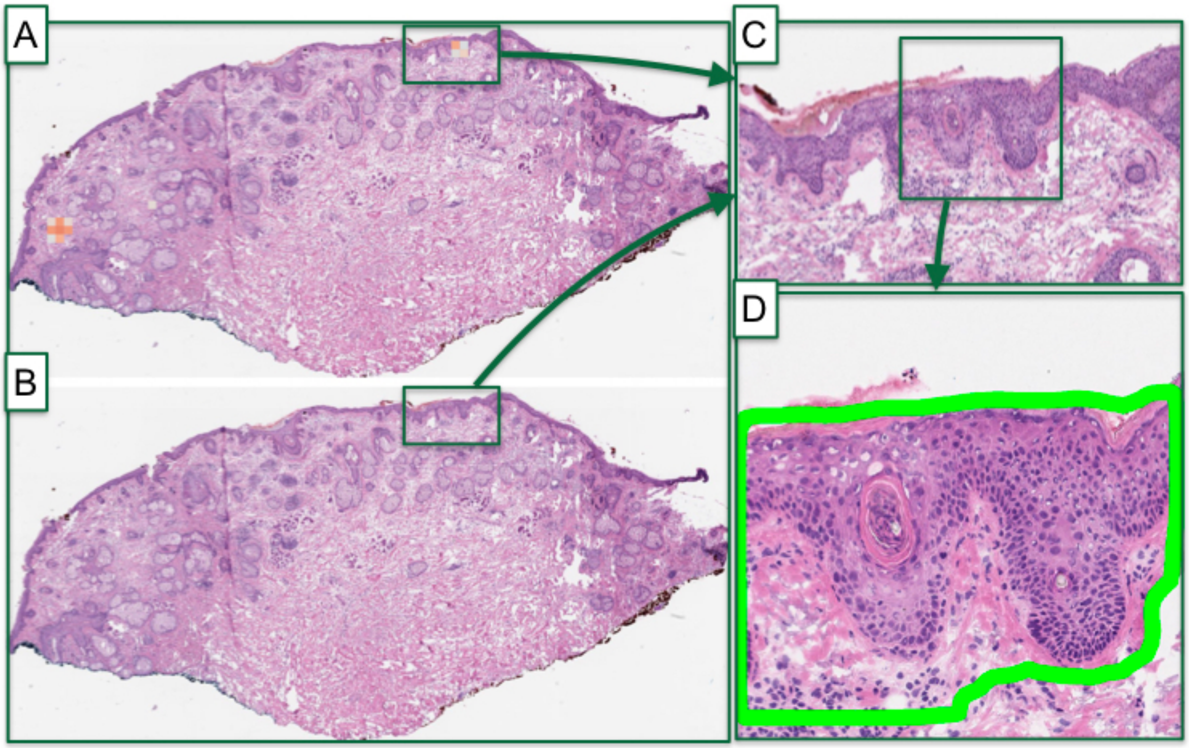

**Supplementary Figure 2: Example of follicle prediction impact on half of one tissue section:** A) heatmap is overlaid on WSI to denote predicted tumor probability (red) by Tumor GNN; B) tumor probability heatmap after accounting for presence of follicles as predicted by the follicle detection neural network; note how in this slide tumor is no longer predicted, which results in an increased area under the curve of 0.05 across the WSI for the case; C) zooming in on one tumor-predicted region called follicle by the follicle detection algorithm; D) zooming in on specific subarray where follicle was predicted by follicle detection network, where predicted follicular structure is outlined in green by the neural network

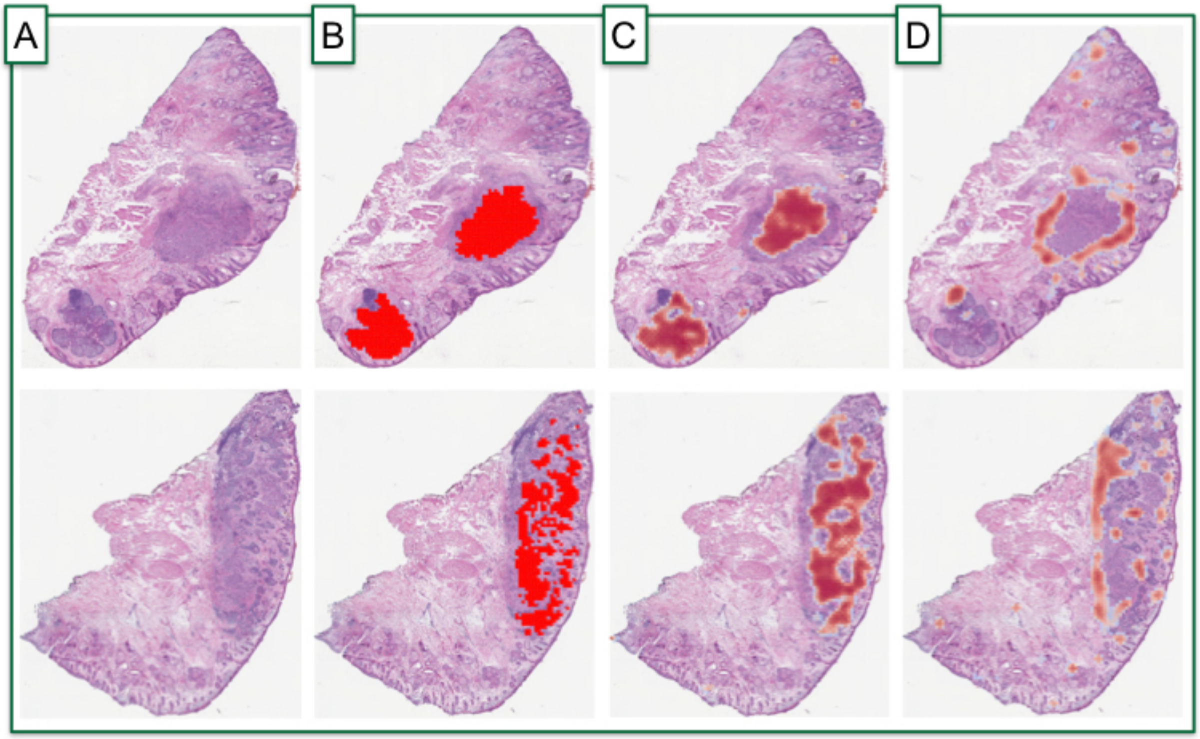


**Supplementary Figure 3: Prediction of inflammation on two separate tissue sections to illustrate importance of taking inflammation into account for tumor prediction:** A) original slide image; B) ground truth / annotated tumor denoted in solid red regions; C) predicted tumor regions given by heatmap where degree of red reflects tumor probability via Tumor GNN; D) predicted inflammatory regions given by heatmap where degree of red reflects inflammatory probability via Tumor GNN

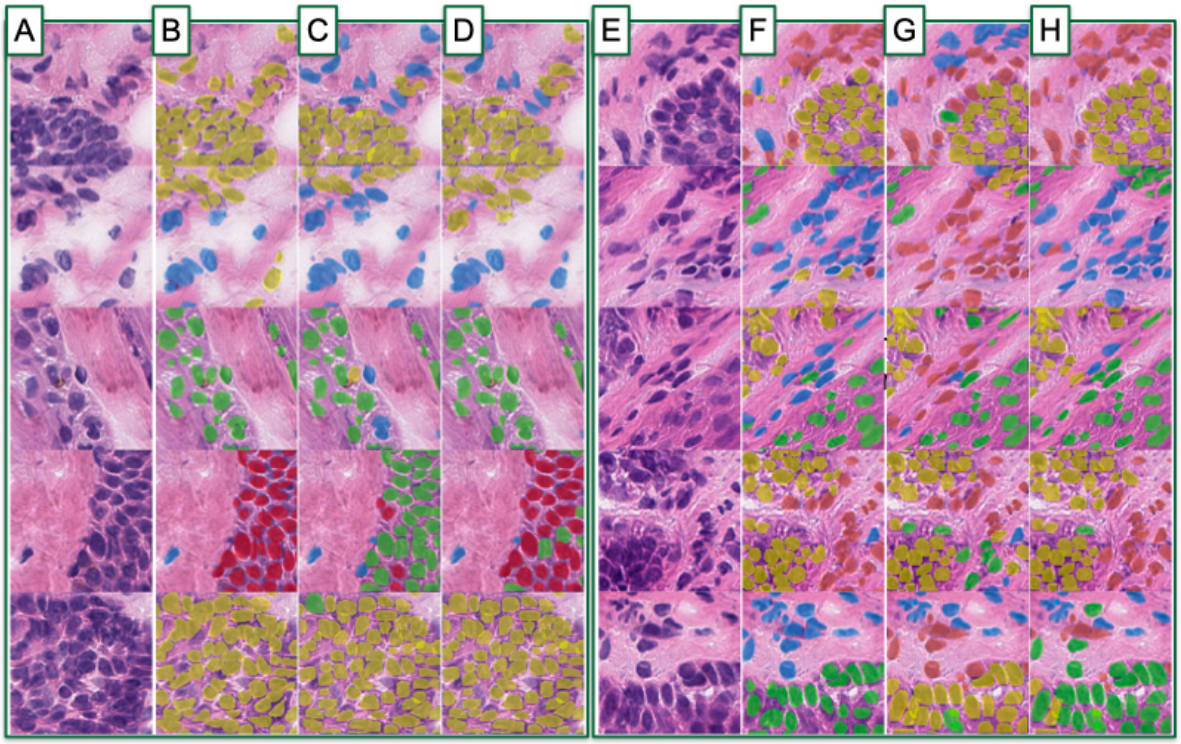


**Supplementary Figure 4: Example outputs from nuclei prediction workflow to delineate BCC (yellow), hair follicle (green), inflammatory (orange), fibroblast (blue), and epidermal keratinocyte cells (red):** A,E) original subimages; B,F) overlaid annotated cells and cell assignments; C,G) Detectron2 predicted cell assignments; D,H) Cell graph neural network predicted cell assignments

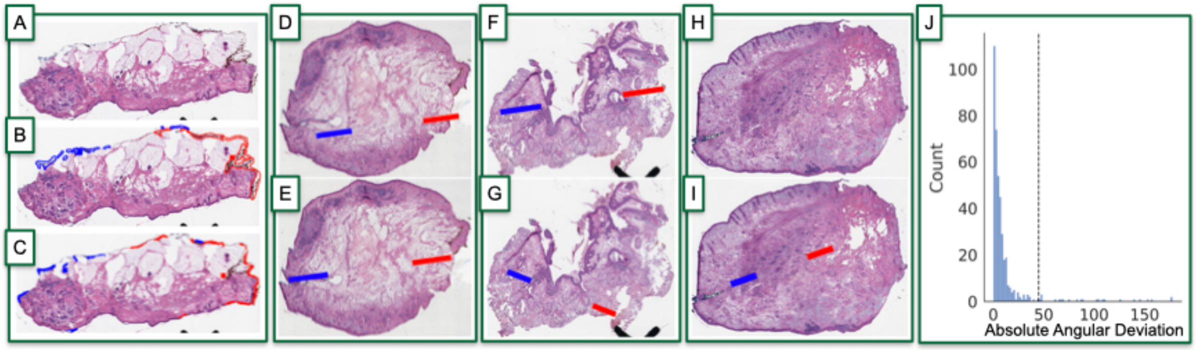


**Supplementary Figure 5: Tissue orientation compared to ground truth:** A) Original slide image; B) pathologist annotation of red and blue inks given by outlined splines; red and blue squares denote center of mass for red and blue inks respectively; C) ink detector's prediction of red/blue ink and ink centers of mass; D-E) example of optimal ink detection and orientation; D) tissue orientation (i.e., line between red and blue inks) as inferred from pathologist's annotations; E) tissue orientation (i.e., line between red and blue inks) as predicted by ink orientation algorithm; F,G) example of suboptimal ink detection and orientation due to sectioning quality; with same comparison from D-E); H,I) example of suboptimal ink detection and orientation due to missing red ink on right side; while pathologist was unable to annotate H), the algorithm still managed to report orientation for I); J) histogram denoting angular difference between pathologist-annotated orientation and algorithm-predicted orientation, in degrees, where each element reflects tissue section

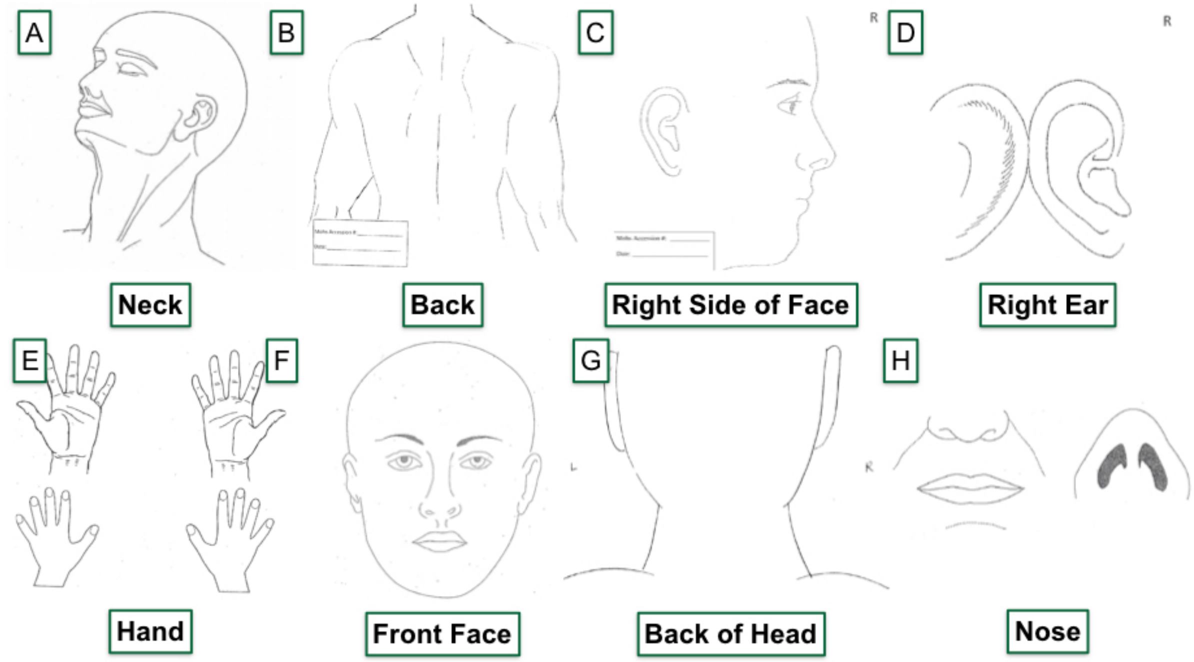


**Supplementary Figure 6: Example templates of anatomic locations for surgical tumor mapping:** A-F) examples of different anatomical locations that can be selected by the user for real-time mapping

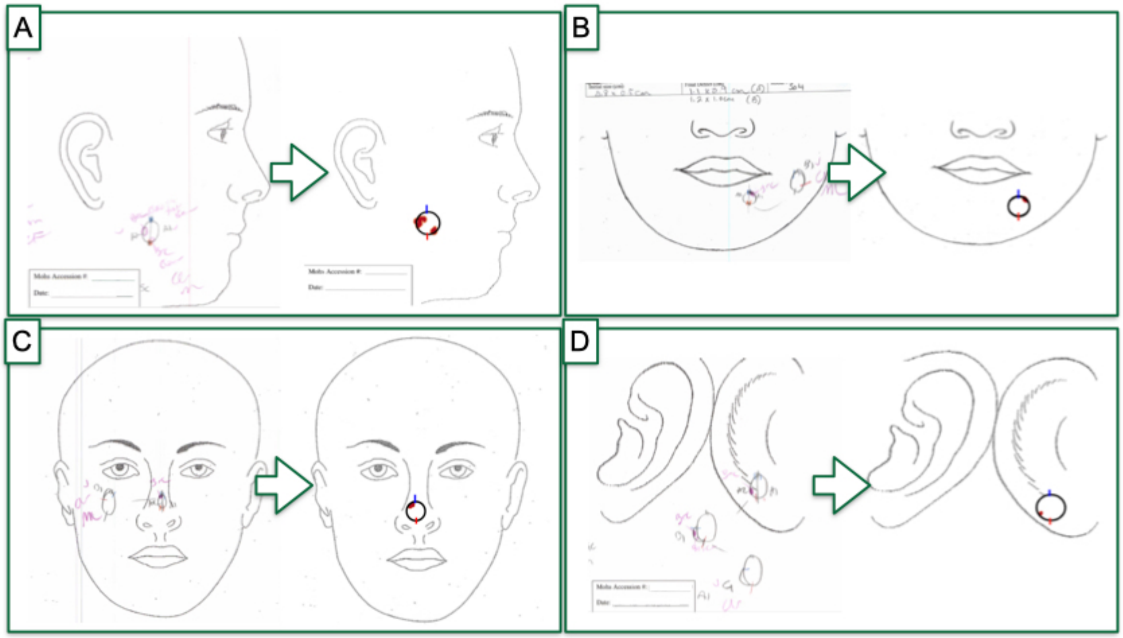


**Supplementary Figure 7: Comparison of hand-drawn surgical tumor maps (left) to algorithm predicted tumor maps (right):** A-D) four separate cases, predictions from first site predictions compared to ground truth

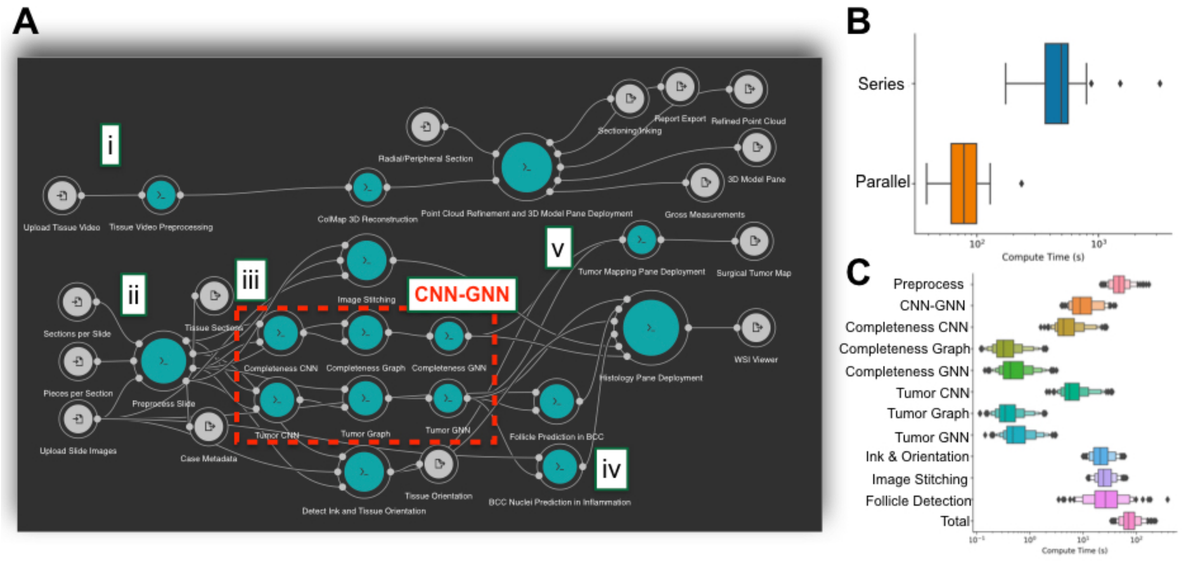


**Supplementary Figure 8: Workflow diagram and speed:** A) *ArcticAI* workflow diagram, visualized using the Rabix Composer: i) 3D modeling subworkflow, comprised of tissue preprocessing, ColMap reconstruction and point cloud refinement and application deployment; ii-v) histological assessment and tumor mapping subworkflows; ii) slide preprocessing; each tissue section in parallel passes through iii) image stitching, CNN-GNN, and ink/orientation algorithms, which are themselves executed in parallel; iv) optional follicle and nuclei prediction based on Tumor GNN results; finally, results from all sections/WSI are combined for visualization using v) the Histology and tumor mapping panes; B) boxplot denoting total execution time per case for ii-v), given serial and parallel workflow execution; C) boxenplot denoting execution time of ii-v) subcomponents and total given optimized parallel execution across WSI/sections

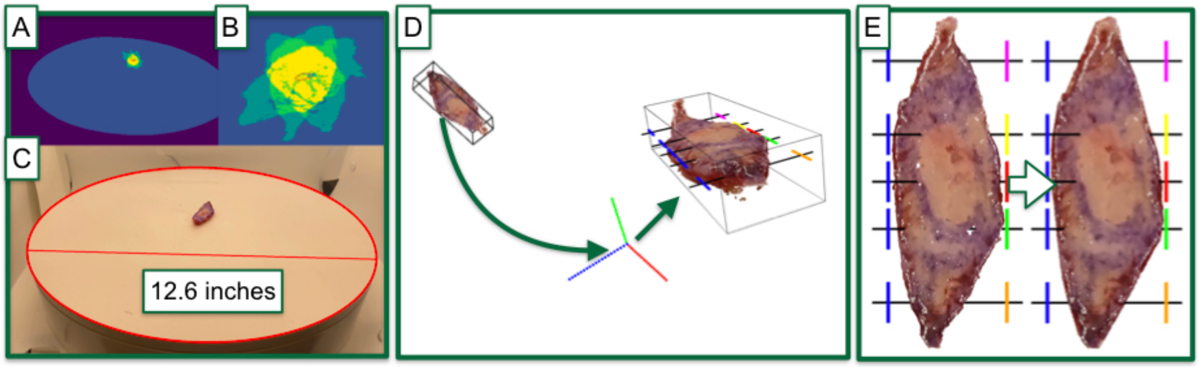


**Supplementary Figure 9: Illustration of automated 3D tissue modeling workflow steps:** A-C) tissue video preprocessing; A) tissue (yellow; imaged across multiple timesteps) and turntable (blue) are identified in video; B) orange ellipse in middle charts path of center of mass of tissue, used to help delineate tissue versus other image objects; C) calibration of image pixels to length measurements, where red ellipse and line are results from automated ellipse detection algorithm, where double the major axis length is approximated to be the diameter of the turntable, 12.6 inches; D,E) 3D model post-processing; D) reconstructed tissue is initially placed at arbitrary orientation; tissue is filtered and rotated/aligned to origin to automate placement of tissue sections/inks;

size is recorded using bounding box; E) 3D tissue model is additionally refined via interpolation to smooth the model for viewing

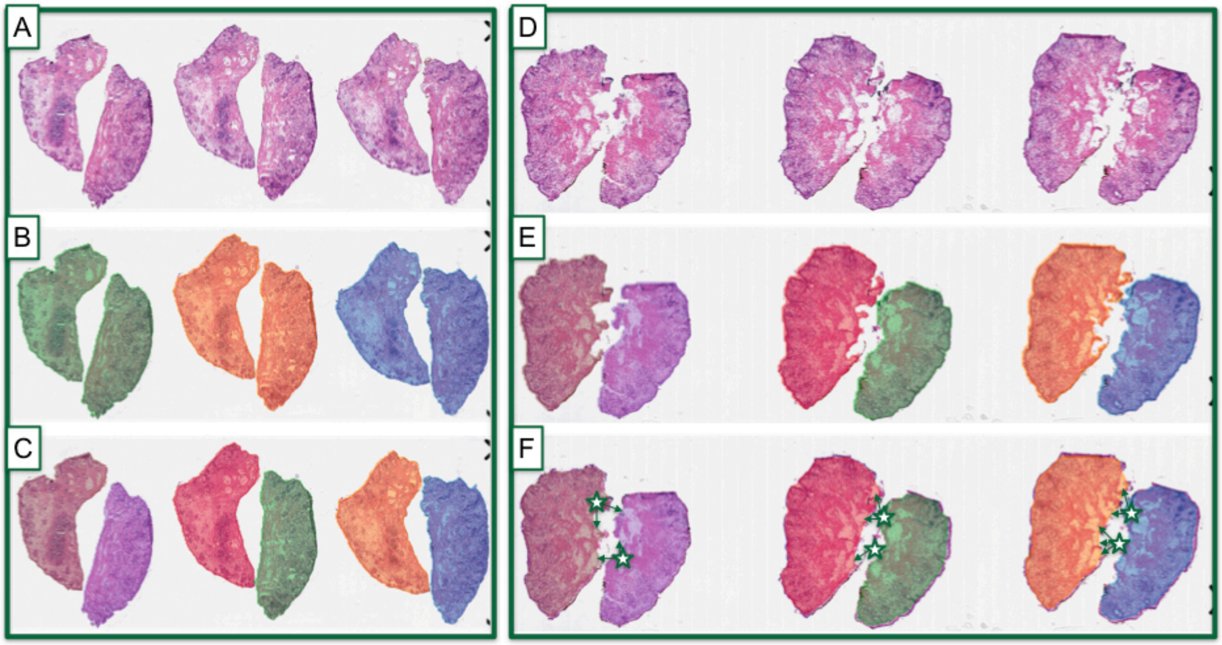


**Supplementary Figure 10: Histological assessment WSI preprocessing and separation of problematic conjoined sections for two cases:** A) original slide image (of three WSI) for first patient; B) tissue section assignment via 3 colors, where there are two conjoined pieces per section; C) separation of conjoined pieces for each section yields a unique piece identifier, each of which is separately processed via the histological assessment subworkflow; D) original slide image (of three WSI) for second patient; E) illustration of tissue patches selected for Tumor CNN-GNN algorithm; F) illustration of tissue patches selected for Completeness CNN-GNN algorithm, where arrows emanating from stars denote location of candidate tears which are normally missed but here are picked up through an alpha shape algorithm

### Supplementary Tables

#### Supplementary Table 1: Nuclei Prediction Workflow Accuracy

| Task | Model | Metric | Score | 2.5% CI | 97.5% CI |
|---|---|---|---|---|---|
| **Nuclei Detection** | **Detectron** | AP50 (0-1) | 0.224 | 0.220 | 0.227 |
| | | Dice Coefficient | 0.847 | 0.846 | 0.847 |
| **Nuclei Classification** | **Detectron** | F1-Score | 0.684 | 0.682 | 0.685 |
| | **Cell-CNN** | F1-Score | 0.775 | 0.774 | 0.776 |
| | **Cell-GNN** | F1-Score | 0.856 | 0.852 | 0.856 |

**Supplementary Files**

**Supplementary File 1: Video of 3D modeling workflow for three cases, A-C)**: tissue (left) are extracted frame-by-frame from turntable videos (top) and used to generate 3D models (right), where size is recorded and inking/sectioning recommendations are made

**Supplementary File 2: Illustration of surgical tumor mapping from histological findings in real-time:** A) viewing predicted tumor and inks from one tissue section via histology pane; B) histological findings and inks are mapped to circle via morphing model; C) real-time video of operating the mapping pane to place a tumor mapping area, from which the predicted tumor map is automatically plotted; D) predicted surgical tumor map; E) hand-drawn tumor map

**Supplementary File 3: Comparison of hand-drawn surgical tumor maps (left) to algorithm predicted tumor maps (right):** Expanded set of comparisons via PDF file for majority of test set cases
