## Supplementary Discussion for "ArcticAI: A Deep Learning Platform for Rapid and Accurate Histological Assessment of Intraoperative Tumor Margins"

**Supplementary Materials**

**Supplementary Discussion**

However, complete real time margin assessment is currently bottlenecked by tumor size and complexity, which impacts the application of this technique beyond the MMS setting. Technologies which optimize surgical care efficiency in this setting may ameliorate these constraints and are thus worthwhile investments. *ArcticAI* uses 3D reconstruction, graph neural networks, tissue mapping and workflow automation techniques to improve the efficiency and completeness of intraoperative surgical tumor resection to reduce the potential for costly repeat procedures and/or tumor recurrence. We expect this platform to improve communication between the surgeon, histotechnician and pathologist in the intraoperative setting, driving efficiency and completeness of intraoperative histological assessments. In turn, AI augmented real-time intraoperative margin assessments may yield significant fiscal savings for hospitals and reduce patient morbidity and mortality. Artificial intelligence has the potential to improve tumor removal through coupling the speed and accuracy neural networks with expert domain knowledge, which can increase the efficiency of healthcare delivery at multiple levels.

The *ArcticAI* platform is a margin analysis tool that addresses pragmatic surgical workflow considerations by targeting known bottlenecks of real-time surgical excision; from histological preprocessing to frozen section completeness, to tumor orientation and mapping. Furthermore, histological assessment results are on par with existing state-of-the-art results for BCC frozen sections. This is largely due to the incorporation of graph neural networks, which can integrate spatial information across the tissue, improving predictions while requiring less data. This platform is the first to employ graph neural networks for margin assessment on frozen tissue sections. Laboratory automation tools such as *ArcticAI* are well positioned to expand real-time margin analysis to more complex tumors given additional validation and testing while alleviating staffing issues by automating time consuming, low skill tasks.

*Future algorithmic improvements.* Given the myriad of workflow steps being optimized, the input parameters pertaining to the gross specimen (e.g., number of subsampled images, outlier point removal), tissue preprocessing (e.g., alpha shape construction) and histological assessment (e.g., learning rate) were coarsely assessed using sensible search grids to determine the optimal configuration. As such, we are planning to replace and improve some of the segmentation and reconstruction algorithms using deep learning approaches (e.g., low resolution tissue detection with detection network) which may circumvent tissue processing assumptions (such as background color threshold). The reconstruction process can also be sped up either by using a faster turntable (since only one-tenth of frames were utilized for this study) or through further subsampling of image frames (depending on the smartphone's capture speed). Gross specimen reconstruction can be further optimized using state-of-the-art multi-view stereo frameworks (MVS) such as Neural Radiance Fields (NERF), which can display the gross specimen at any input orientation. The image features learned through the CNN-GNN approach may be further refined through state-of-the-art deep learning models such as Vision Transformers, training methods such as self-supervision, and through pretraining the model (i.e., initializing or transferring the knowledge) based on training a model on similar frozen histological specimens.

The completeness assessment algorithm did remarkably well in separating holes/tears from other similar structures, such as fat, wispy dermis, and gaps introduced from large follicles, though there were some instances where the algorithm had erroneously predicted these structures. We also noticed instances where the presence of follicles and sebaceous/eccrine glands may have confounded tumor detection and completeness assessment respectively. But, for the most part, hair follicles were not conflated with tumor prediction since the AUC remained unchanged after accounting for follicles and through qualitative assessment (e.g., follicular structures in **Figures 4-5**). While we attempted to control for follicles as a confounder through detection and downweighting of patches containing follicles, we acknowledge that this approach may be further improved in follow up works which allow for simultaneous prediction of follicle and tumor without conflating the two.

Prediction of tumor was highly accurate, much of which can be attributed to the inclusion of inflammatory foci in the prediction algorithm as a separate class. This allowed for inflammation to be detected and excluded from tumor consideration. However, this left open the very real possibility of tumor cells surrounded by inflammation failing to be detected. So, as an added layer of security, we prototyped a neural network algorithm to predict nuclei within pockets of inflammation to rule out tumor, which was annotated for both inflammation and tumor. However, there does not yet exist large datasets of millions of nuclei across many slides for nuclei prediction across frozen sections, as compared to permanent sections for specific cell types (e.g., follicle nuclei, basal cell, epidermal keratinocytes, malignant, dermal fibroblasts, etc.). Detecting and separating individual benign from malignant cells may require millions of tedious cell annotations to achieve the consistency required for clinical use. We plan to investigate methods which can rapidly expand our annotation set with minimal cost: 1) expert-in-the-loop, where neural network generates annotations for pathologists to accept or reject [1]; 2) developing annotation efficient methods (e.g. point annotations) instead of complete nuclear contours or are weakly supervised or self-supervised [2,3]; 3) virtual immunohistochemical staining, which can be used to label malignant cells[4], and 4) other data efficient approaches whereby certain types of annotations may no longer be required to improve the test set prediction. Finally, the web application may benefit from additional mobile application development for maximal usability.

*External applicability.* While two dermatopathologists curated annotations for this dataset, expert annotation is not always guaranteed to agree and can potentially serve as a source of measurement error/uncertainty which may change the accuracy of the model results. Avoiding issues of measurement variability/uncertainty may potentially distort study outcomes. In future studies, we will recruit a team of expert annotators and measure interrater reliability to ascertain the degree of certainty for tumor mapping. We also acknowledge that data collection was limited to a single site and an MMS laboratory and future studies are required to create and test additional external cohorts. For instance, the study cohort is predominantly comprised of older Caucasian males based on demographics of the service region. While in line with the known epidemiology surrounding disease risk groups [5,6], it is essential to consider other diverse groups to ensure external applicability and to control for such variables as race and sex. Differences in sectioning quality, staining, and imaging may also pose additional sources of variability and necessitate a multi-center clinical trial. Finally, while the role of the Mohs surgeon may vary depending on the hospital or health care system (e.g., surgeon also performs histological assessment, overlapping expertise and workspaces), real time intraoperative settings for excision of more complex tumor types may more clearly delineate these roles.

*Next steps.* Pending additional algorithmic finetuning, real-world testing through clinical trials, and assessments of adoptability, there are ample opportunities to apply this technology to studying additional solid tumor types. In skin, Squamous Cell Carcinoma and Melanoma present opportunities to further evaluate the proposed workflow, though each would require additional data collection and algorithmic finetuning. Melanoma is particularly challenging as ancillary immunohistochemistry is often required for identification of positive margins. Other more challenging solid tumor types include head and neck, breast, lung, colon, and urinary bladder cancers, which require more significant changes to the surgical workflow to test and implement real-time margin assessment technologies [7,8]. Processing time will vary by tissue size and number of sections, though using an AI-augmented surgical workflow, it is expected that there will be significant reductions in margin assessment time while maintaining or improving accuracy and completeness.
