## Supplementary File 3 for "ArcticAI: A Deep Learning Platform for Rapid and Accurate Histological Assessment of Intraoperative Tumor Margins"

### Hand Drawn and Predicted Surgical Tumor Maps

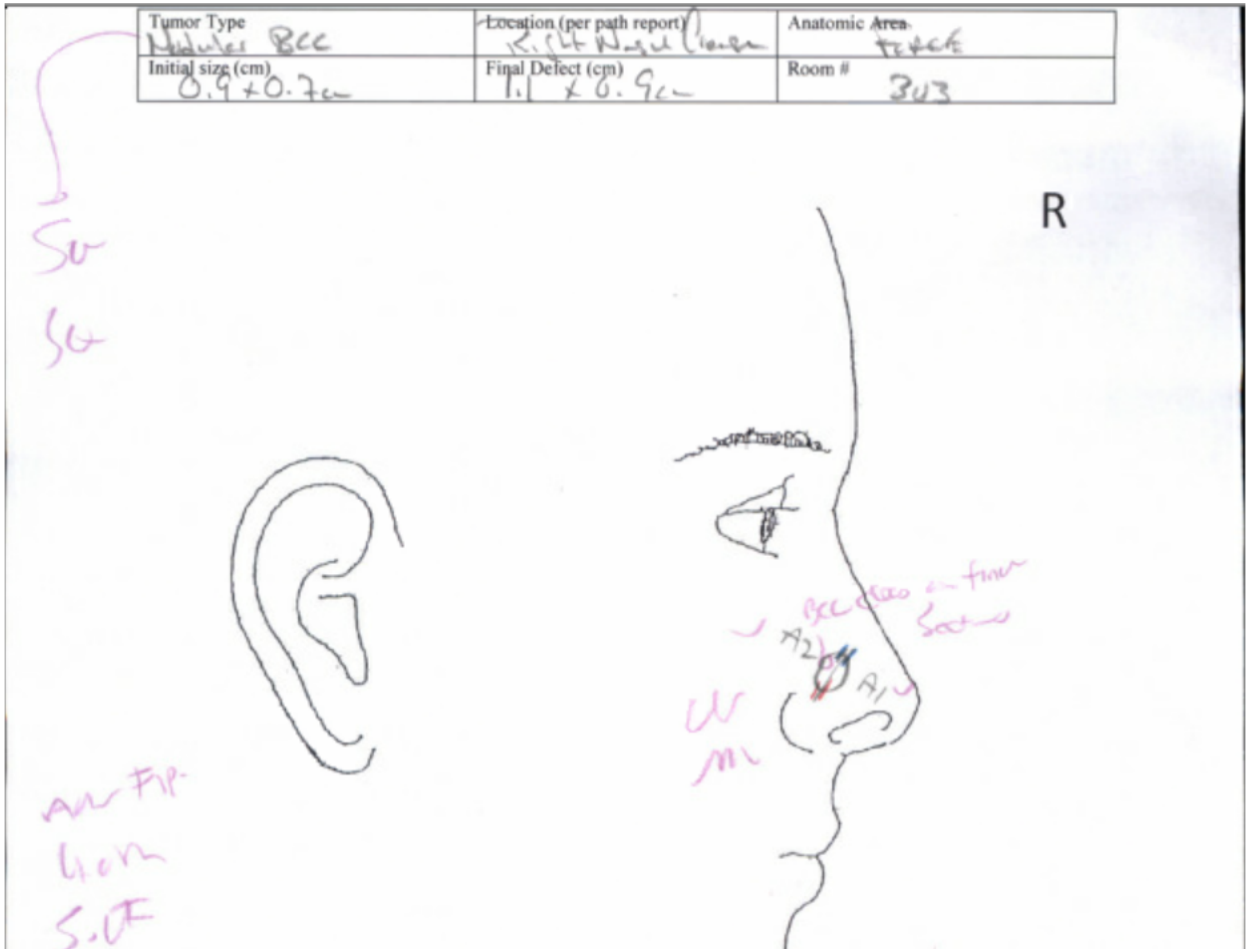


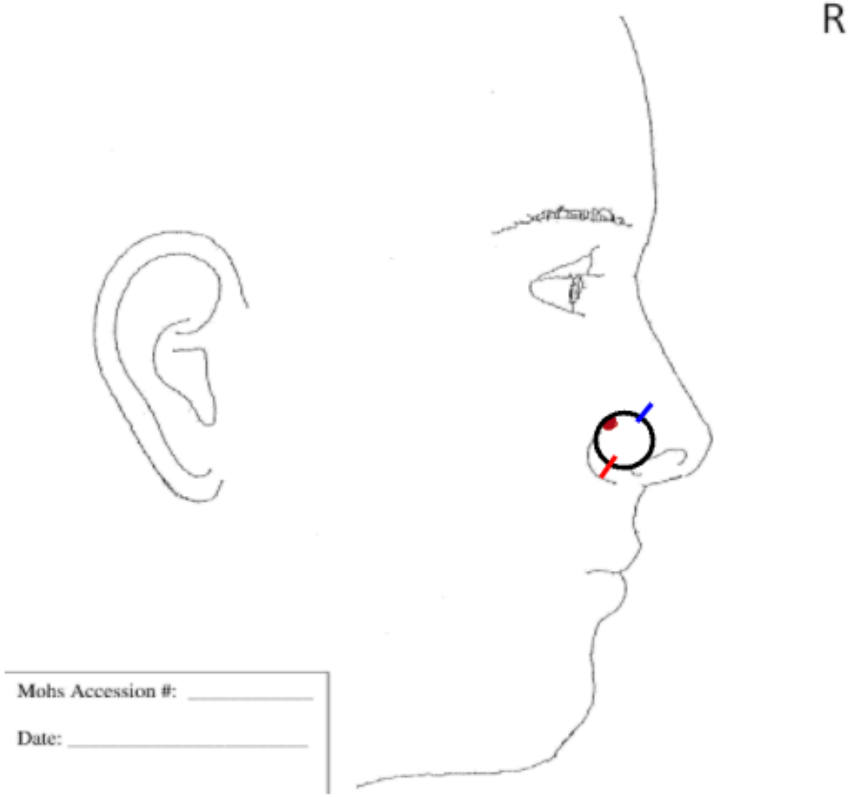

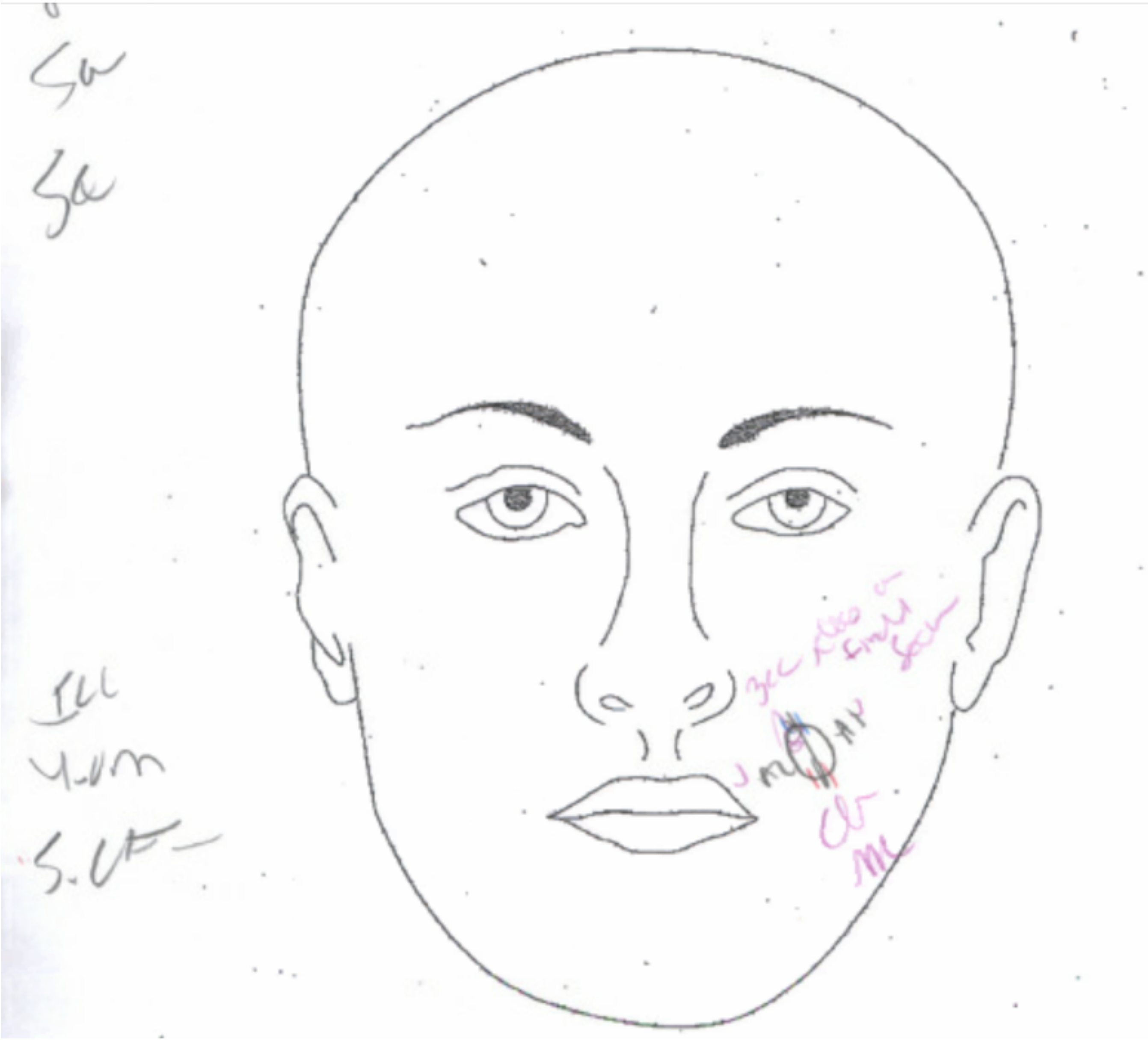

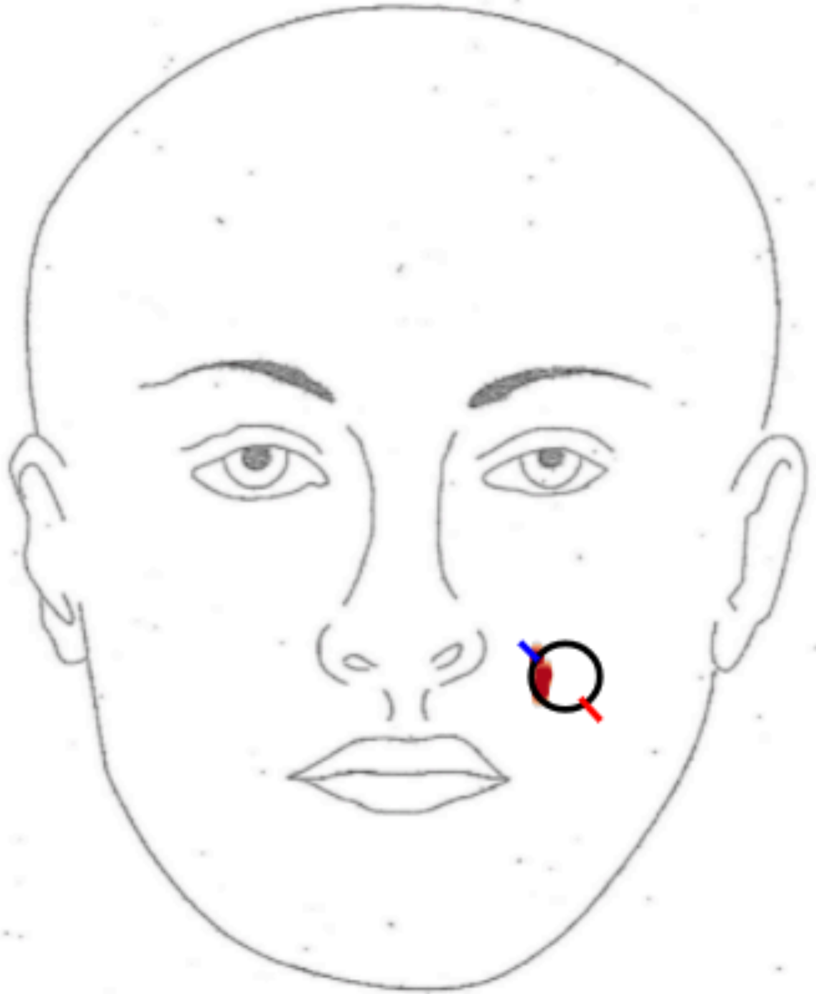

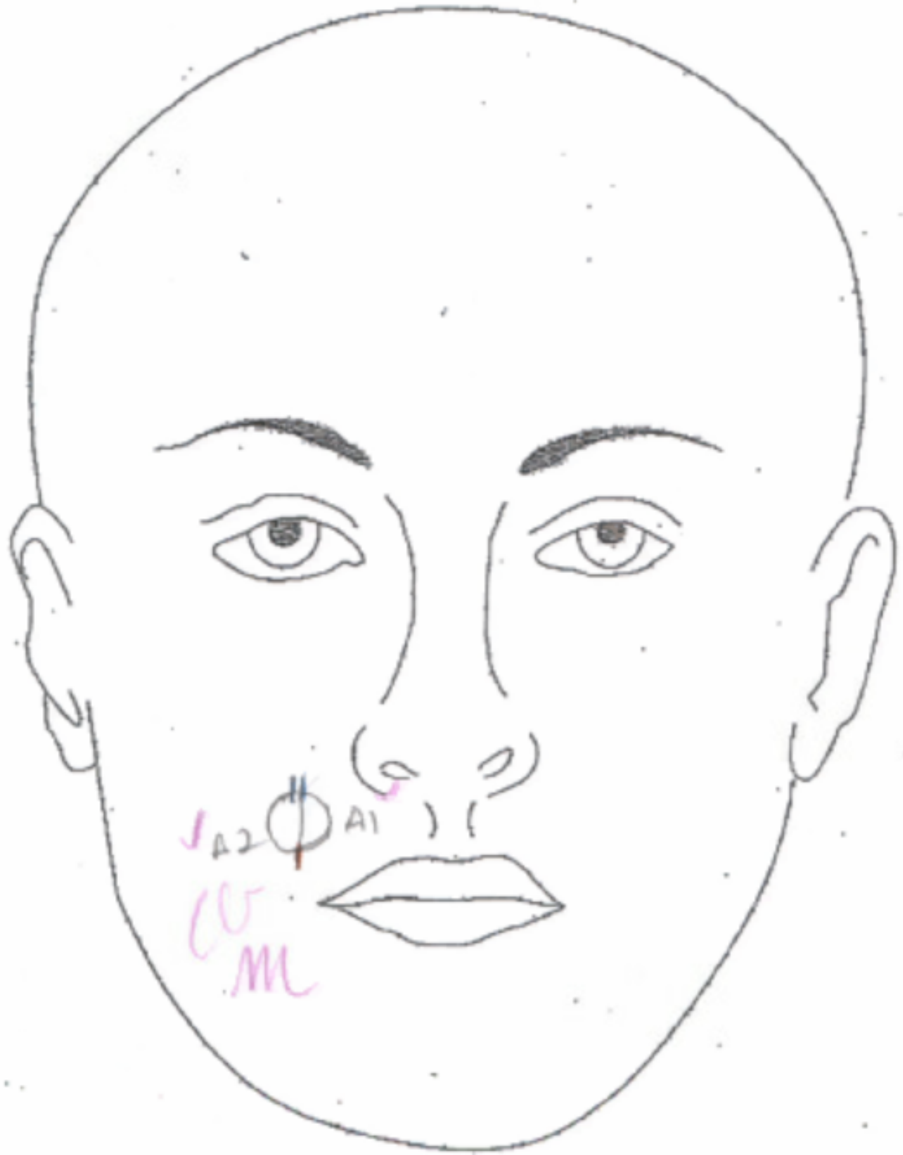
A2
A1

| Initial size (cm) | Final Defect (cm) | Room # |
| --- | --- | --- |
| 0.8 X 0.7 cm | 2.7 x 1.1 cm | 306 |

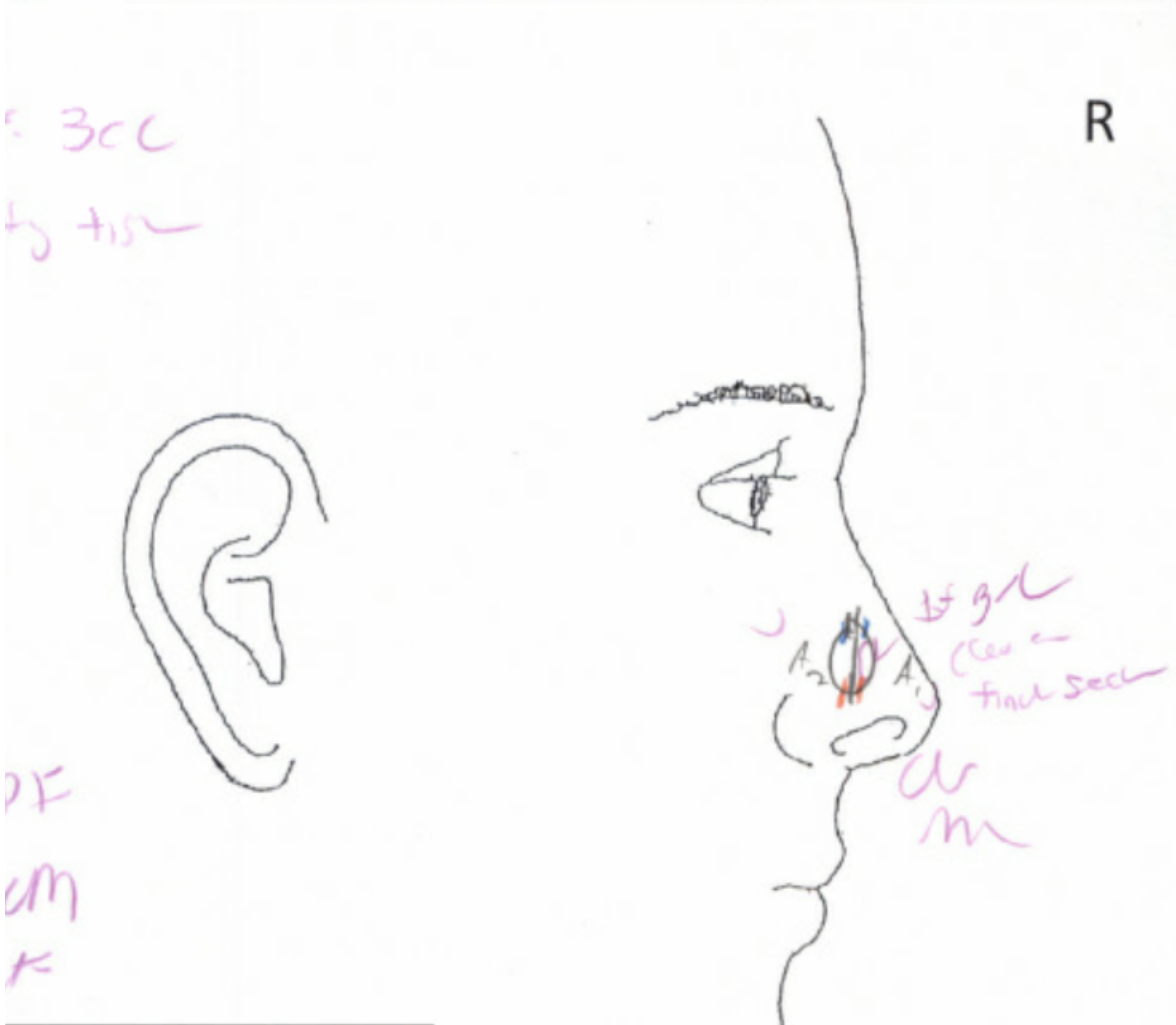


Mohs Accession #:

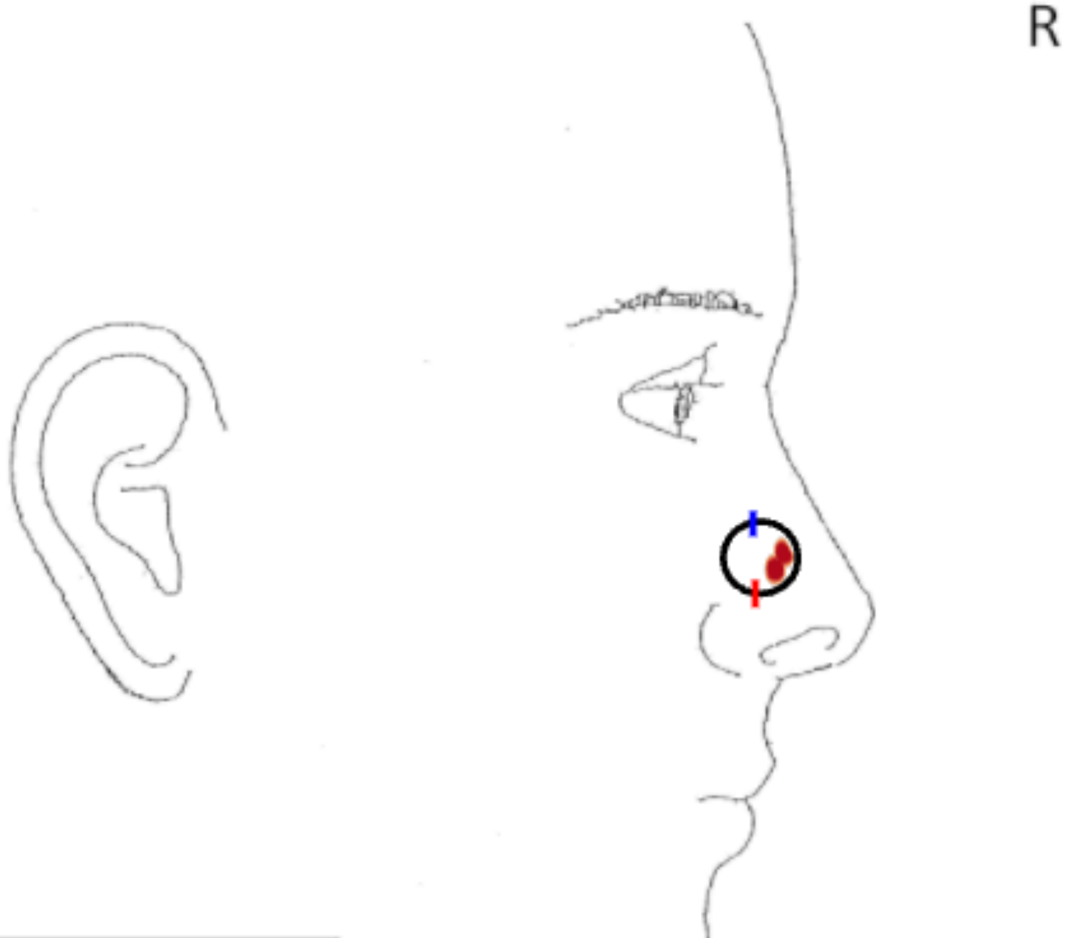


Mohs Accession #: ____________

Date: ____________

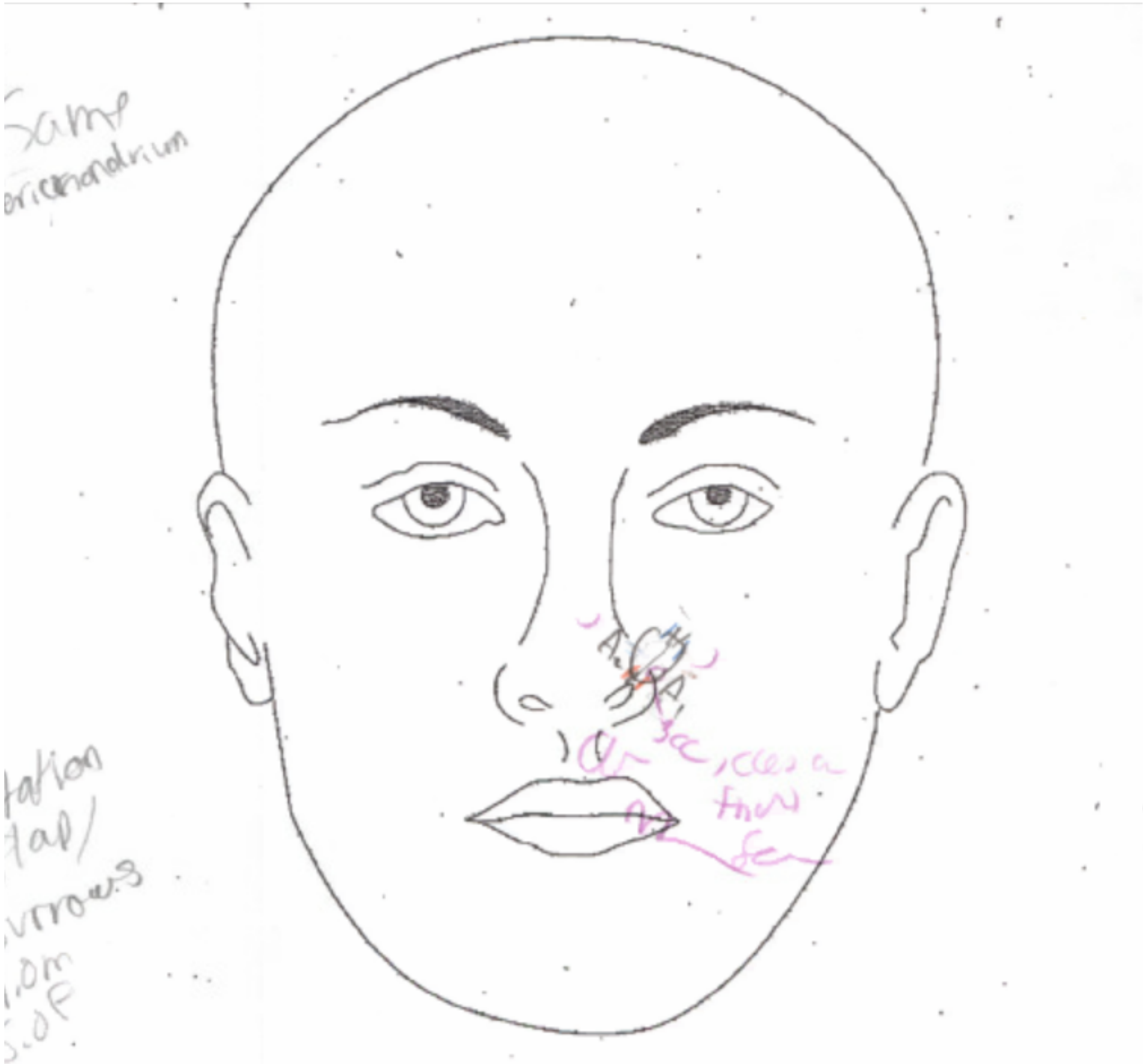

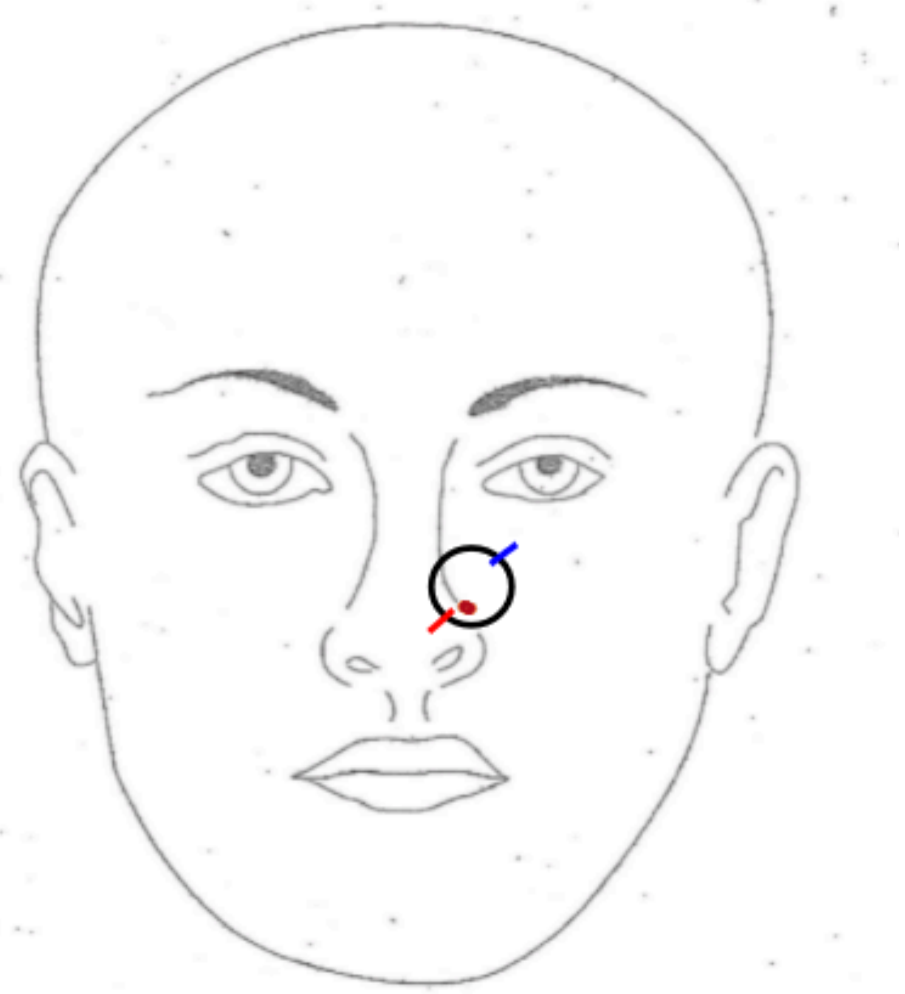

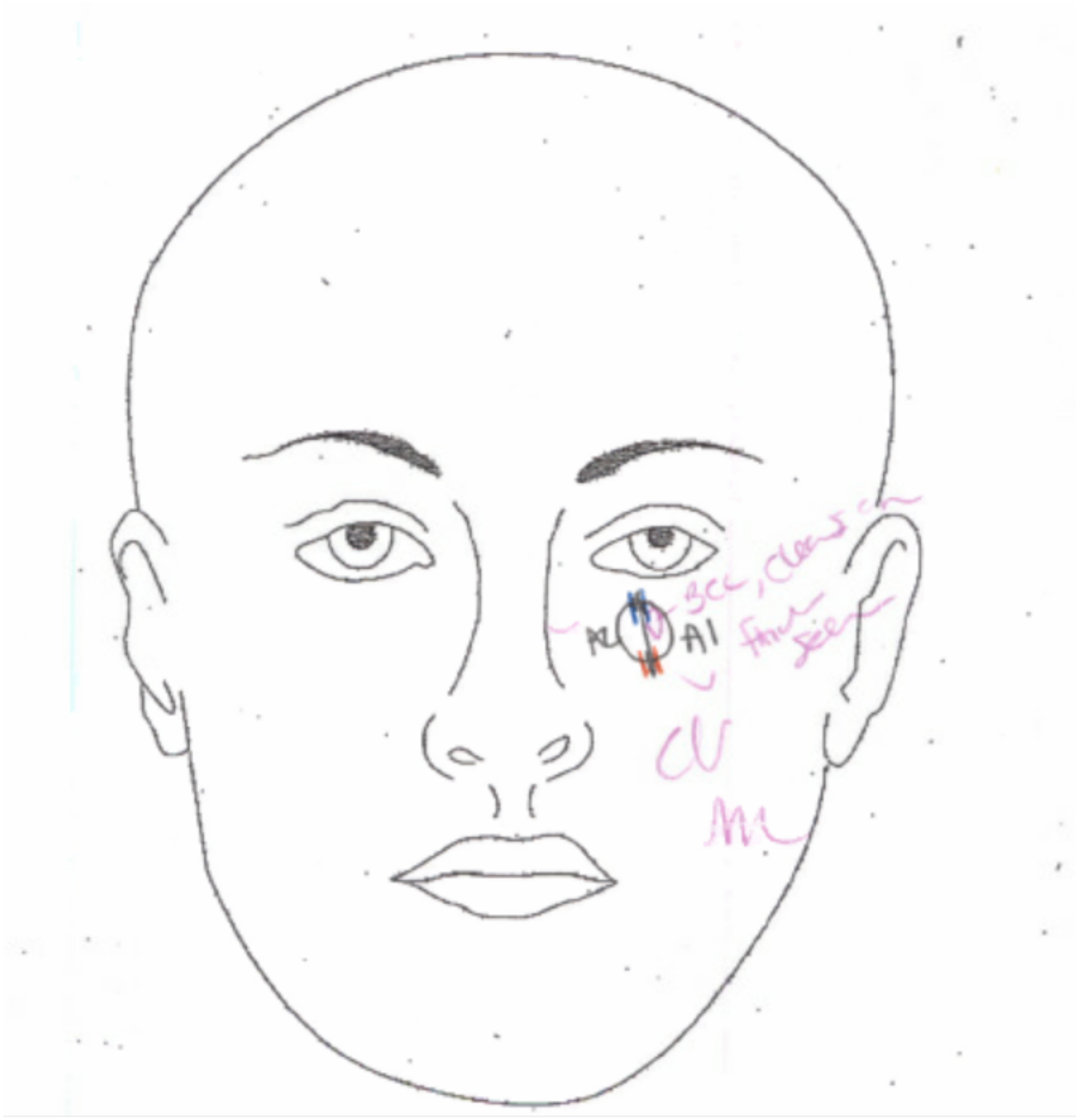

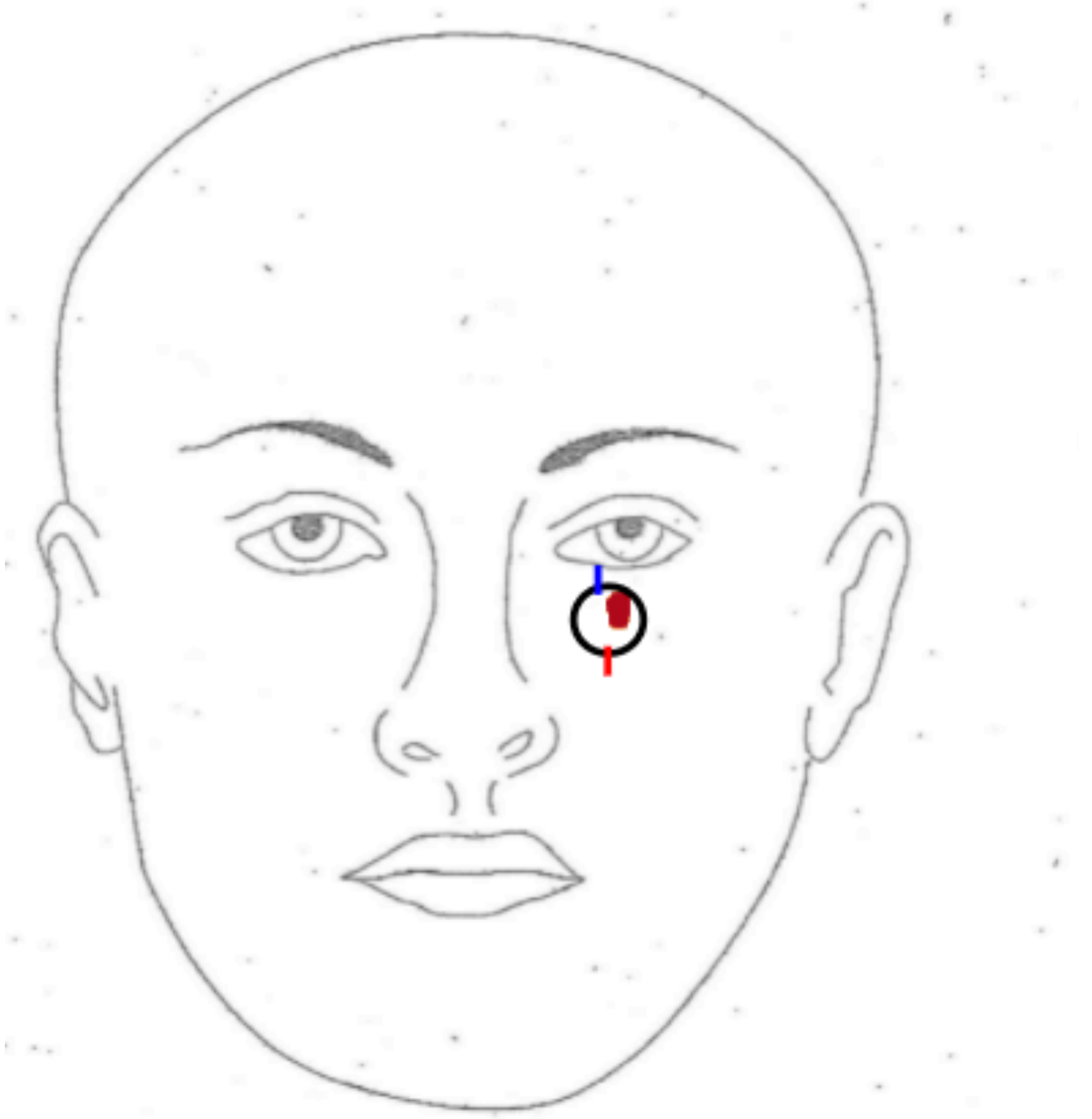

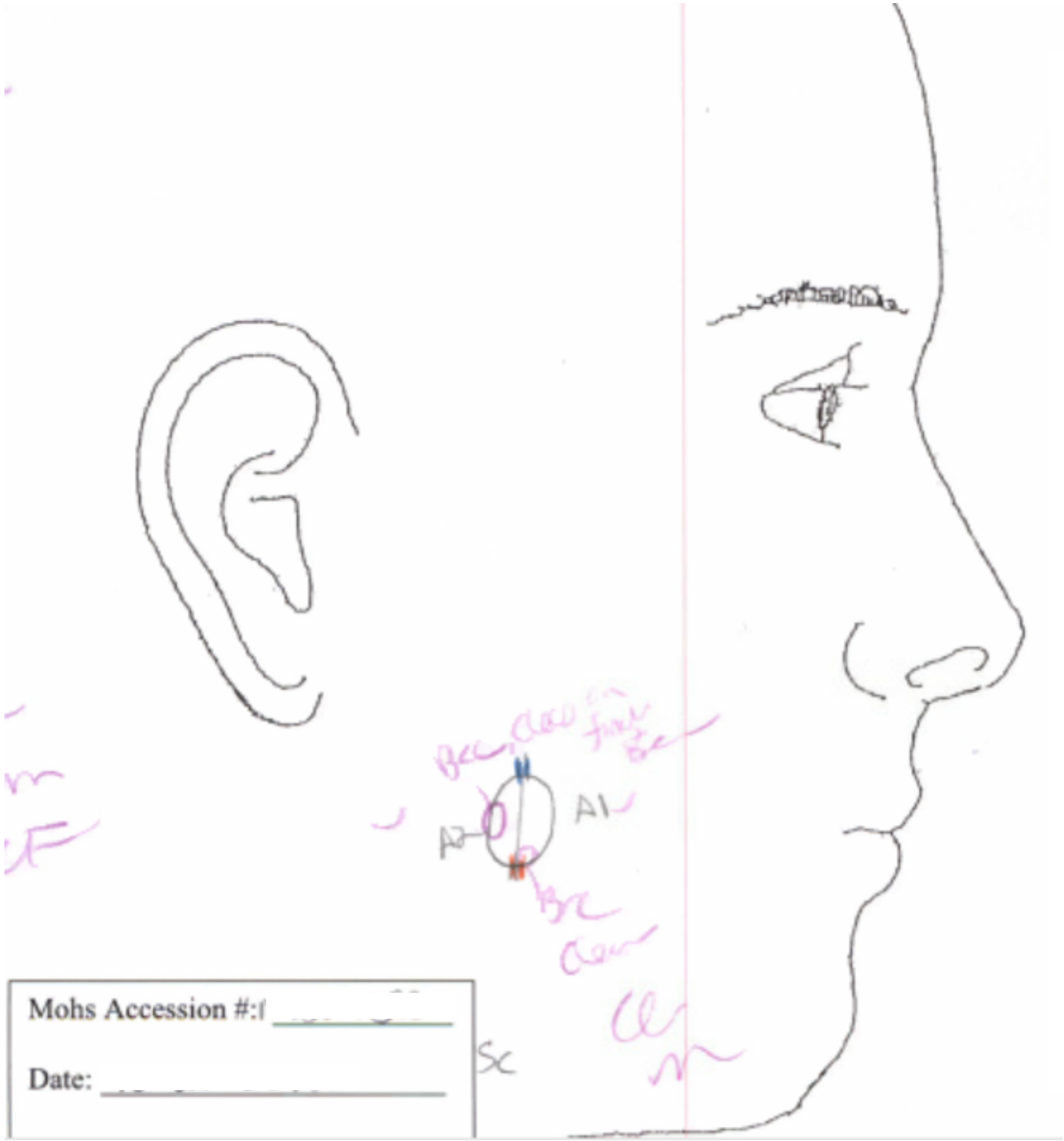
A1
A2
Mohs Accession #:
Date:

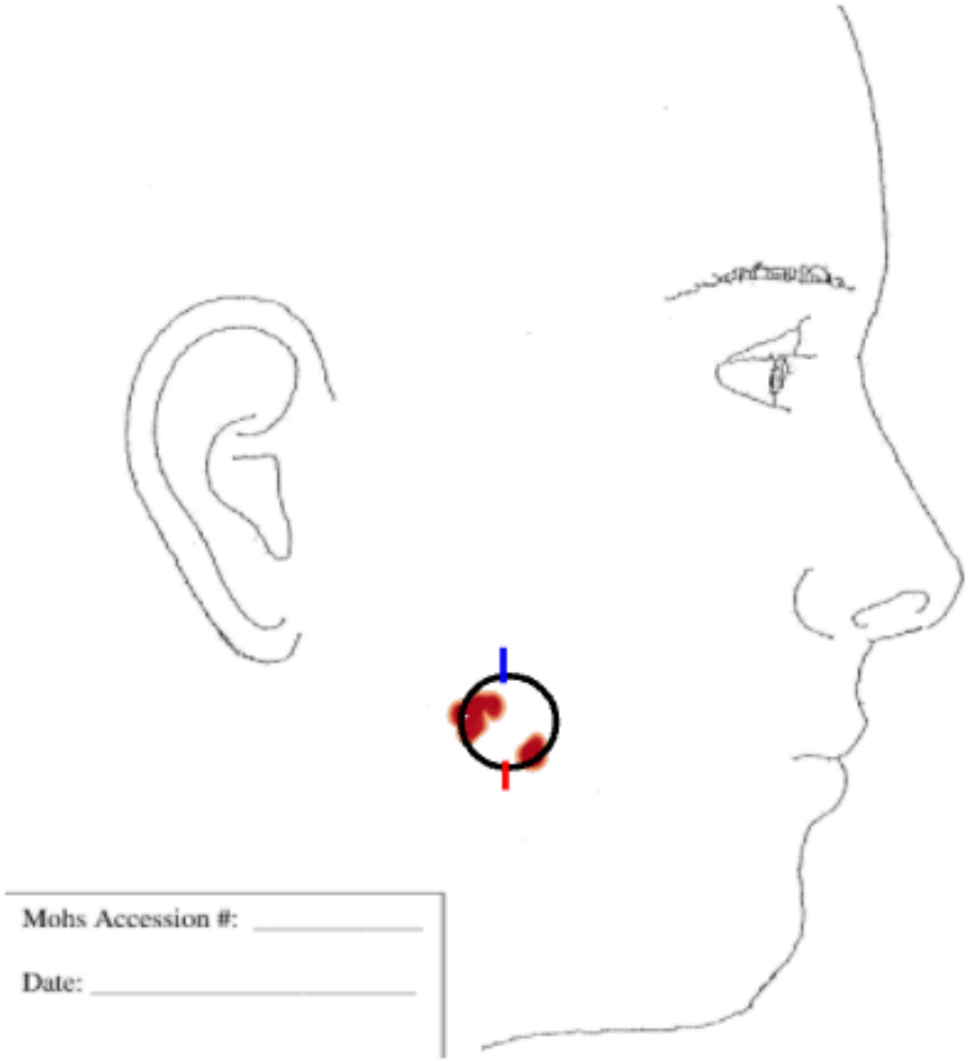
Mohs Accession #:
Date:

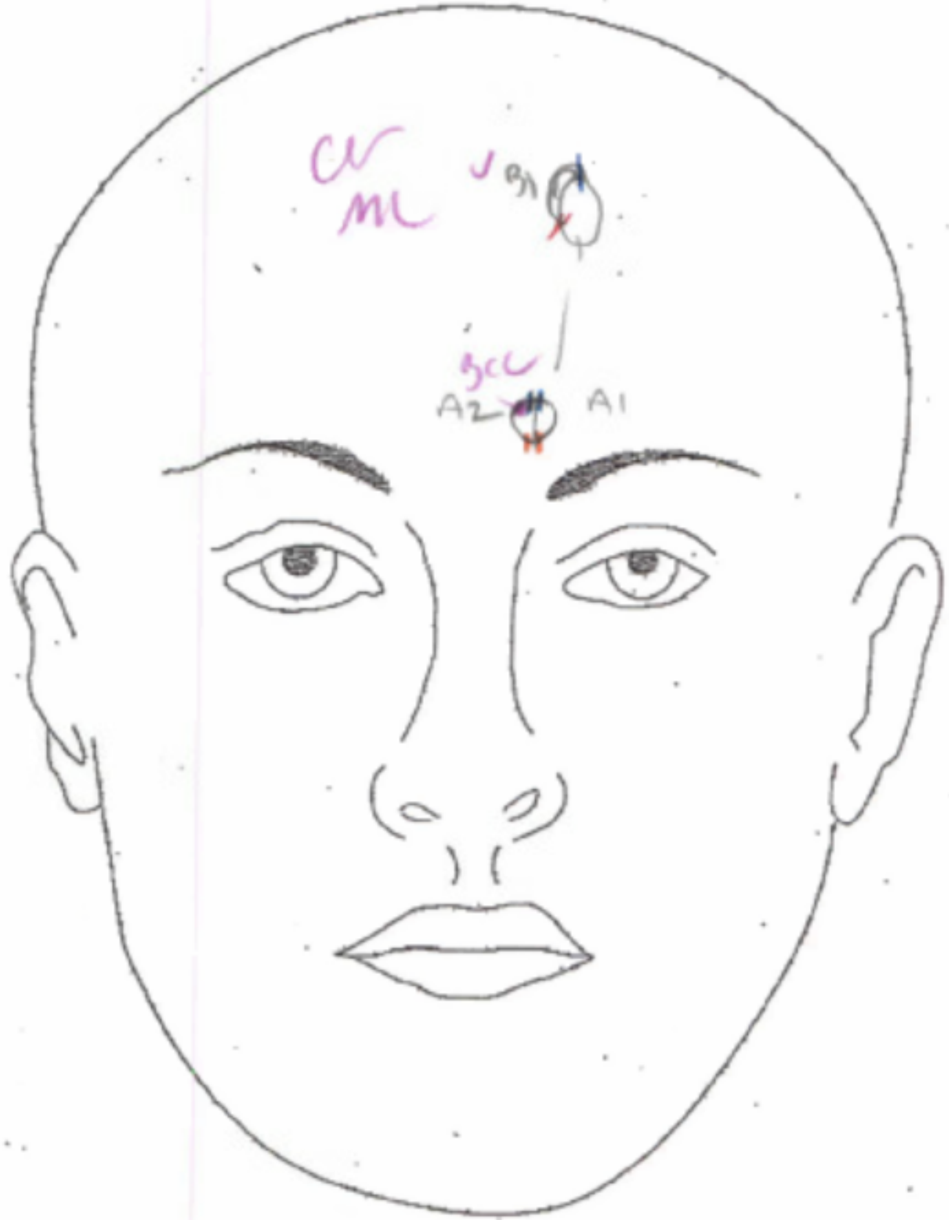
A2
A1

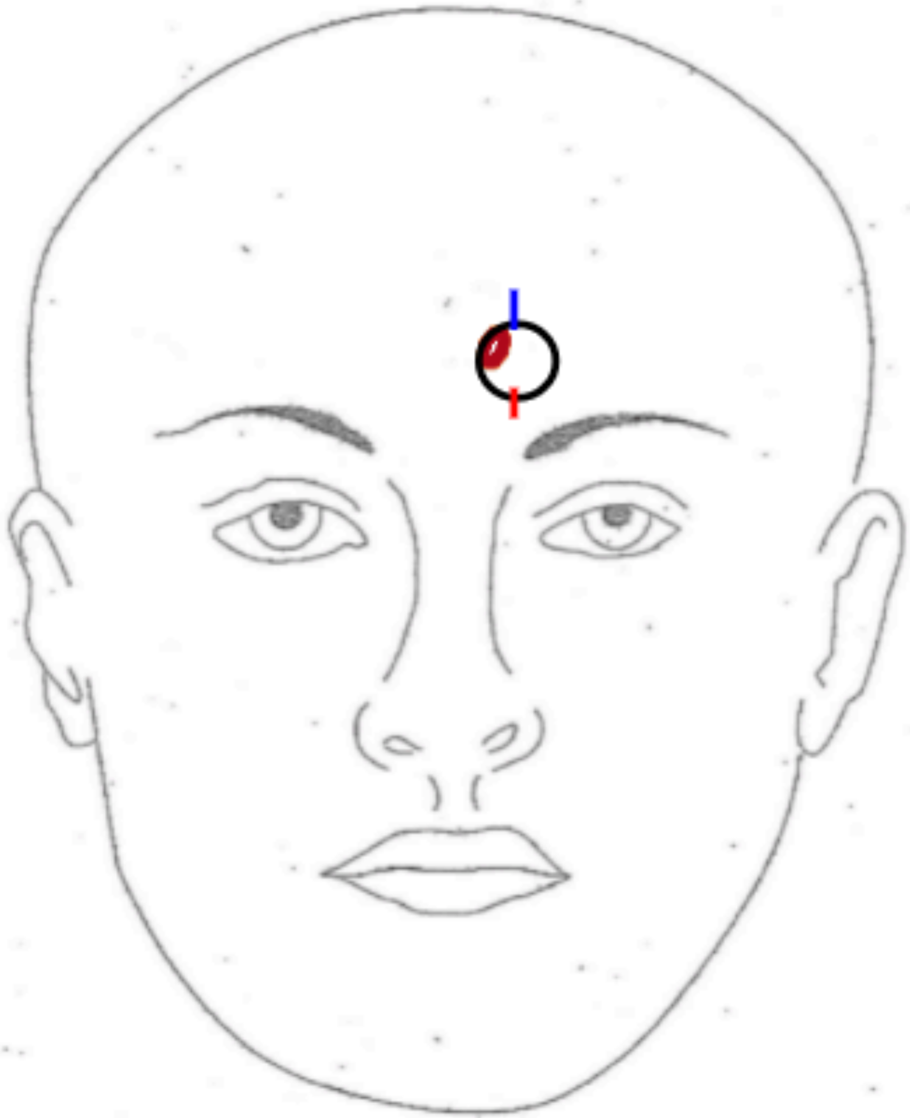

| Initial size (cm) | Final Defect (cm) | Room # |
| --- | --- | --- |
| 1.0x1.3 cm | 1.8x1.5 cm | |

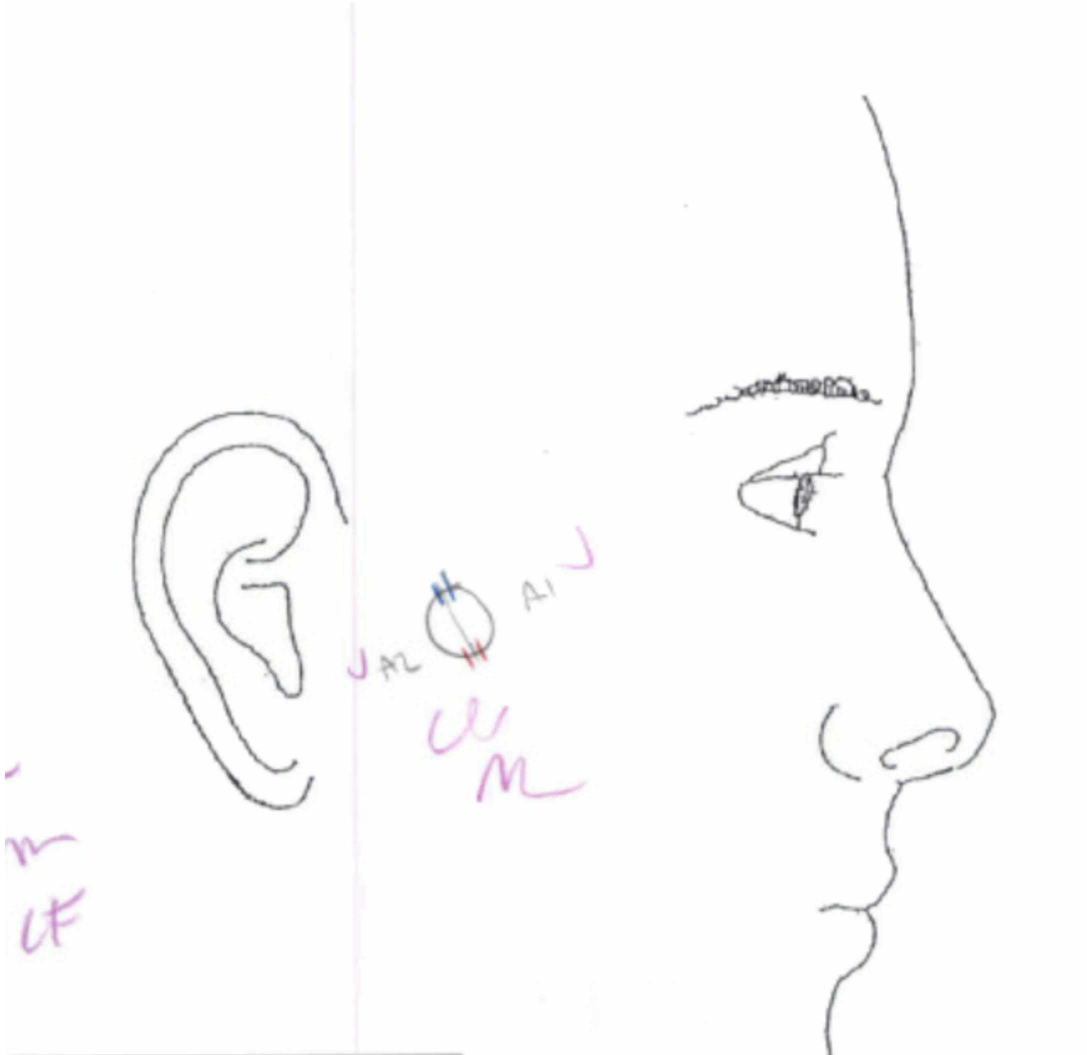


Mohs Accession #: ____________

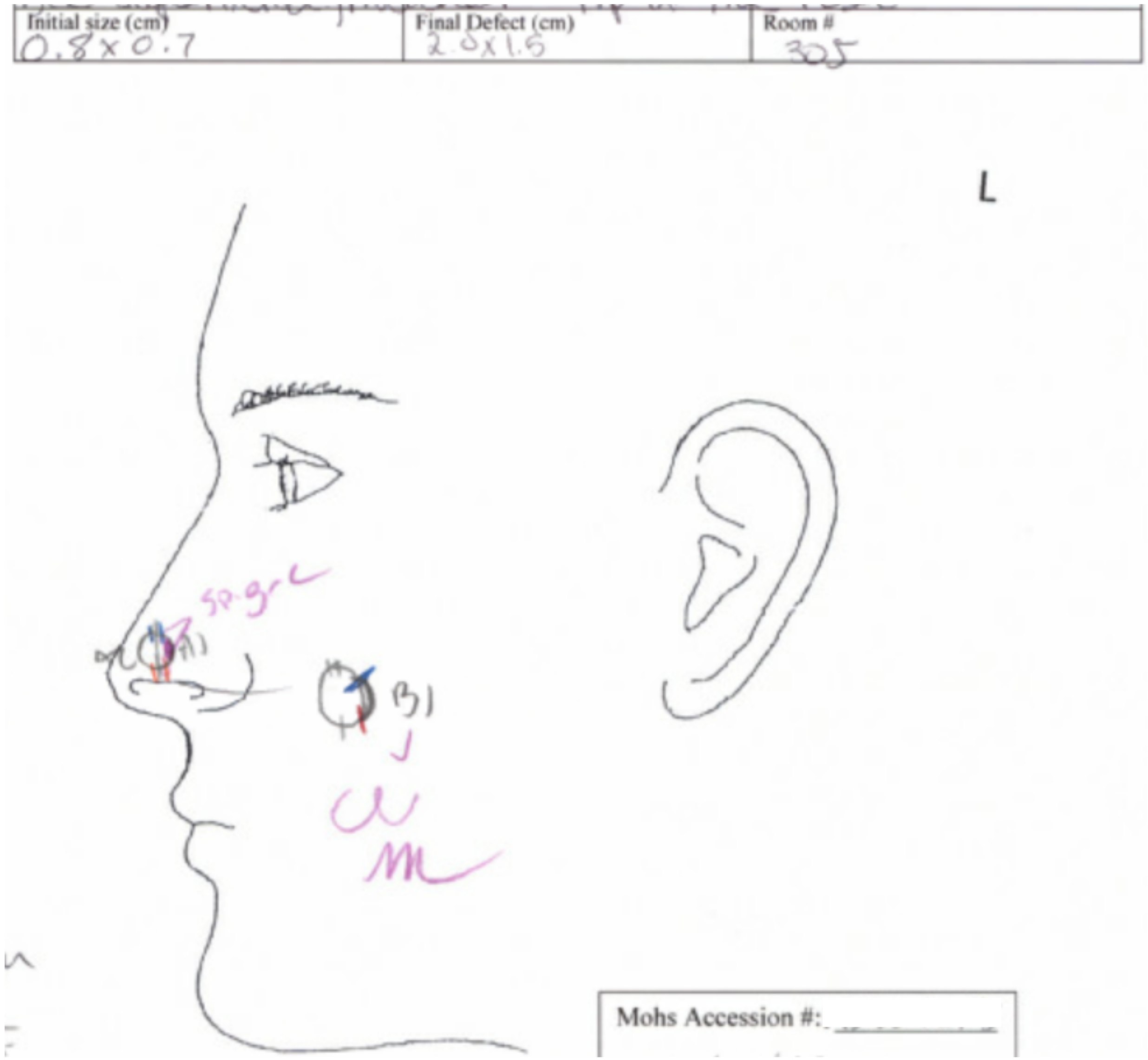
Initial size (cm)
0.8x0.7
Final Defect (cm)
2.0x1.5
Room #
305
L
A1
B1
Mohs Accession #:

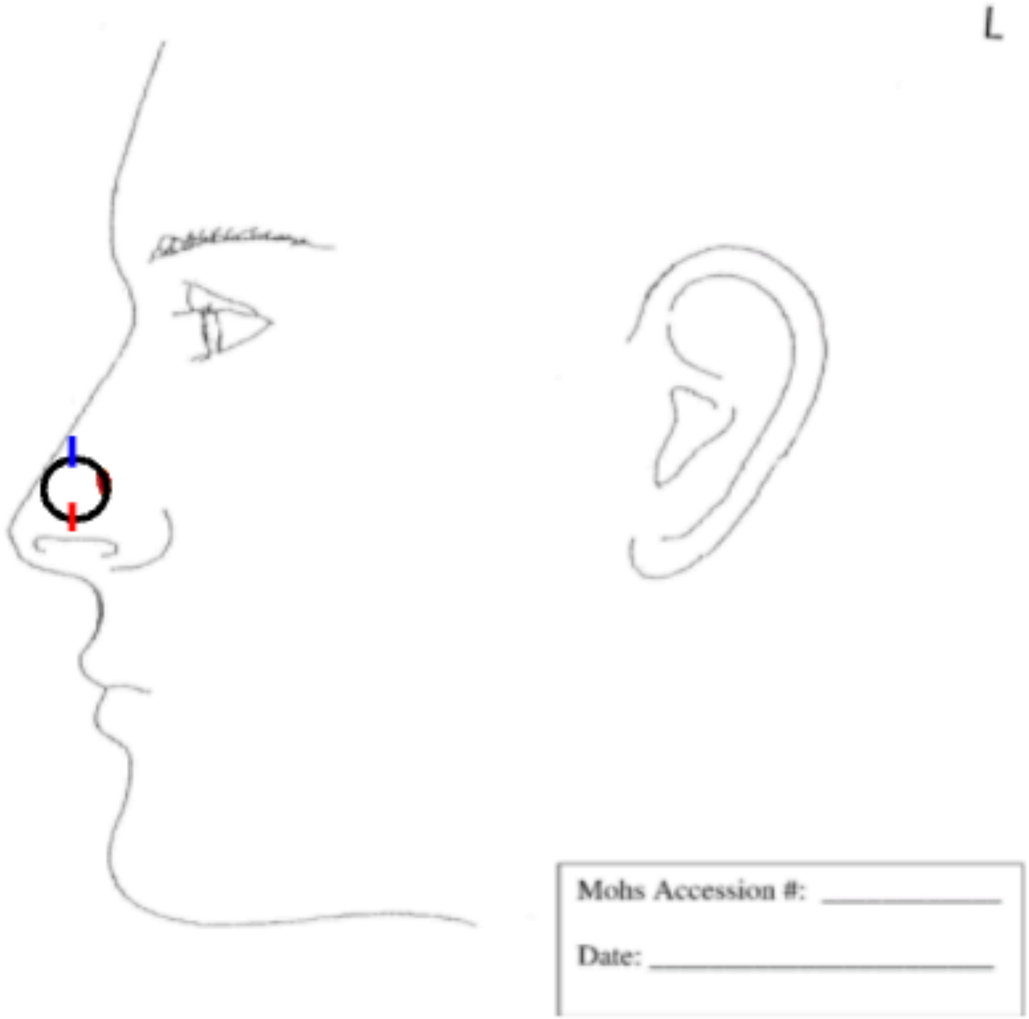
L
Mohs Accession #:
Date:

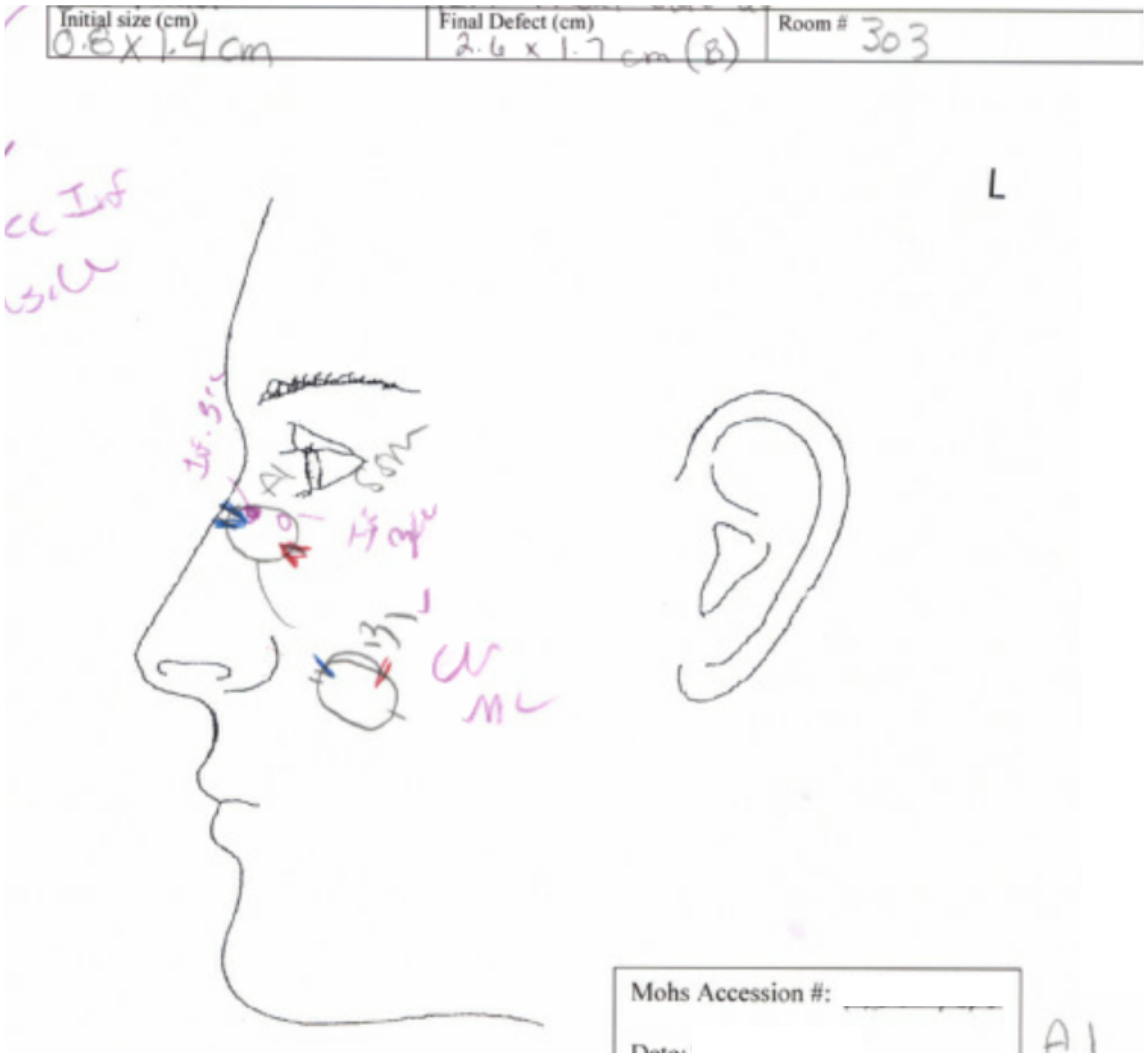
Initial size (cm)
0.8 x 1.4 cm
Final Defect (cm)
2.6 x 1.7 cm (B)
Room # 303
L
Mohs Accession #:

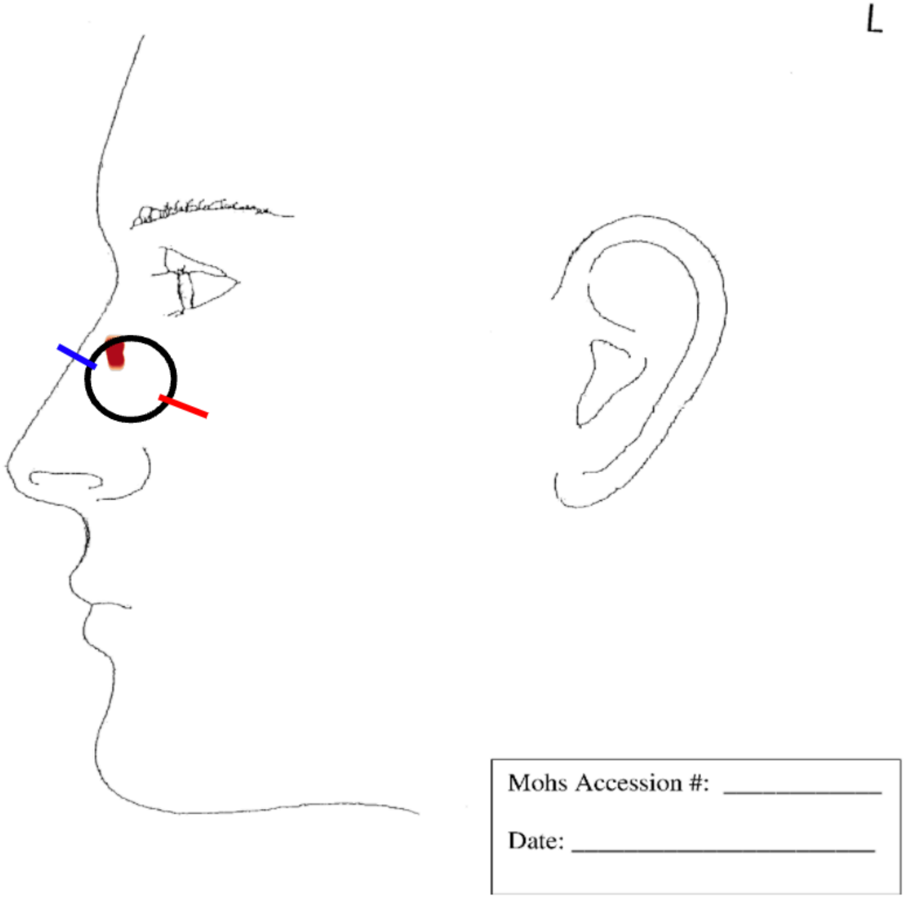
L
Mohs Accession #:
Date:

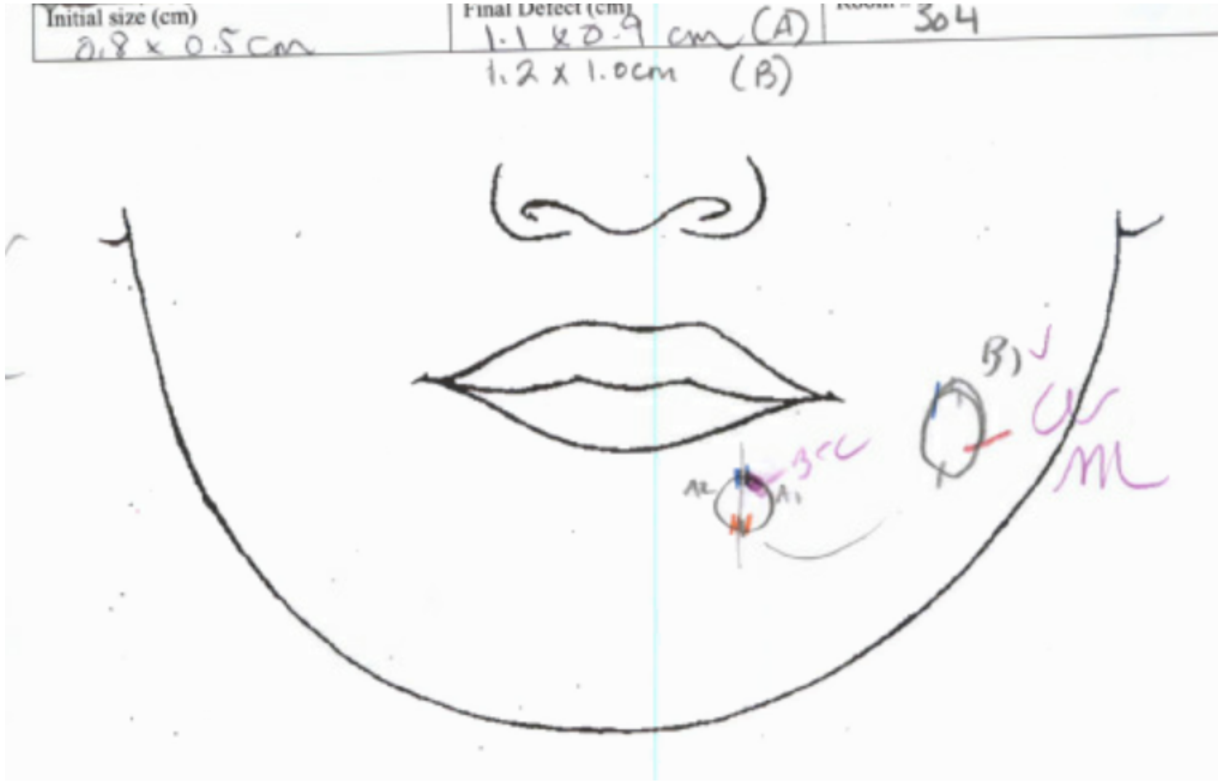
Initial size (cm)
0.8 x 0.5 cm
Final Defect (cm)
1.1 x 0.9 cm (A)
1.2 x 1.0cm (B)
304

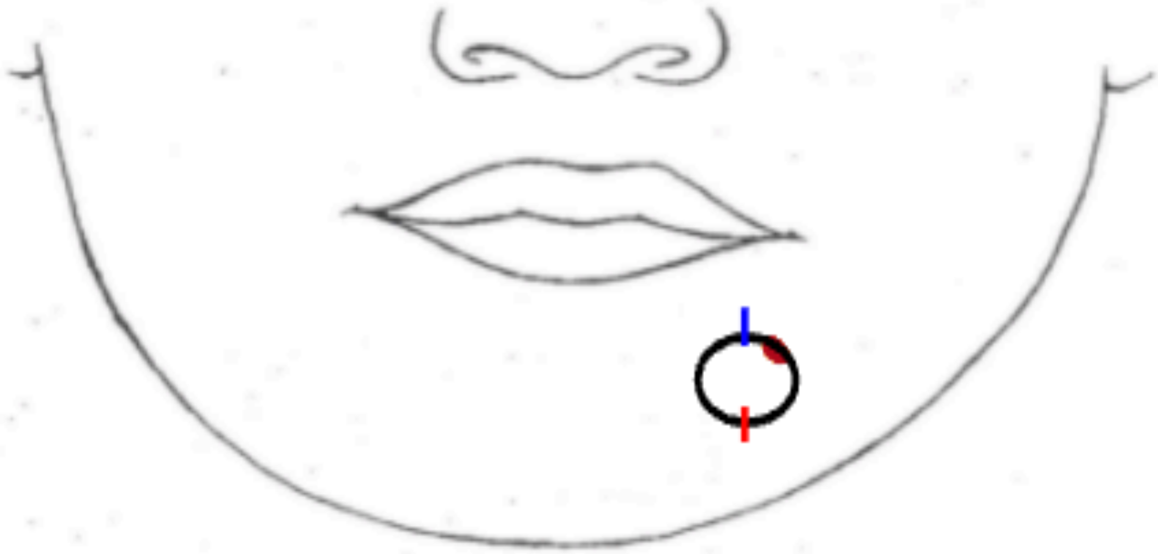

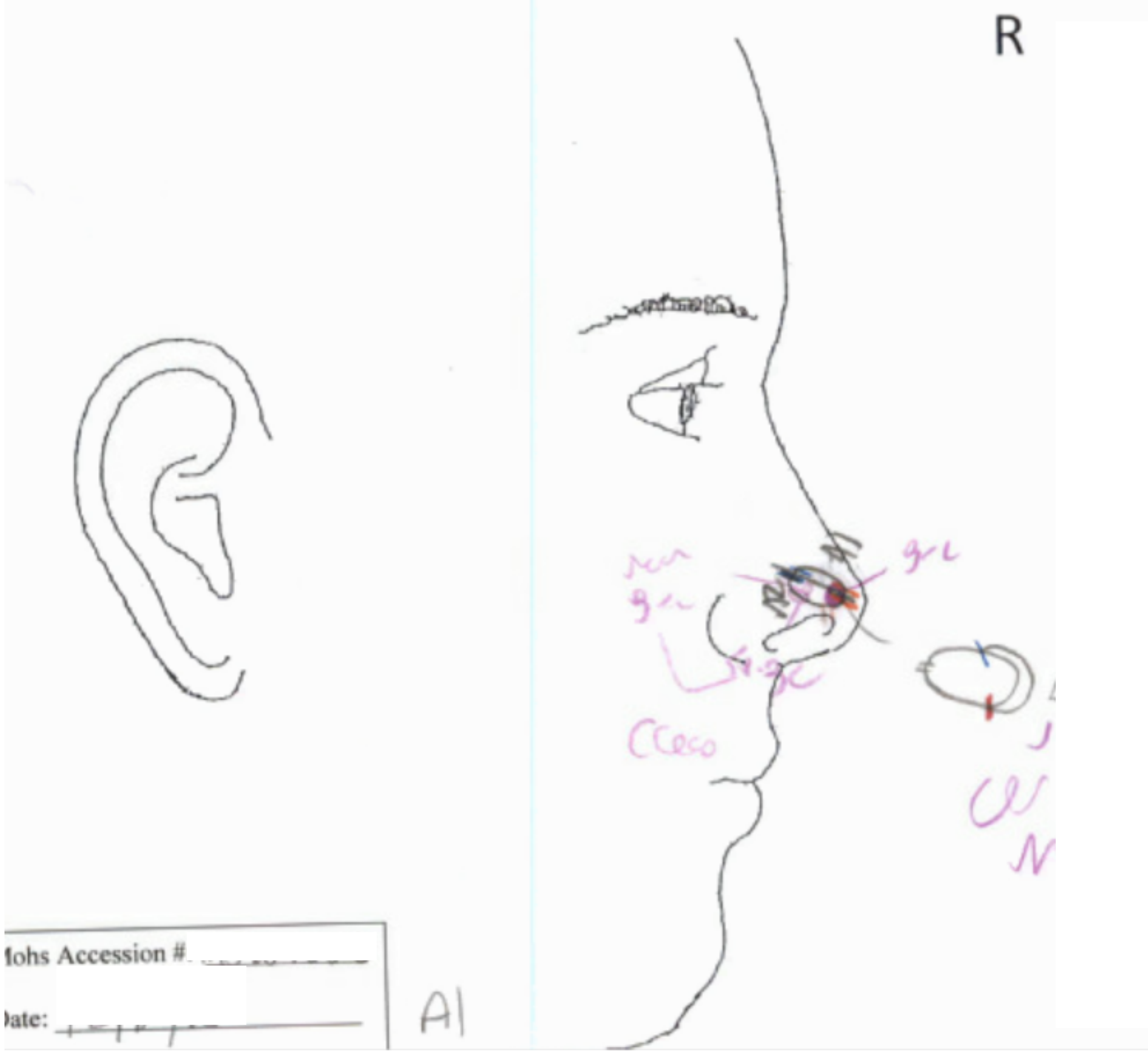
R
Mohs Accession #
Date:
A1

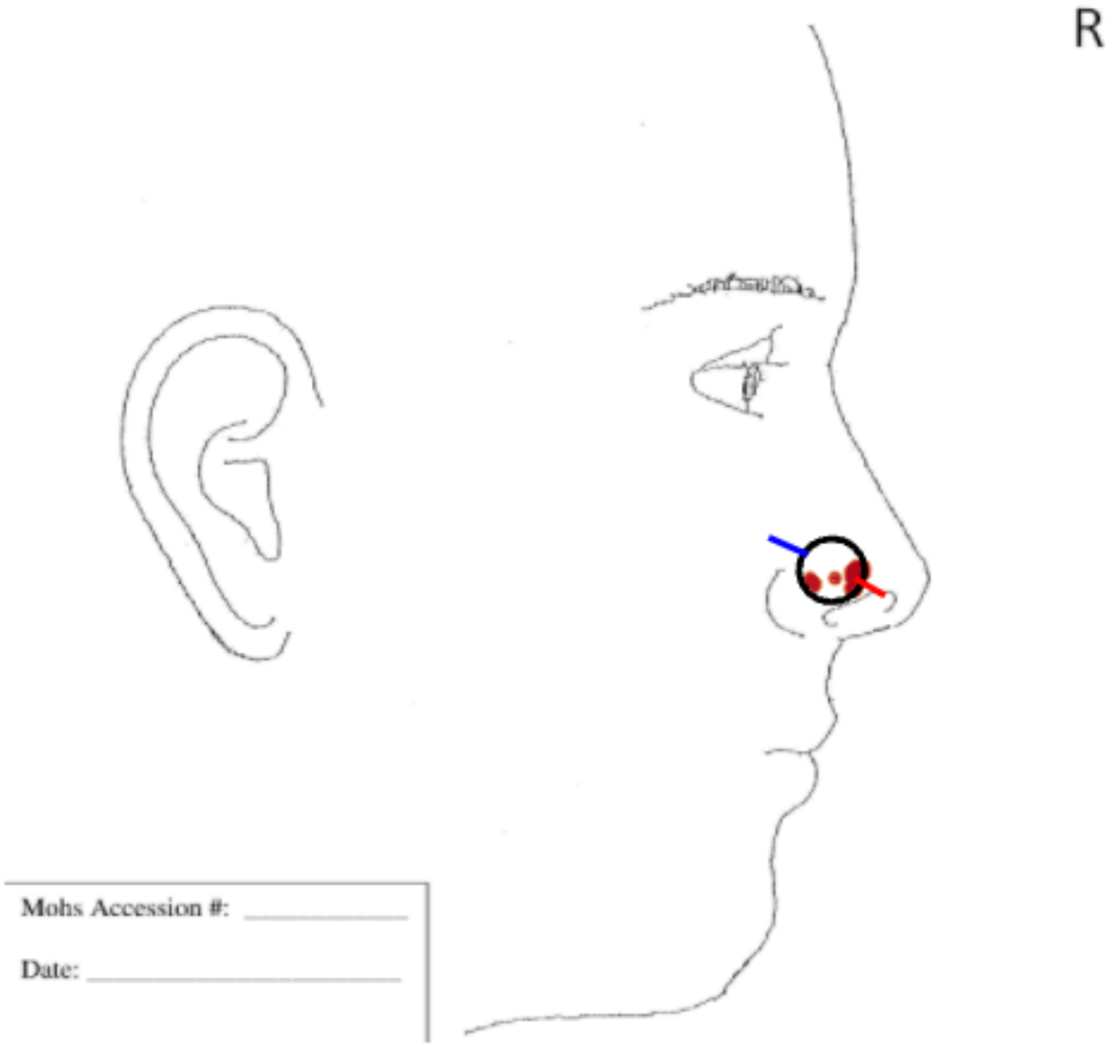
R
Mohs Accession #:
Date:

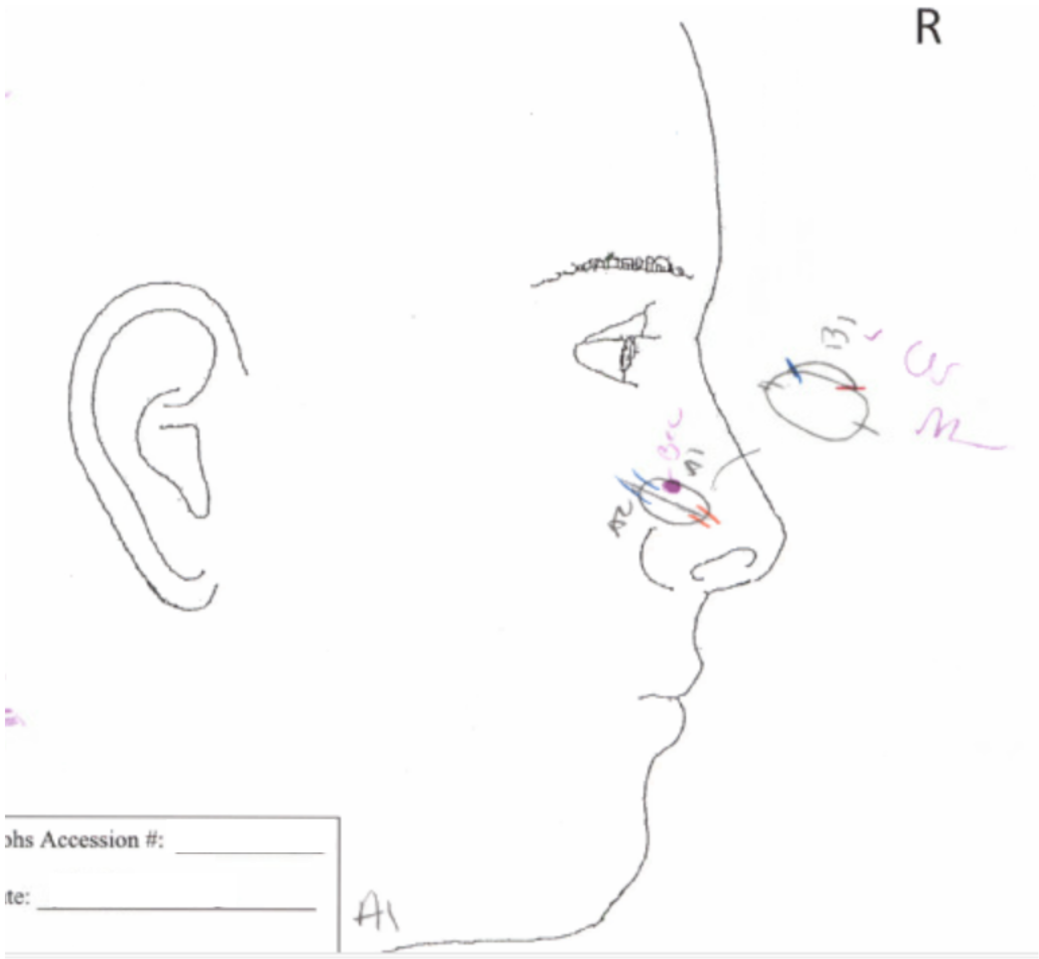
R
hs Accession #:
te:
A1

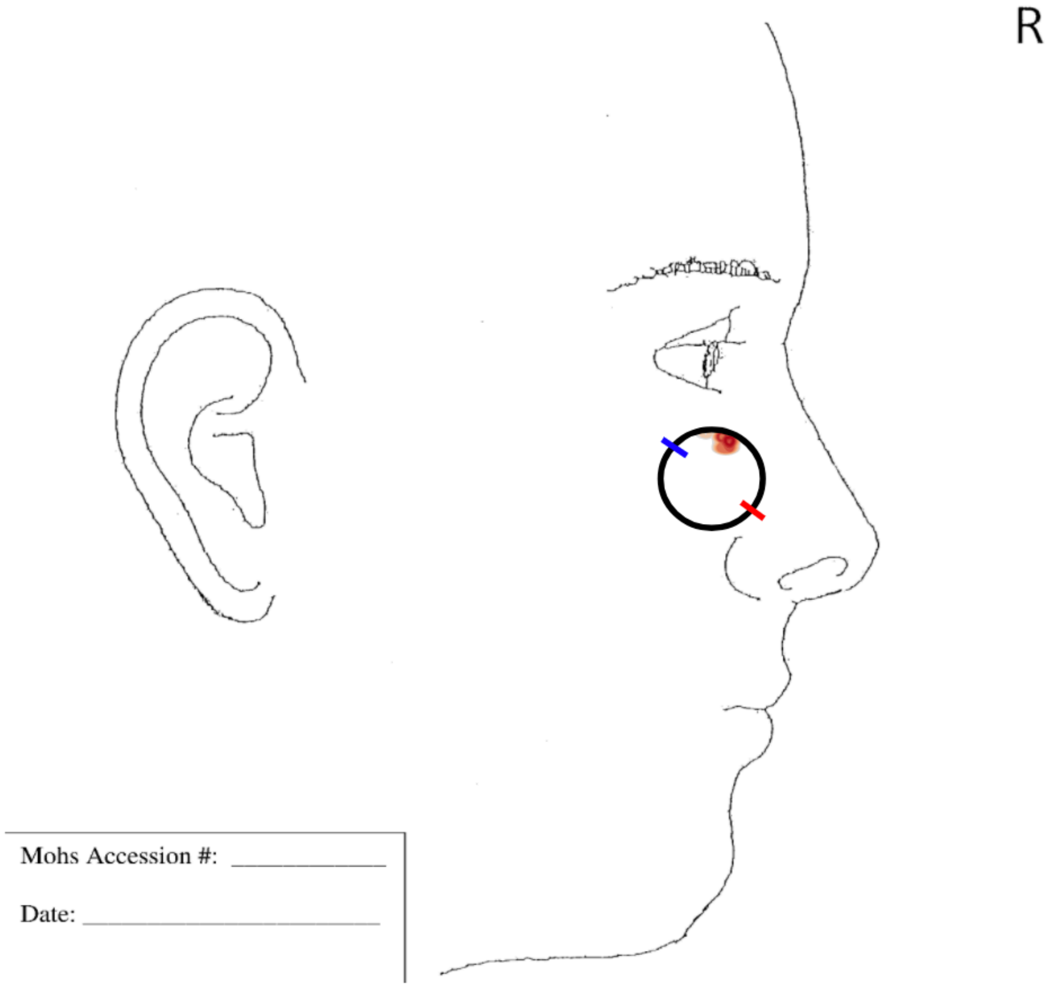
R
Mohs Accession #:
Date:

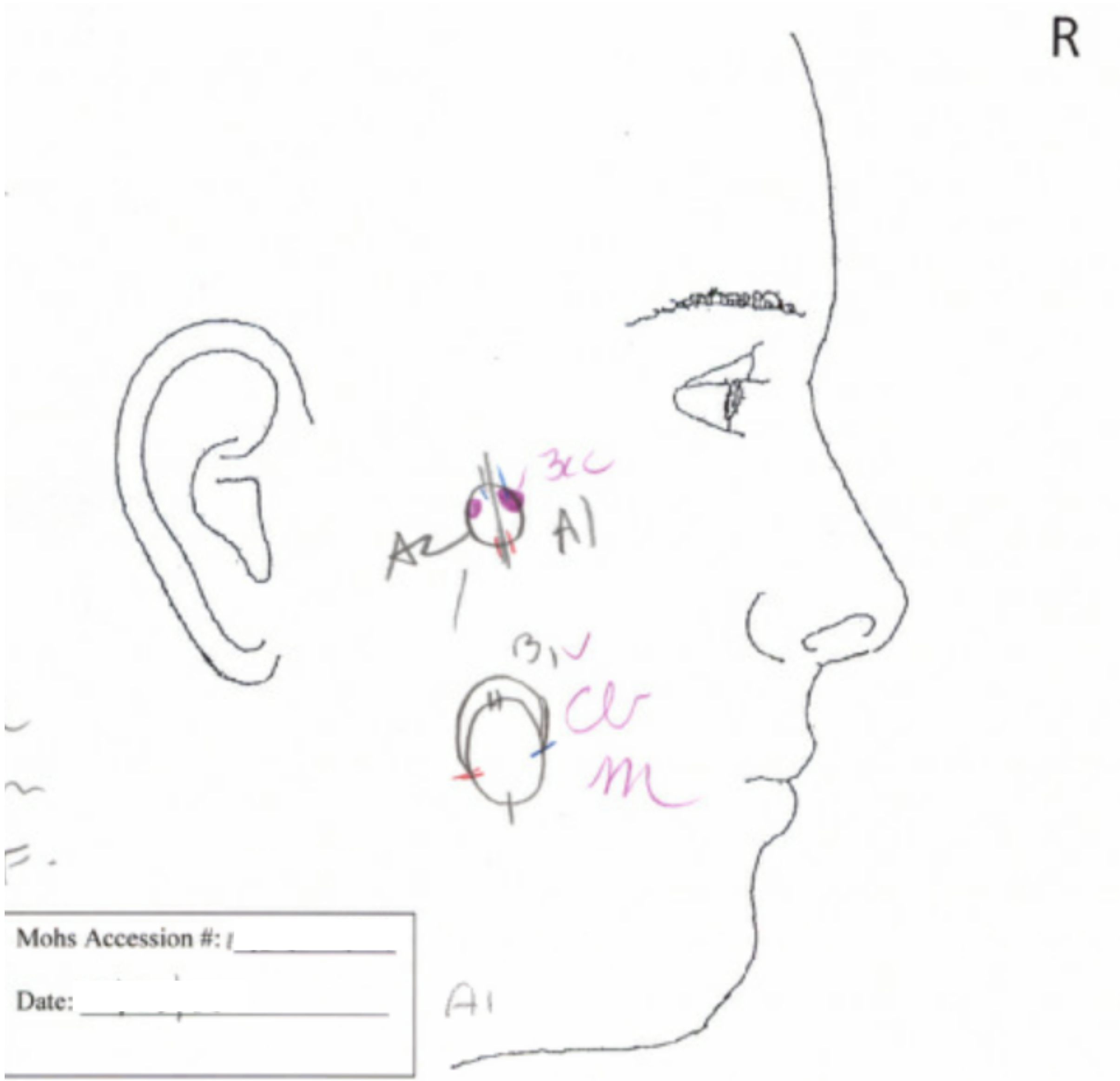
R
A2
A1
Mohs Accession #:
Date:
A1

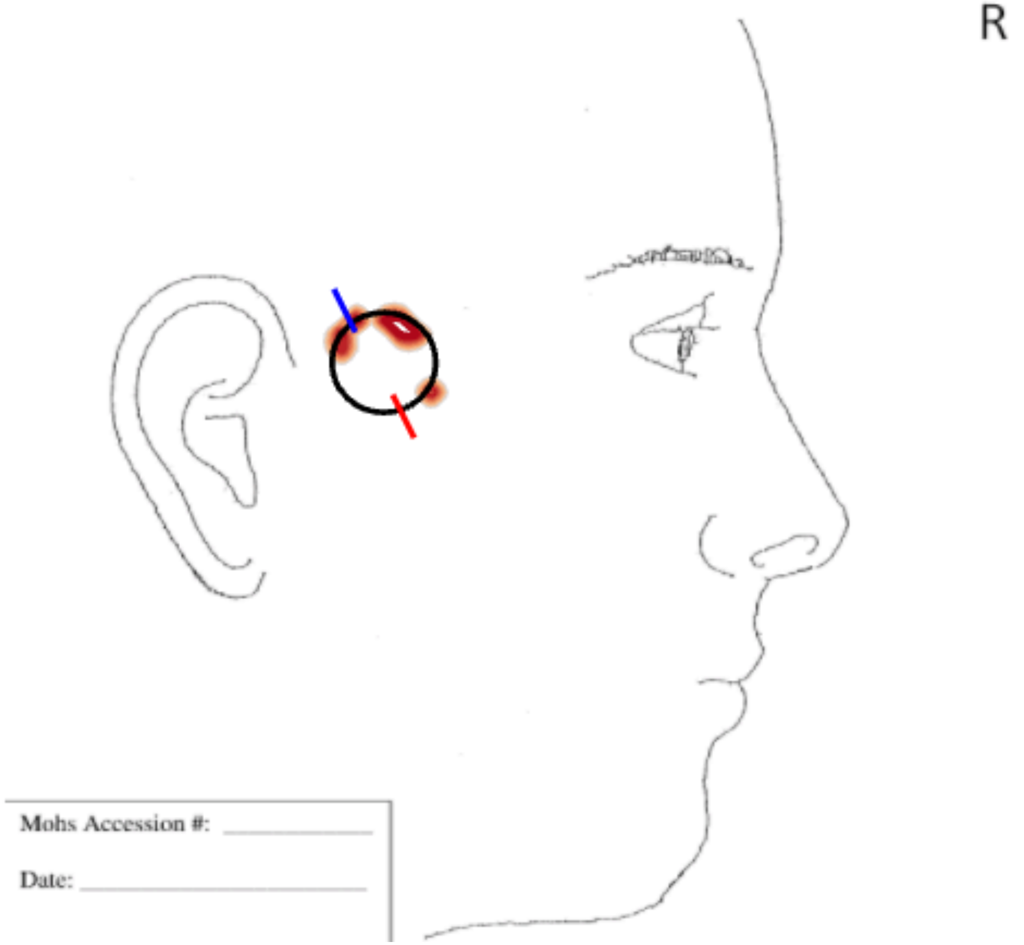
R
Mohs Accession #:
Date:

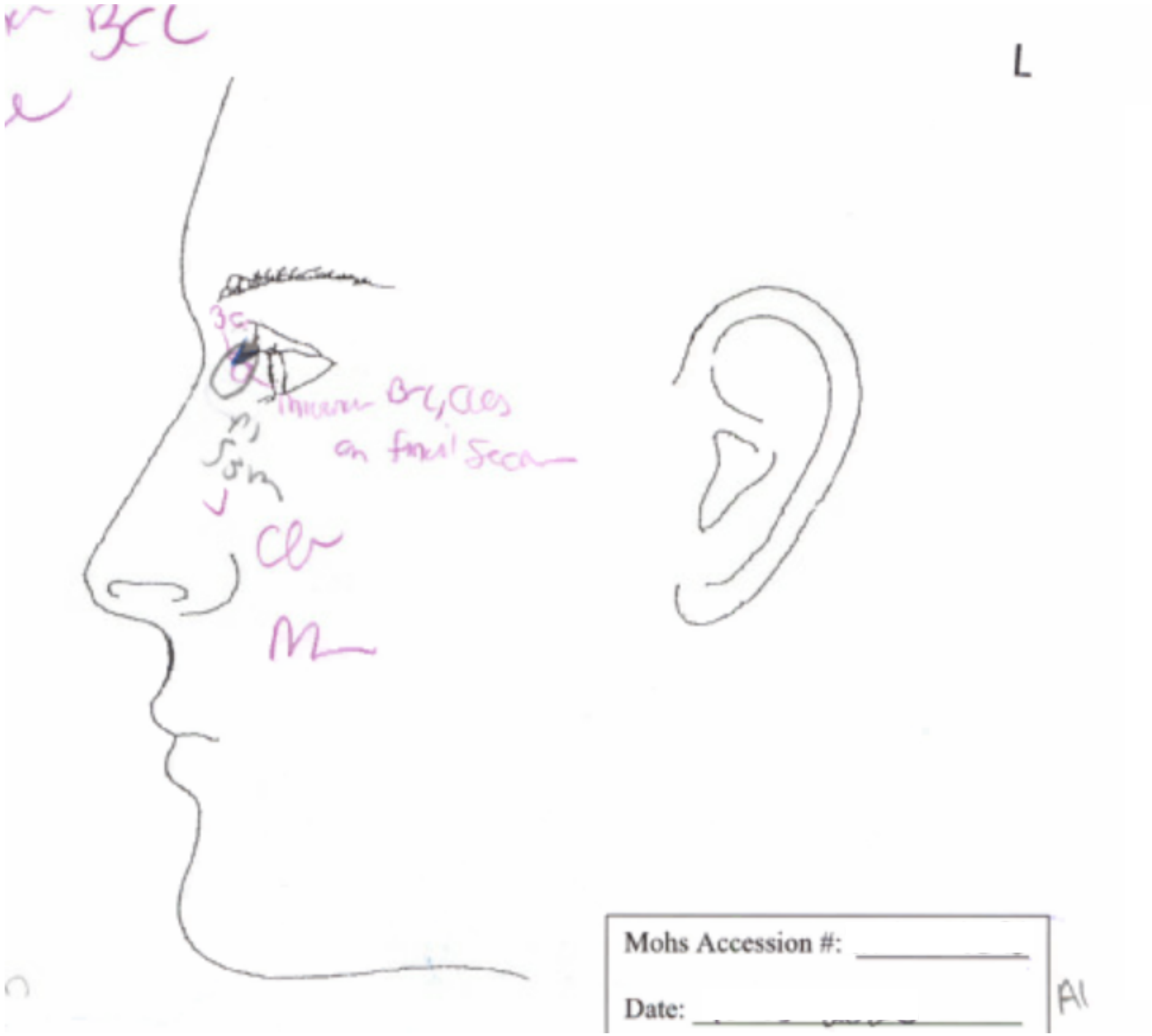
L
Mohs Accession #:
Date:

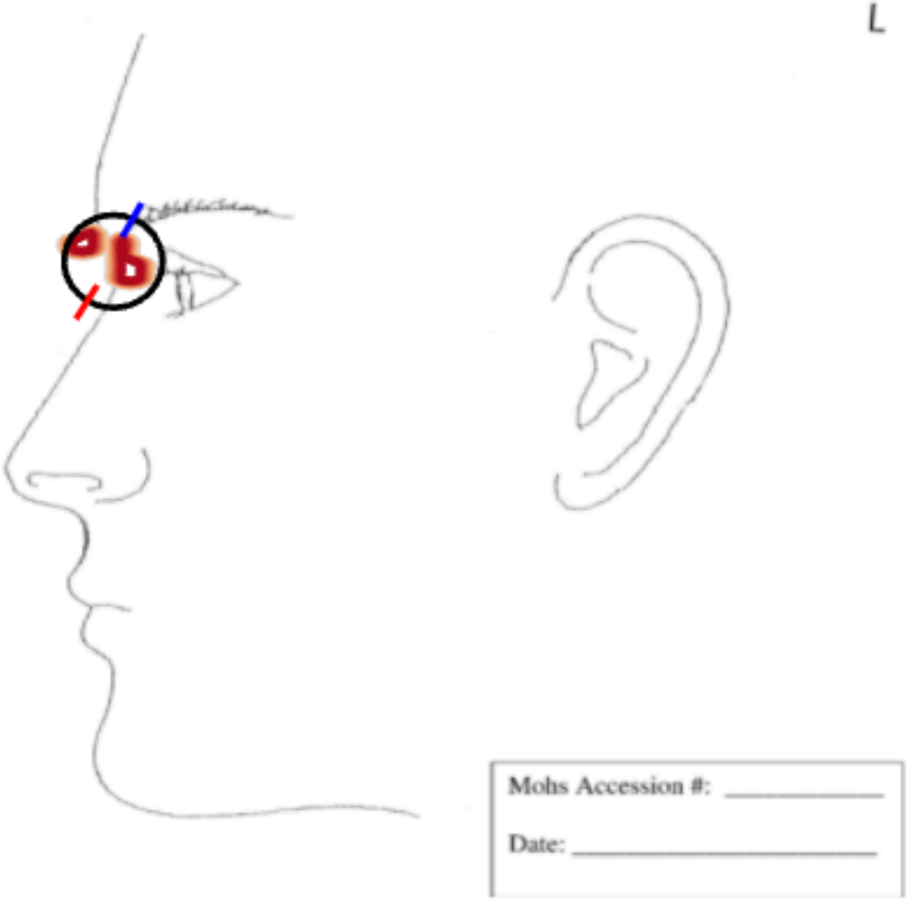
L
Mohs Accession #:
Date:

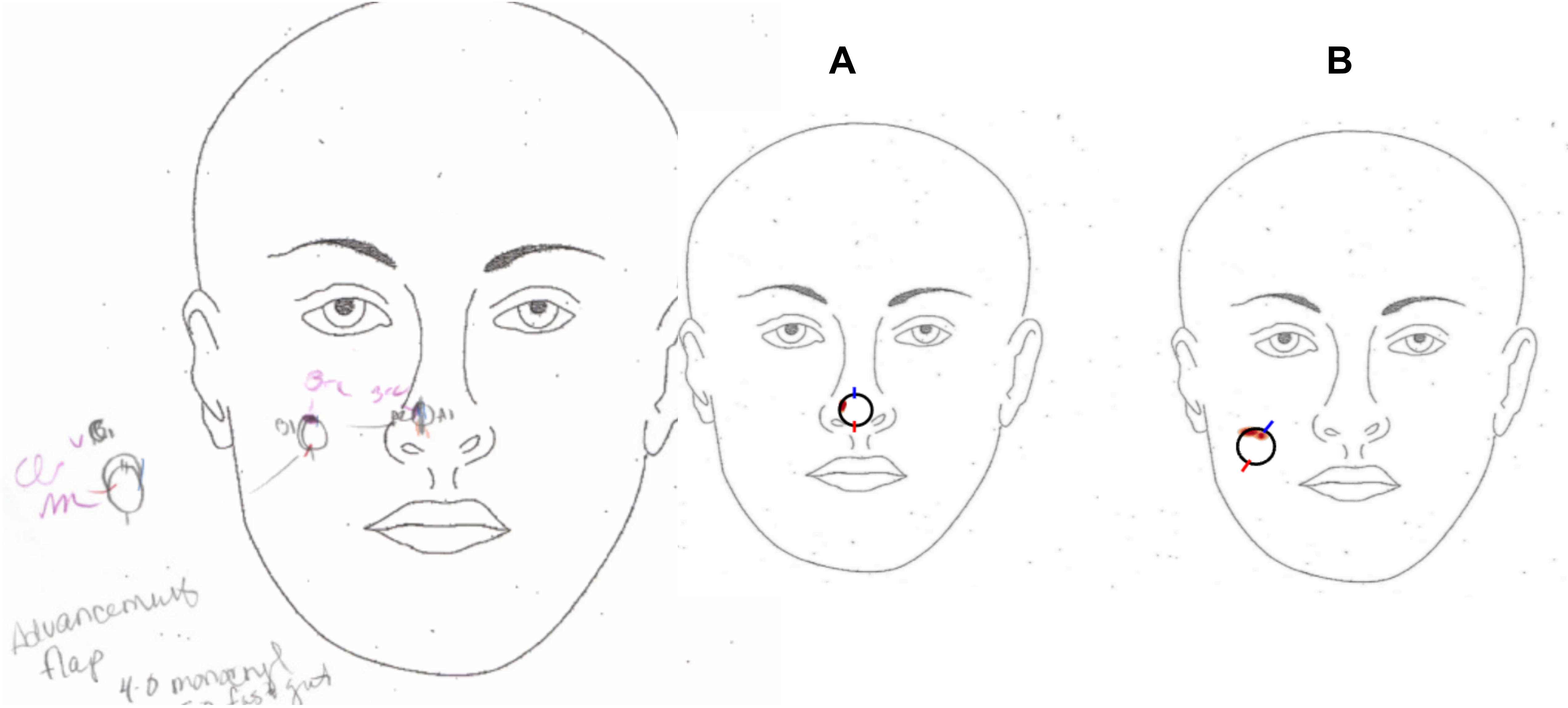
A
B
Advancement flap

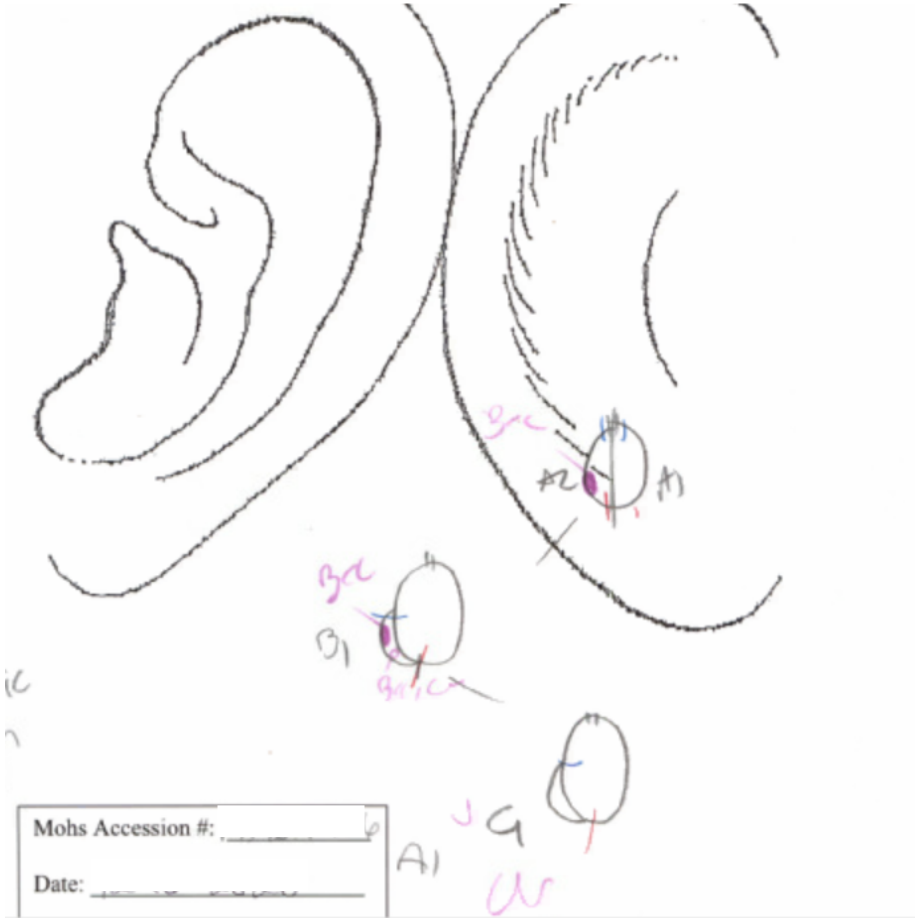


L

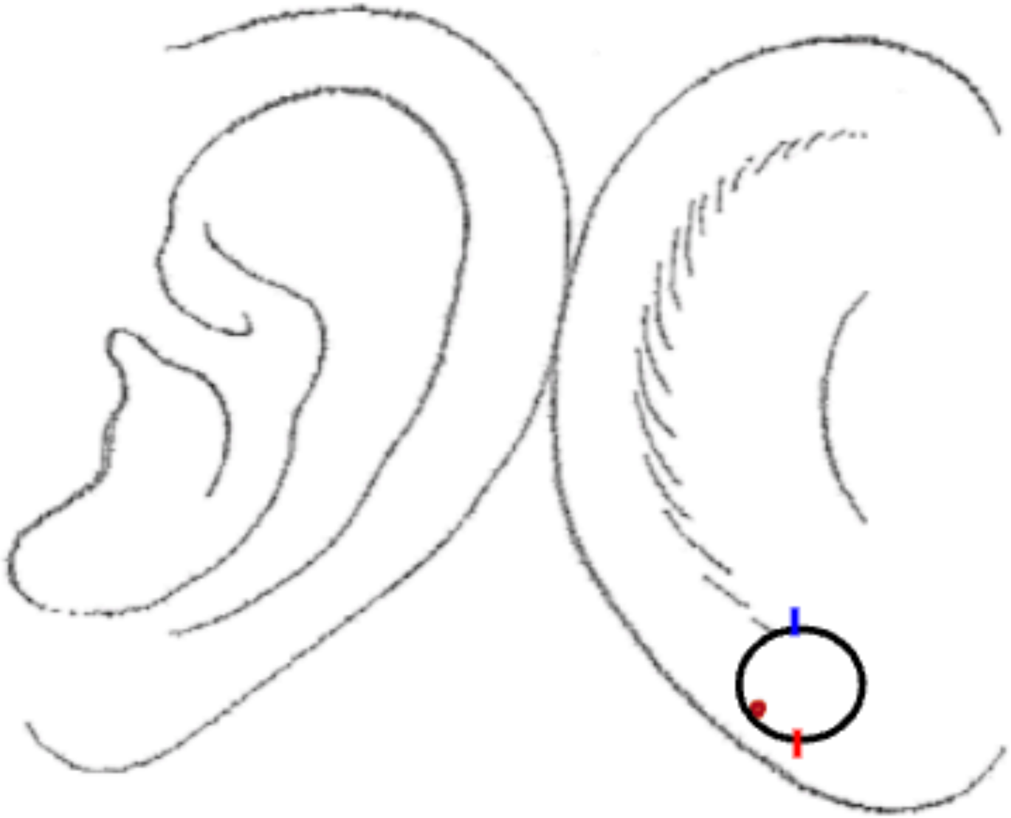

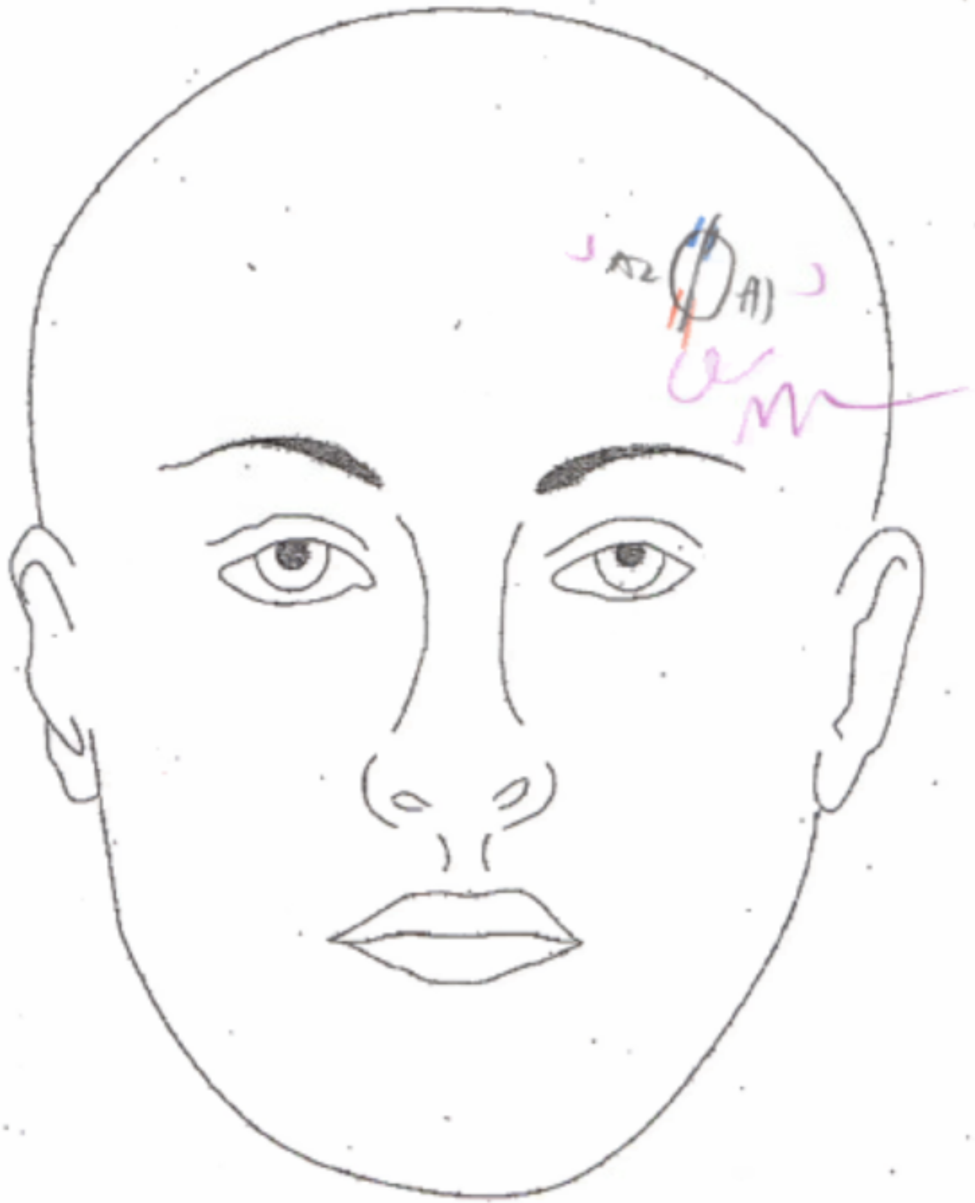
A2
A1

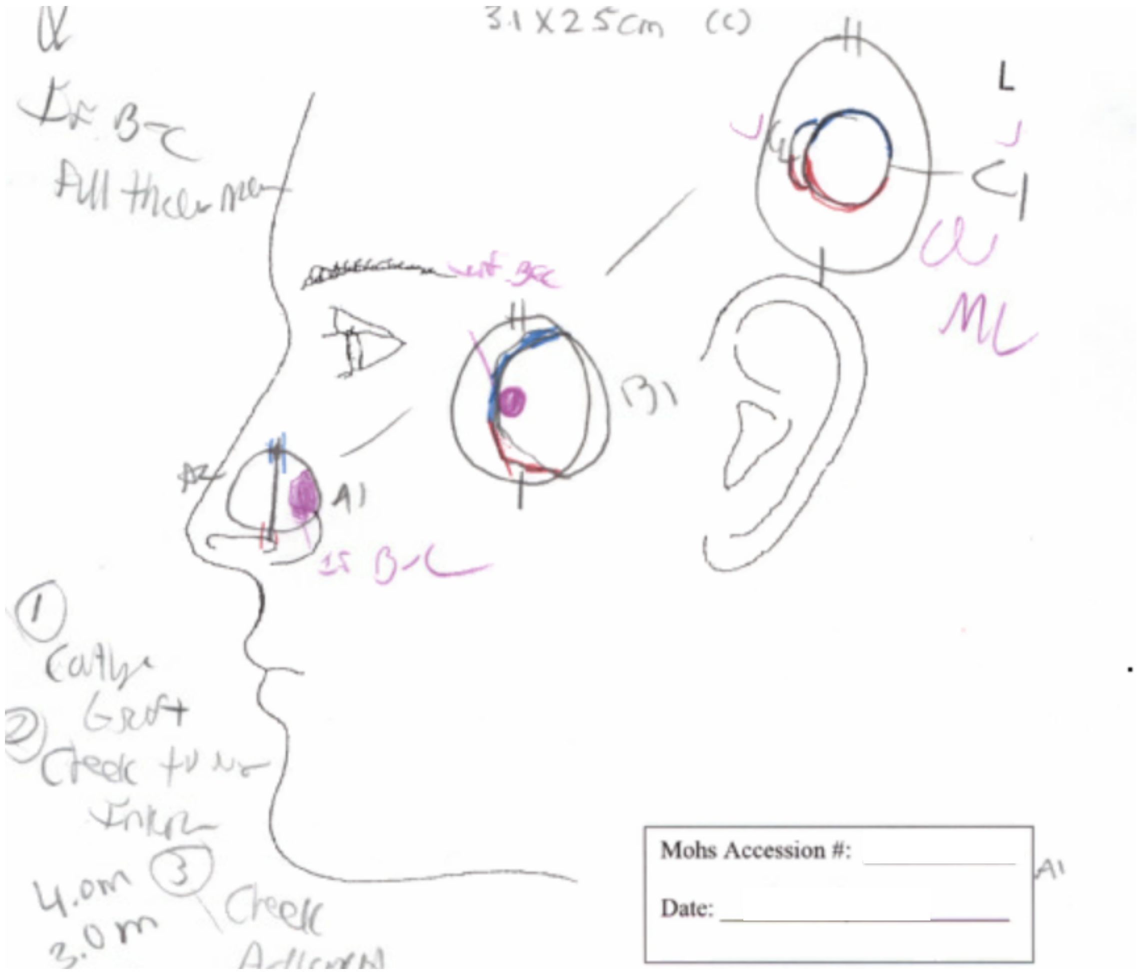


B

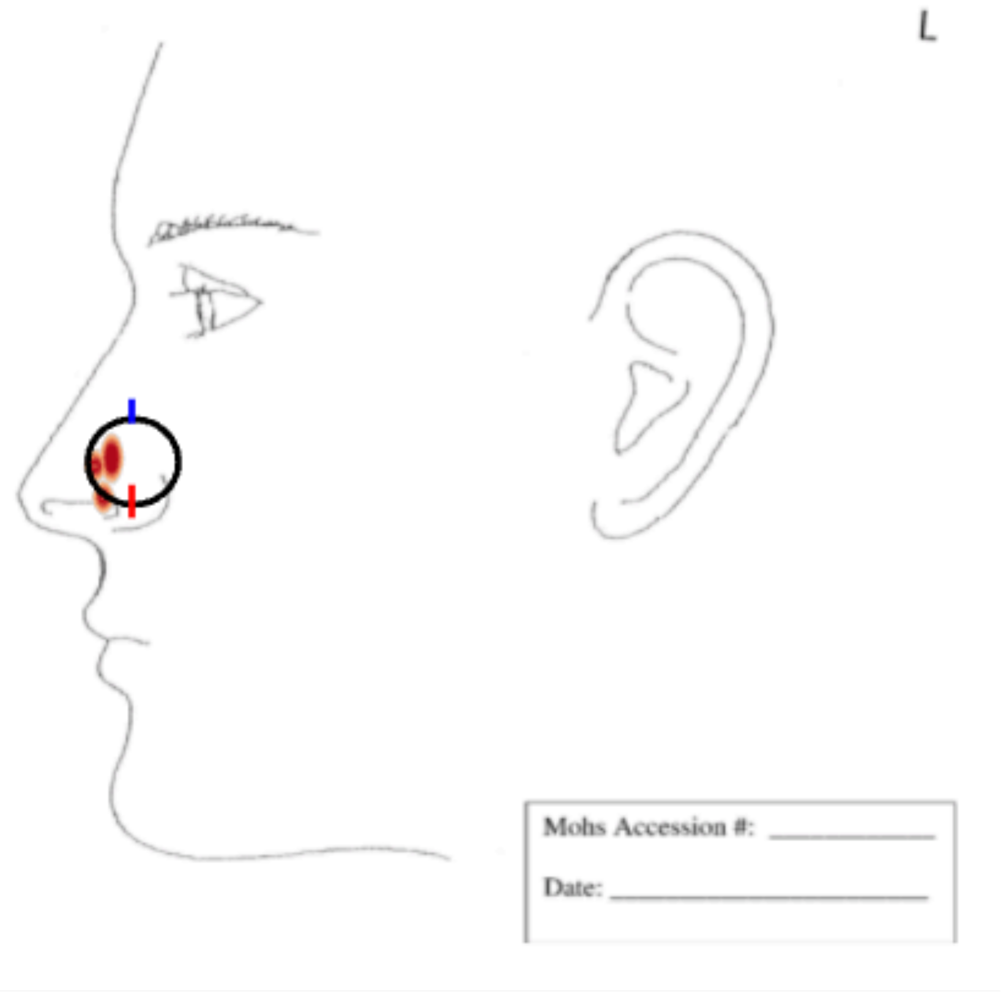

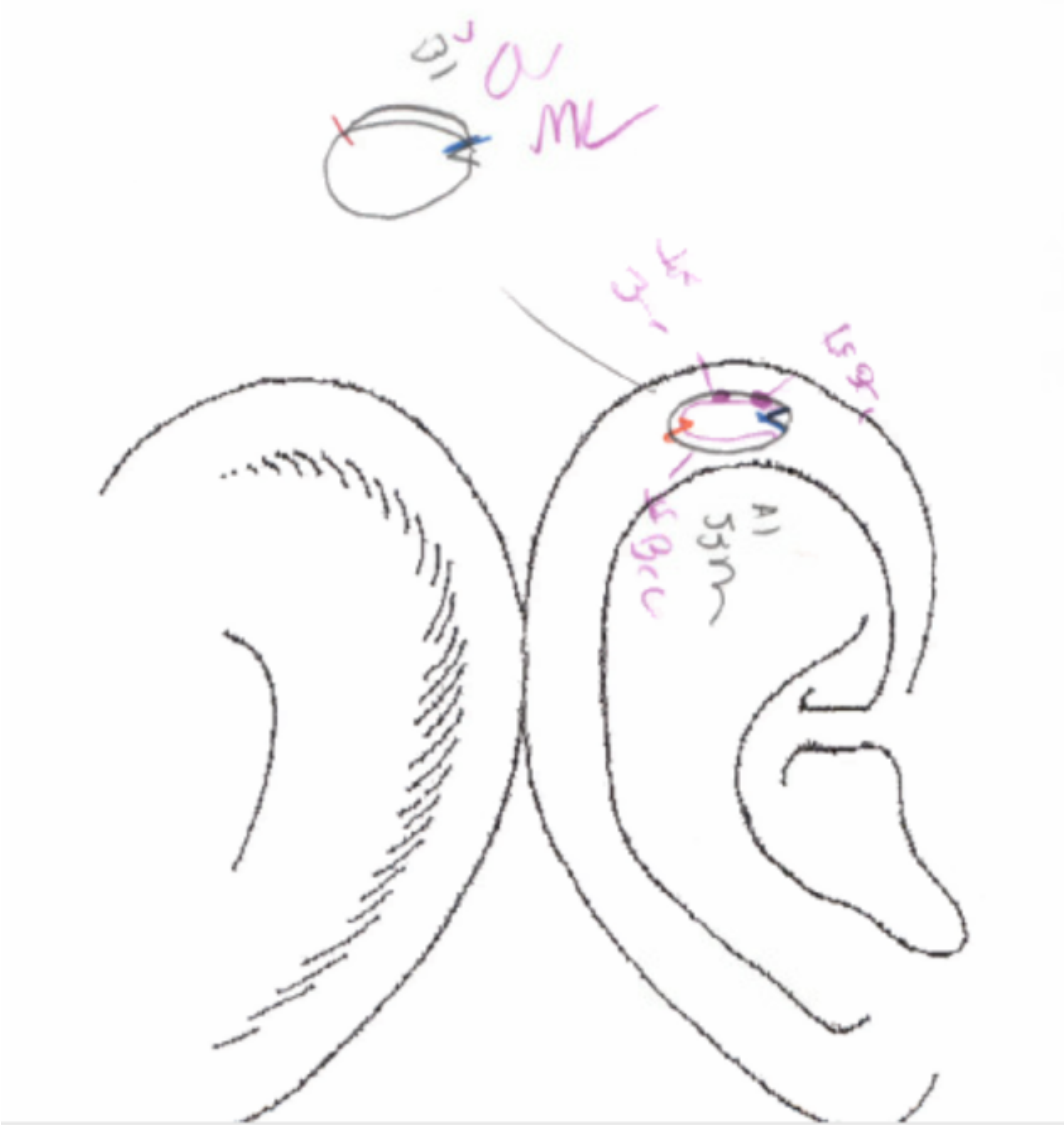

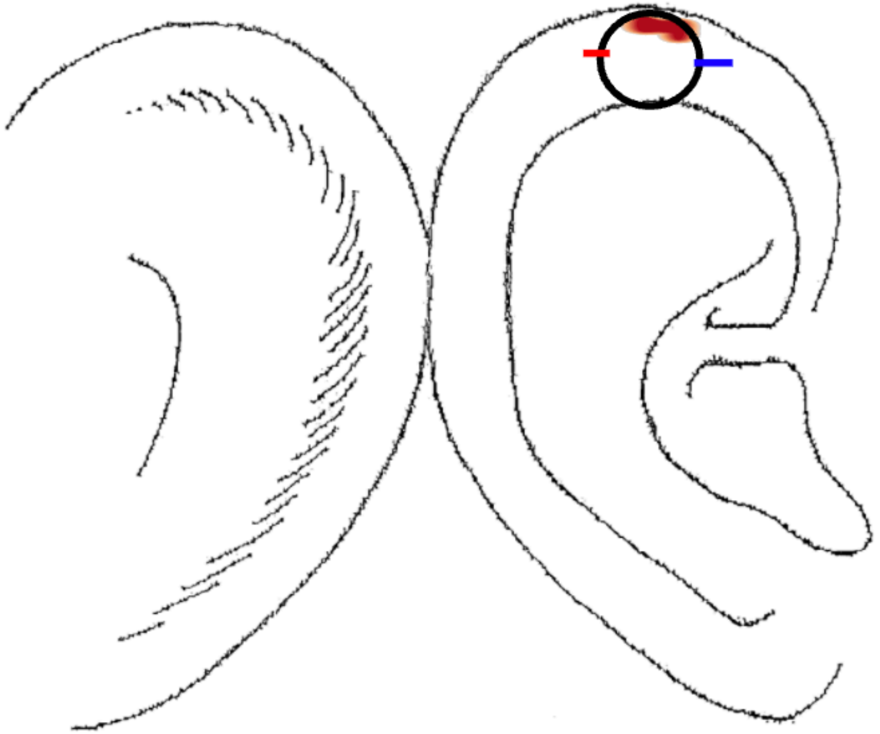

R

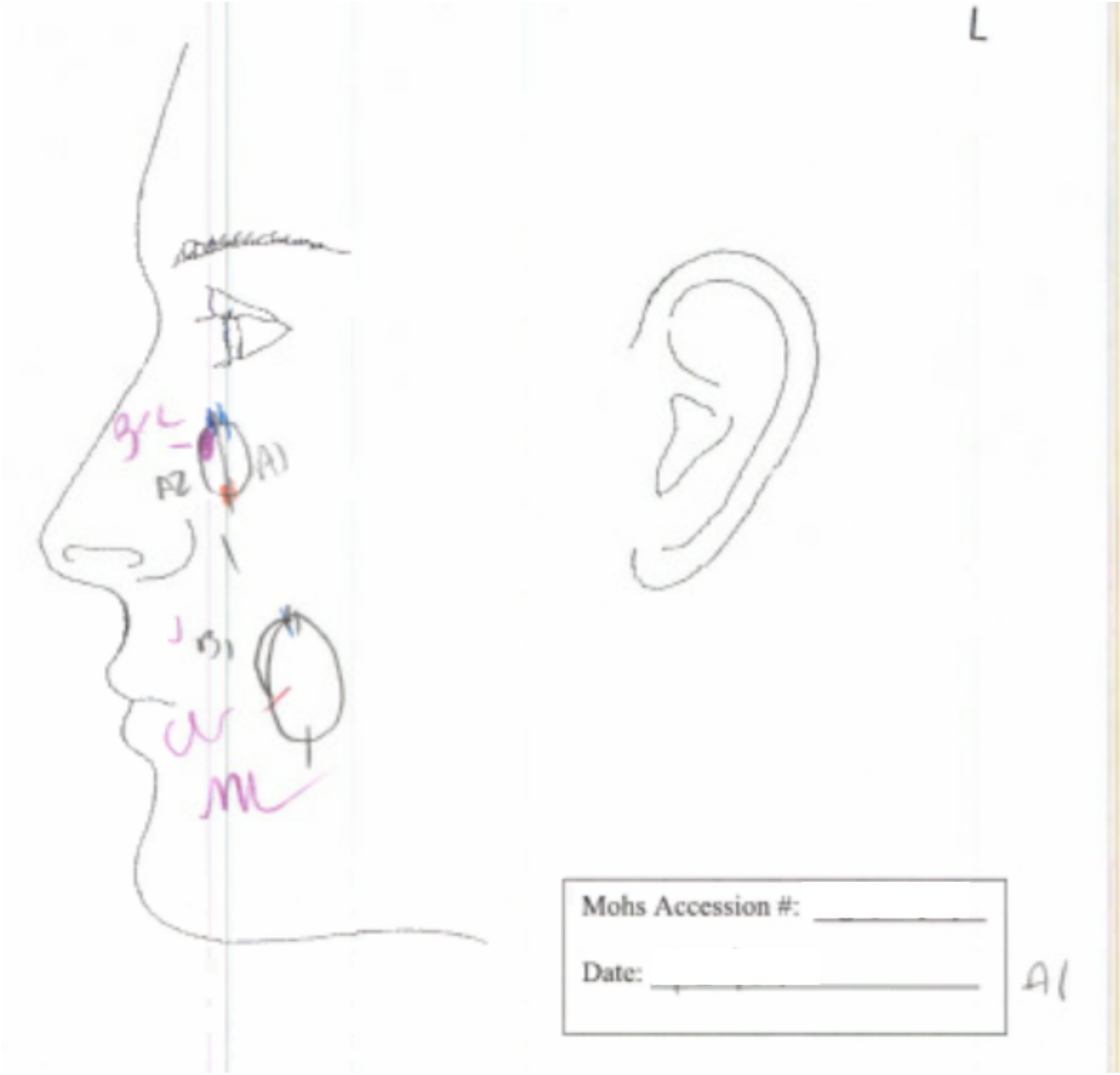
L
Mohs Accession #:
Date:
A1

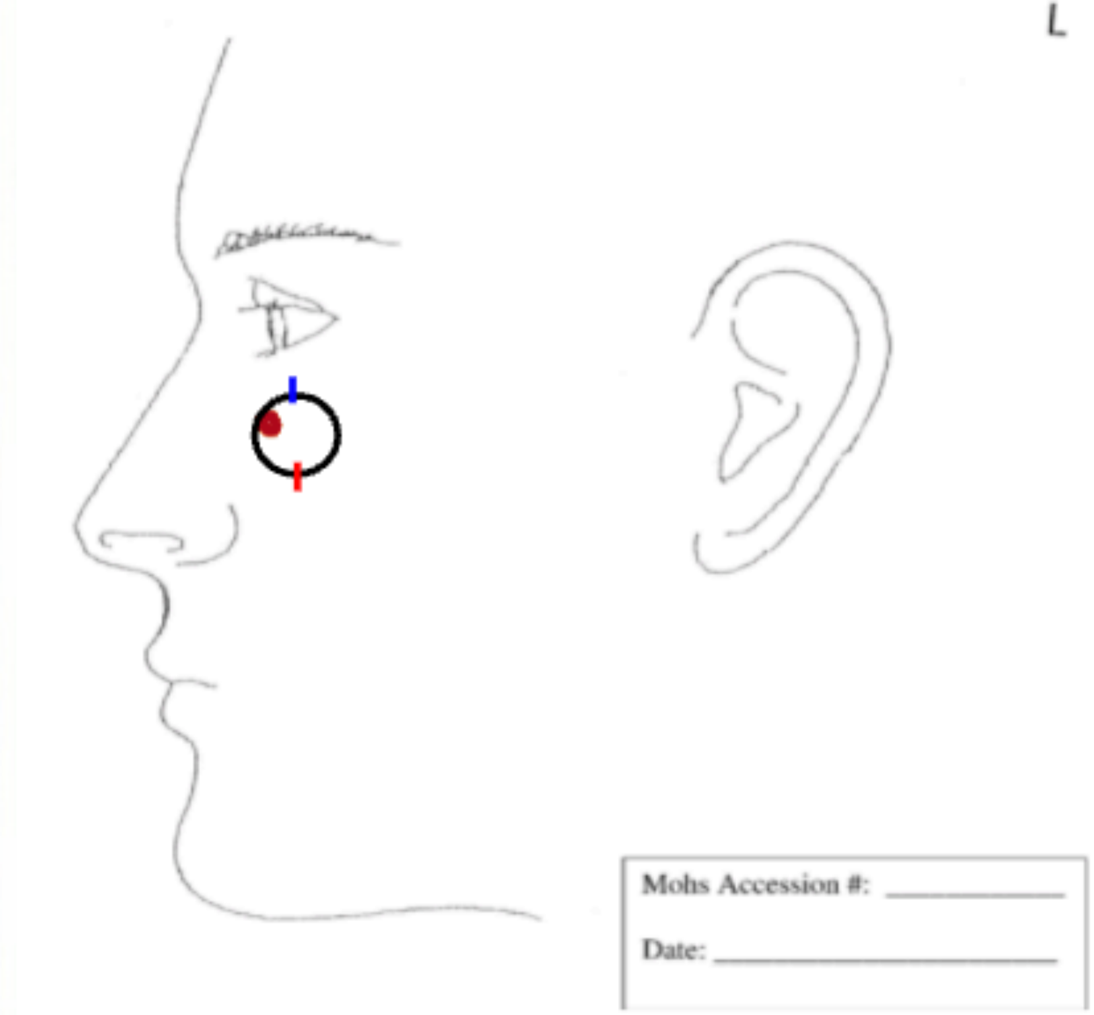
L
Mohs Accession #:
Date:

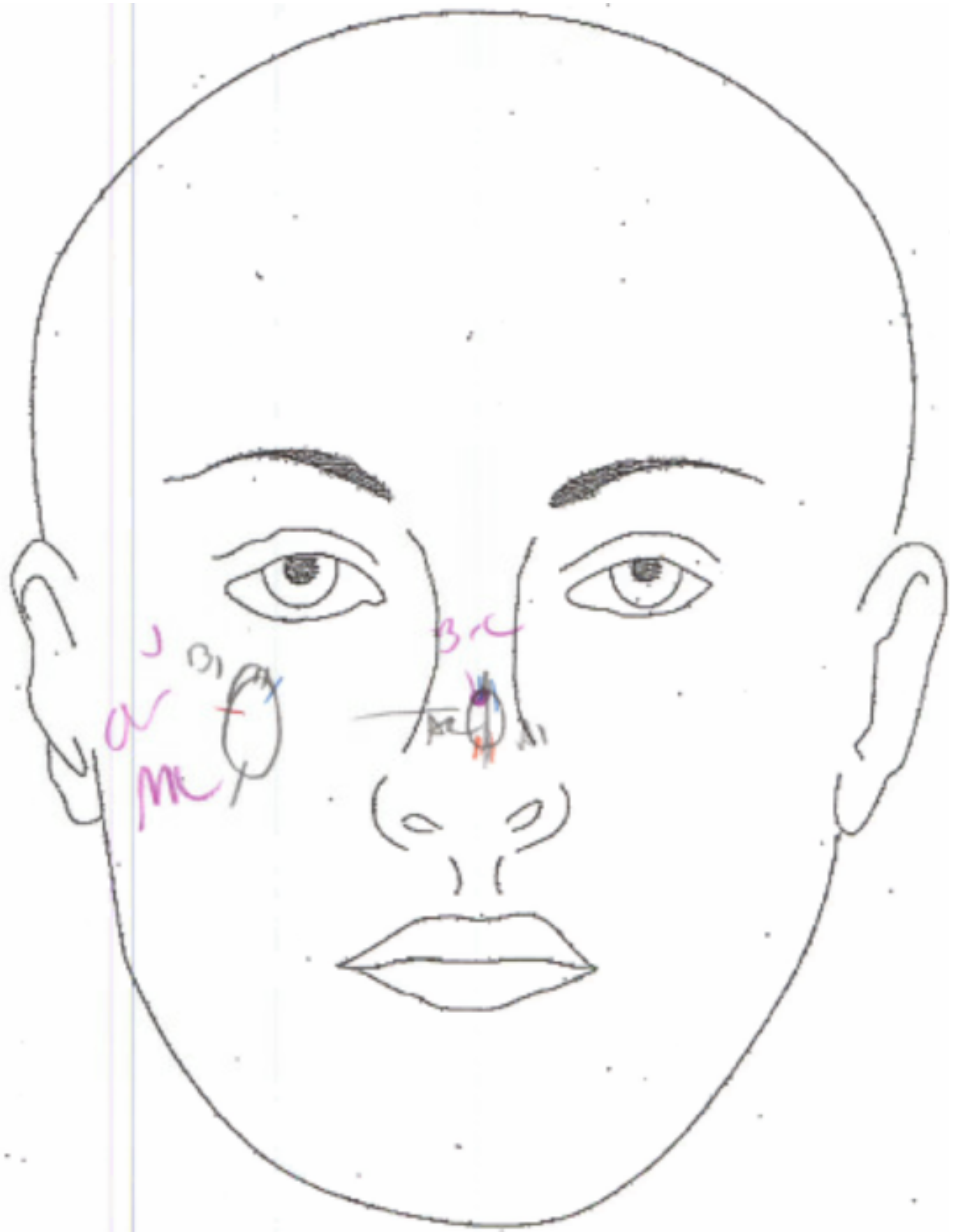

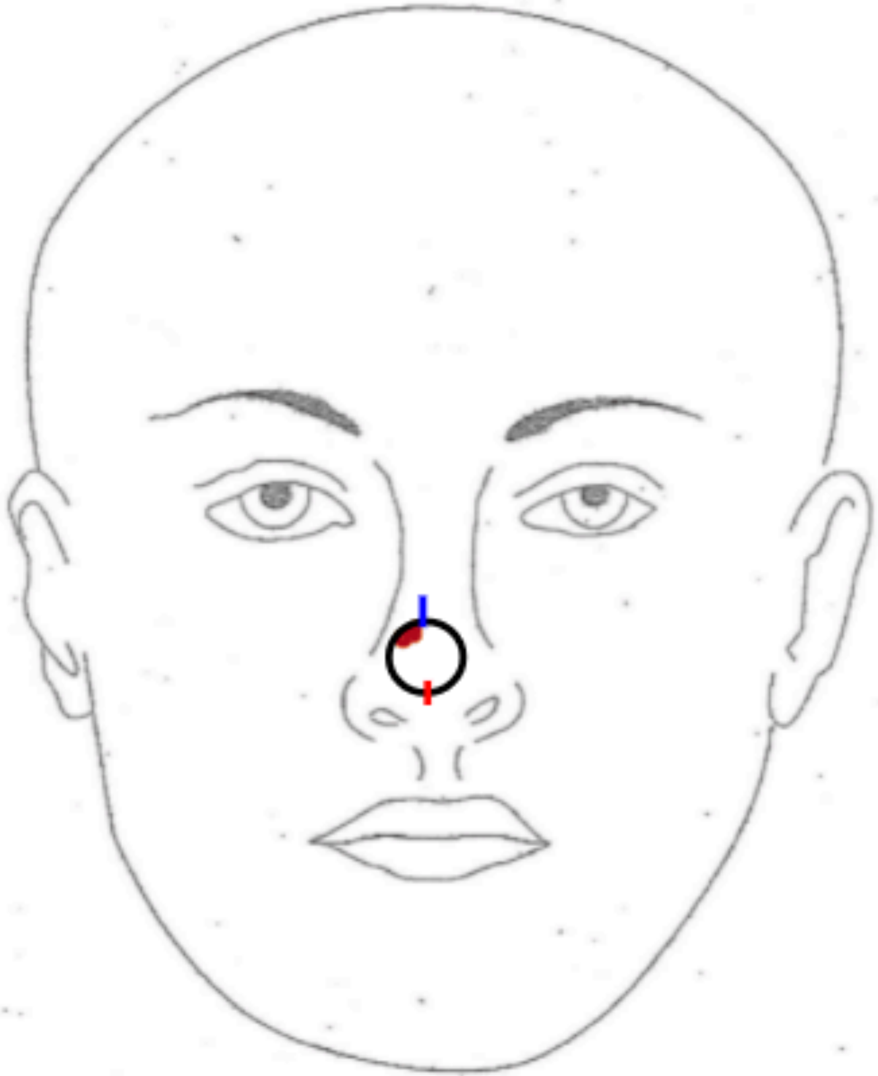

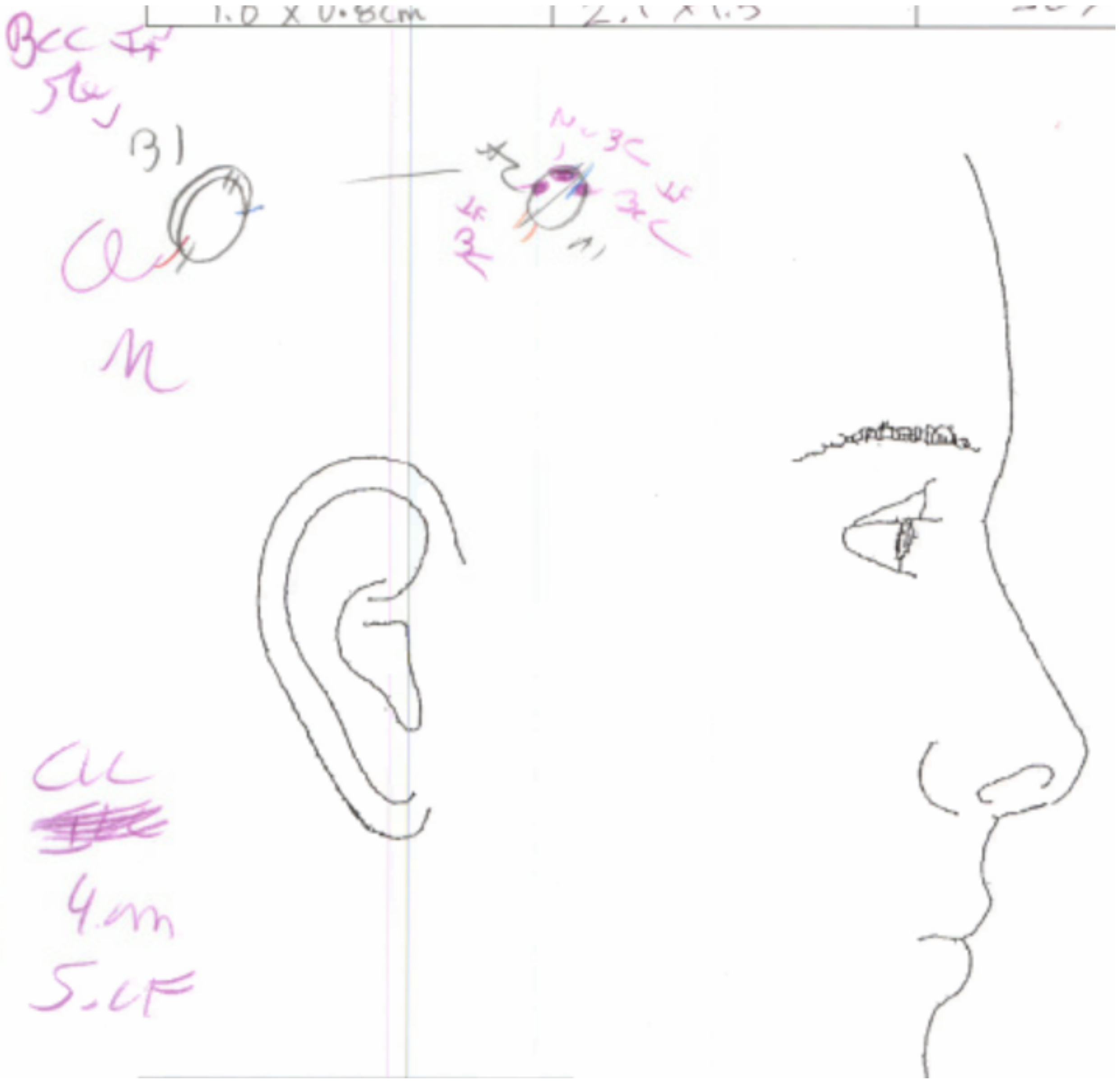

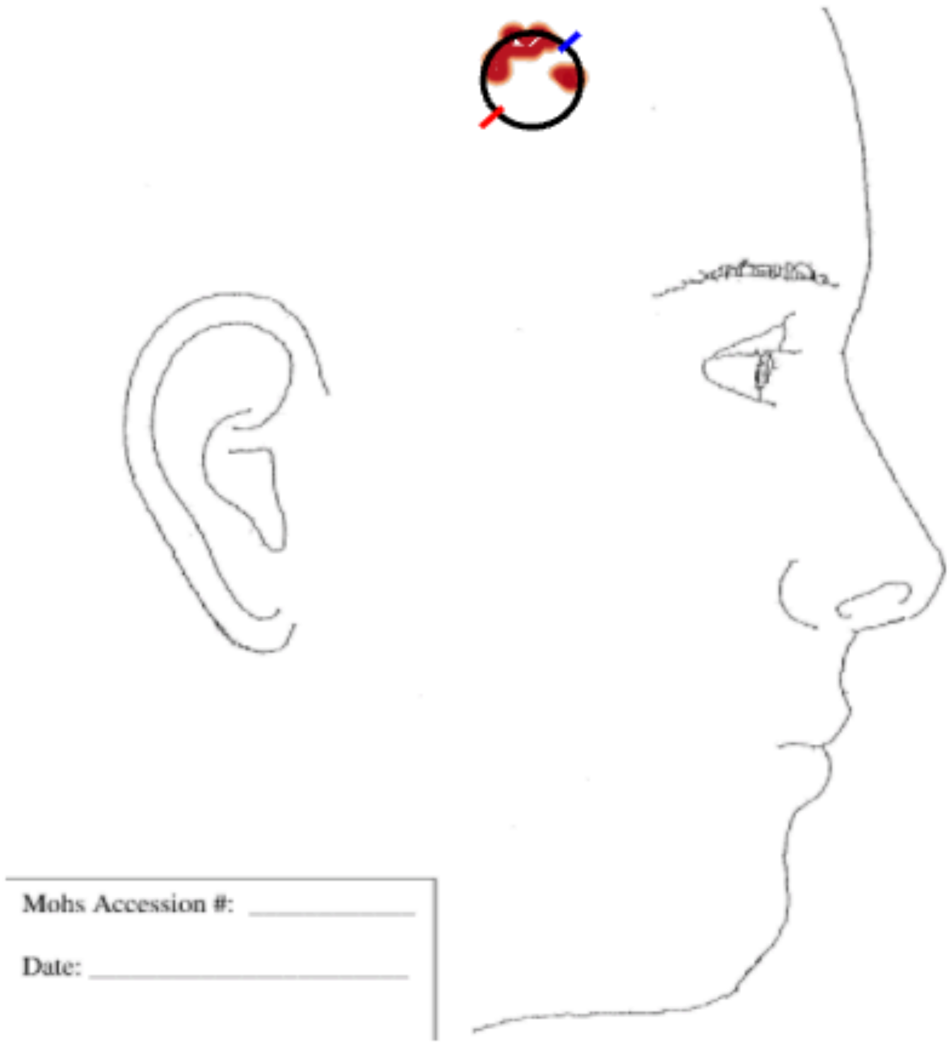
Mohs Accession #:
Date:

R

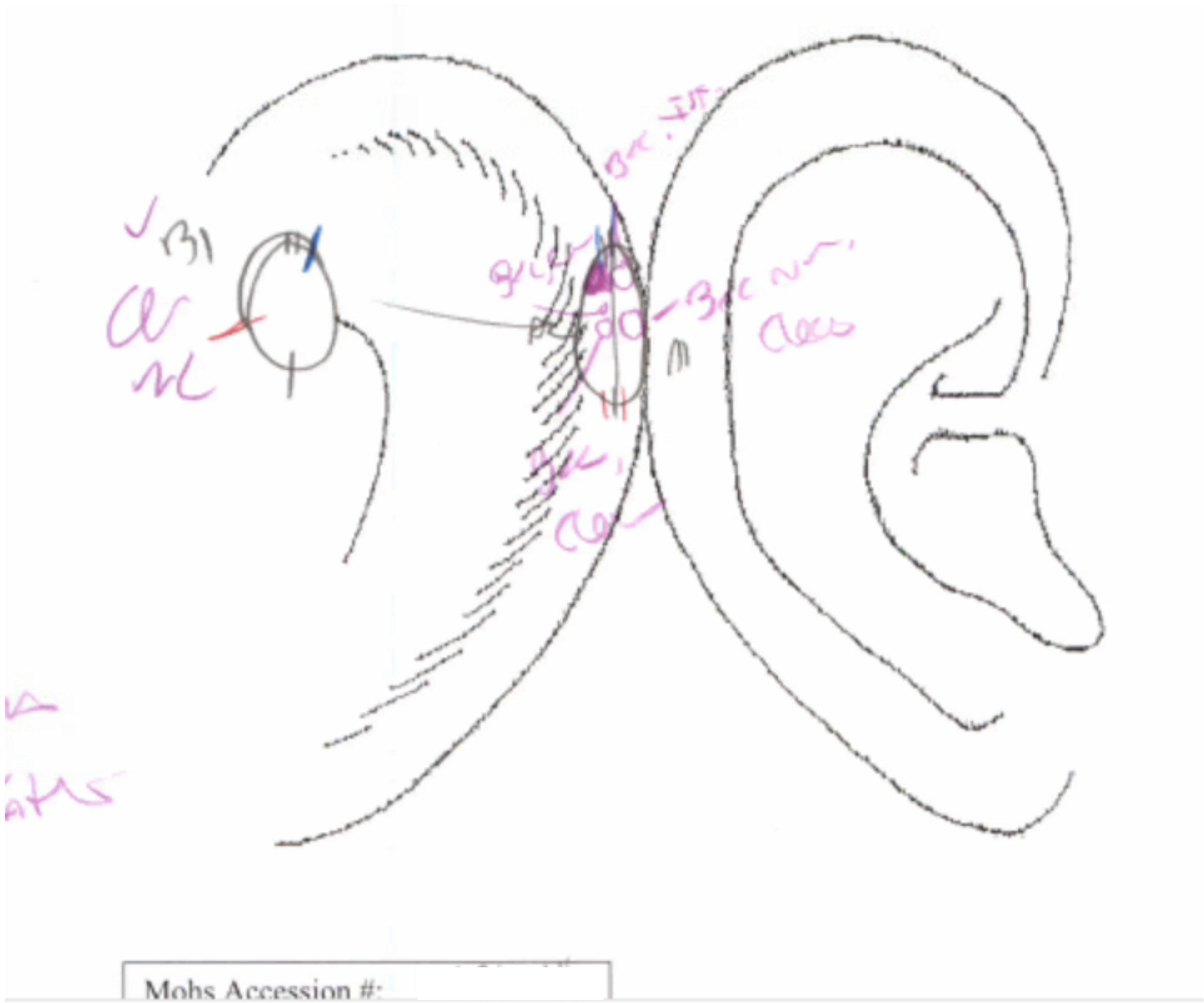


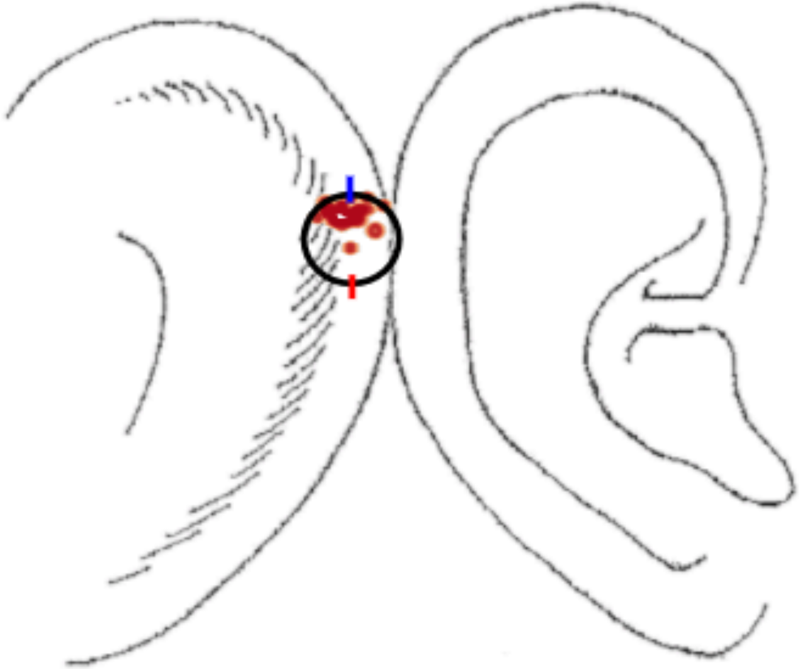

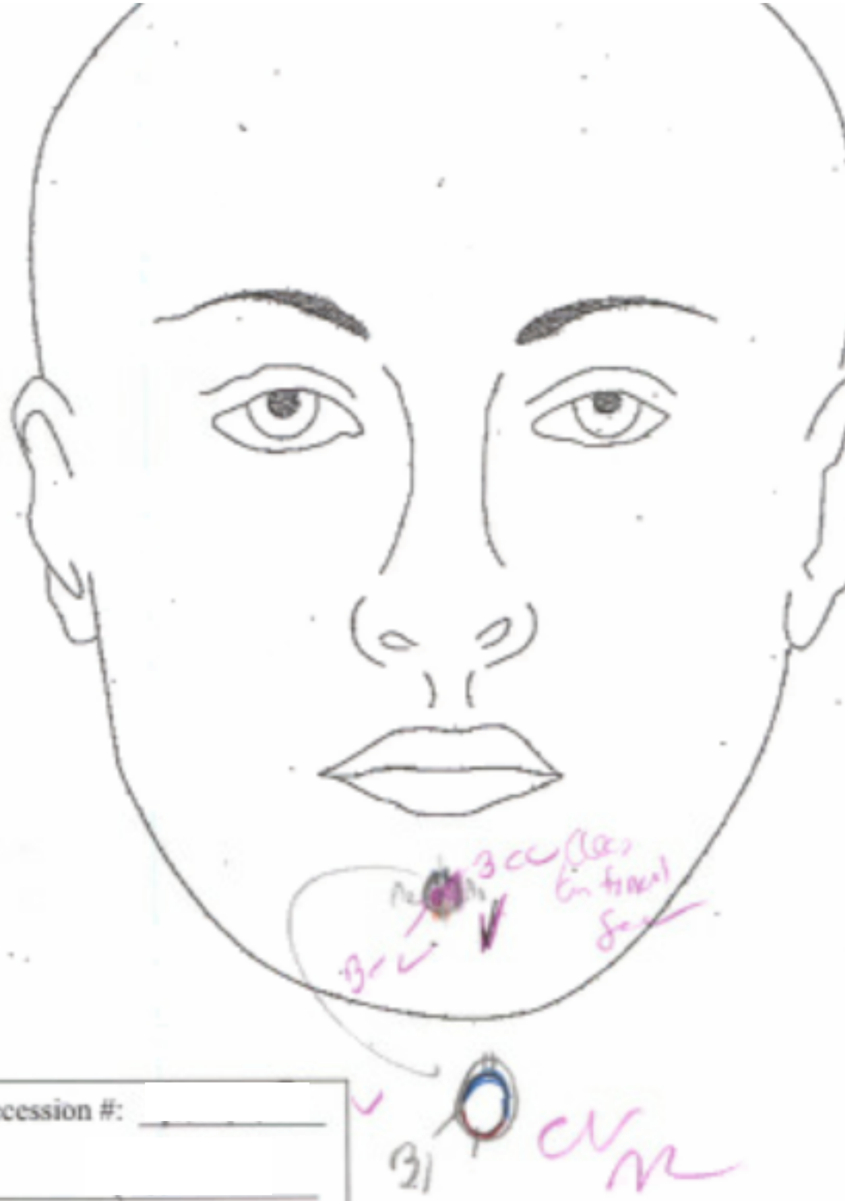

lohs Accession #: ____________

ate: ____________________

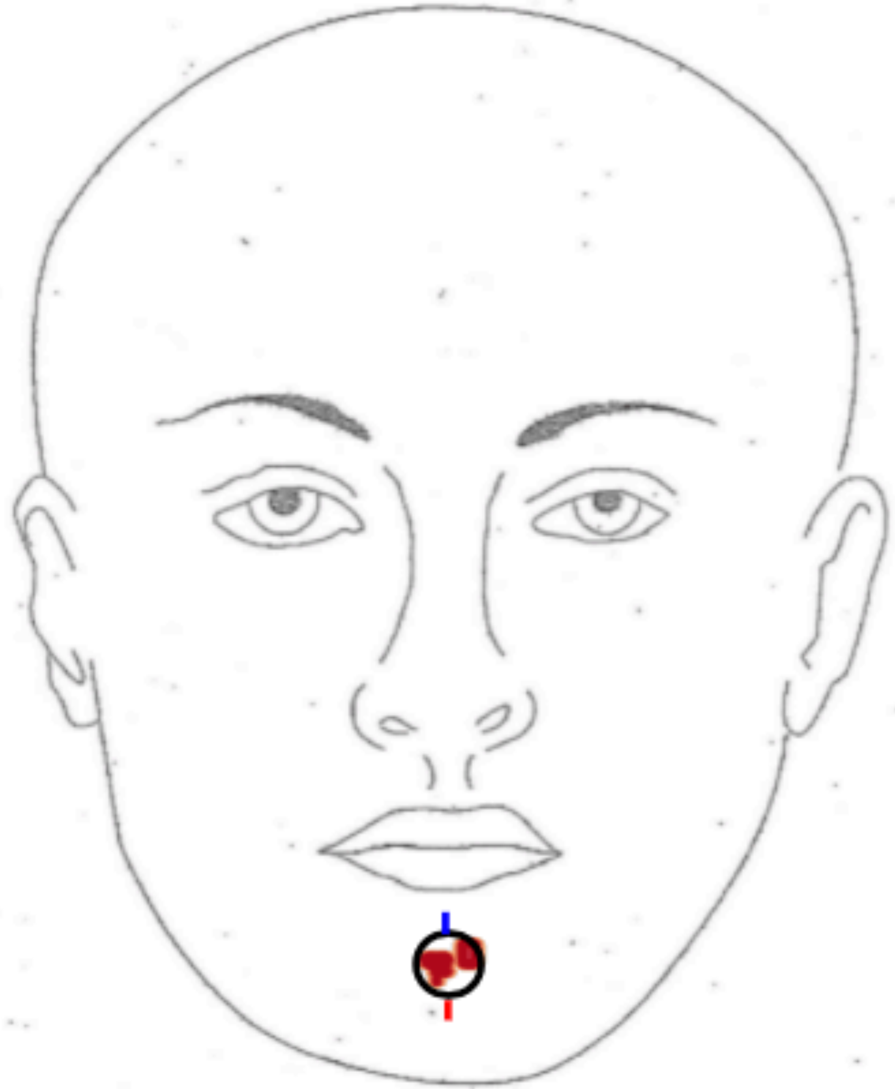

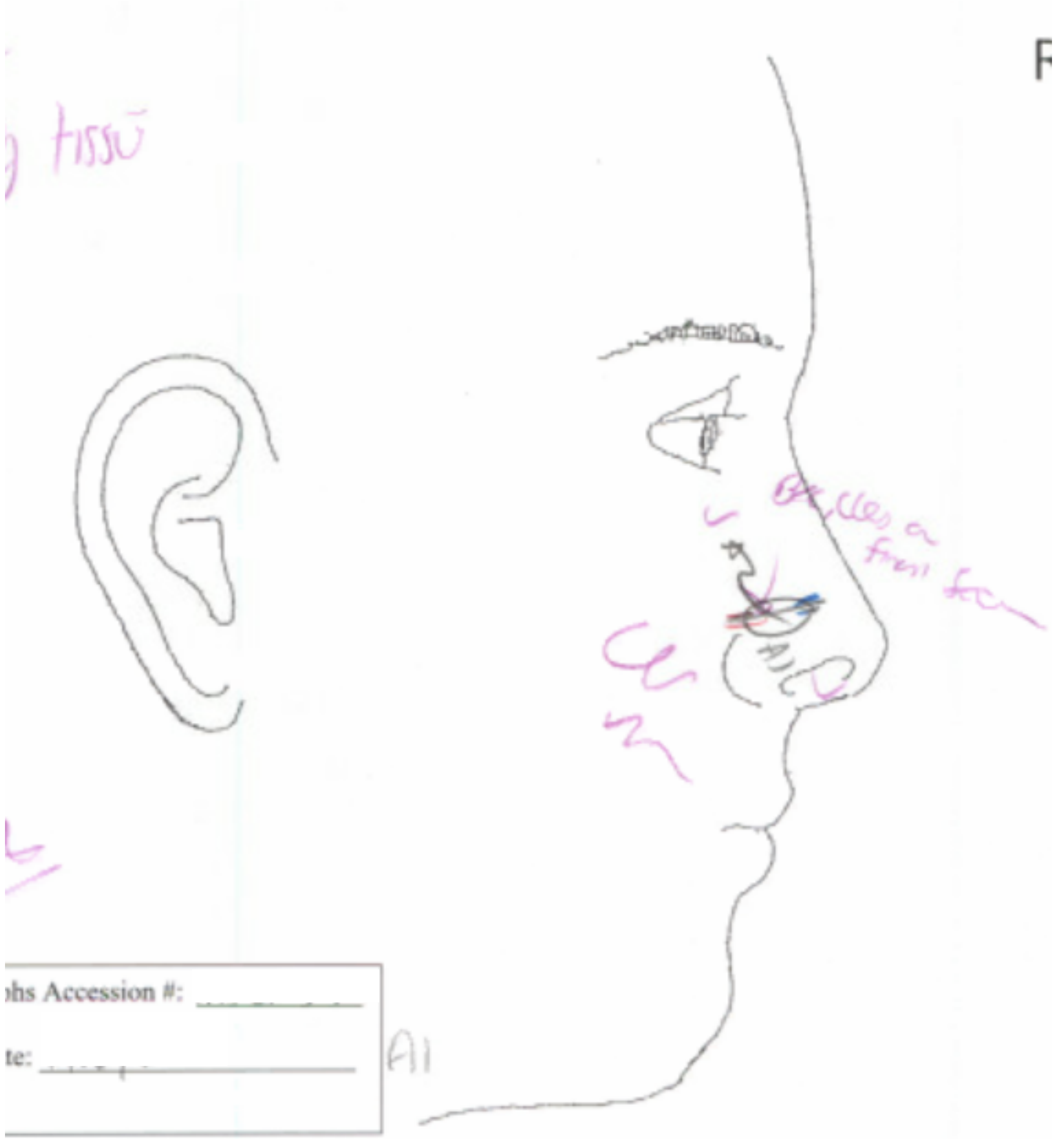
hs Accession #:
te:

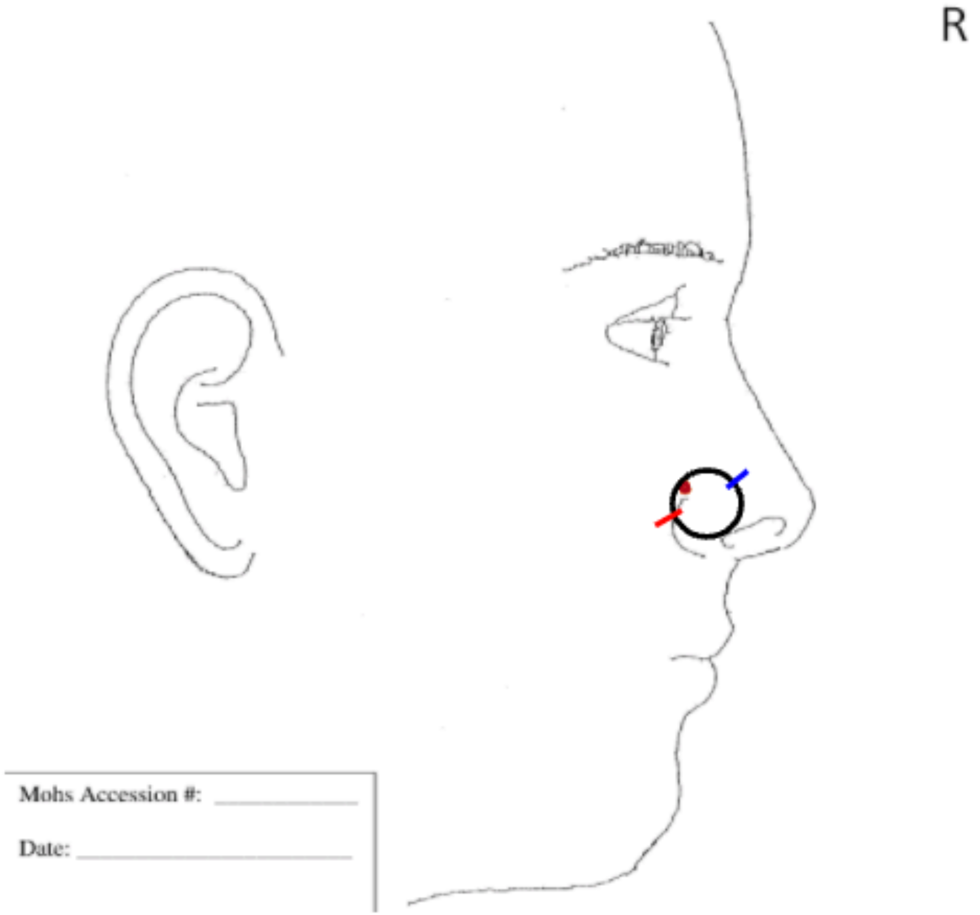
R
Mohs Accession #:
Date:

R

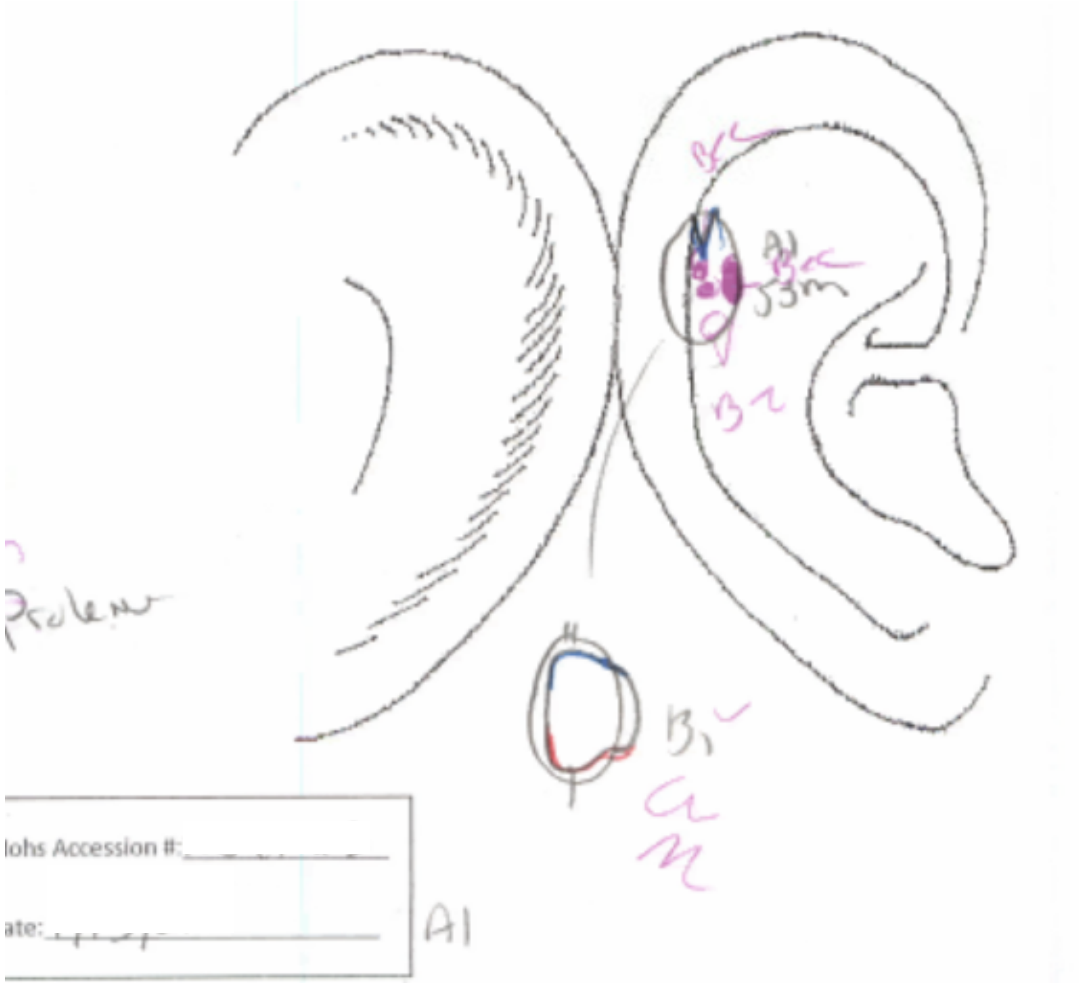


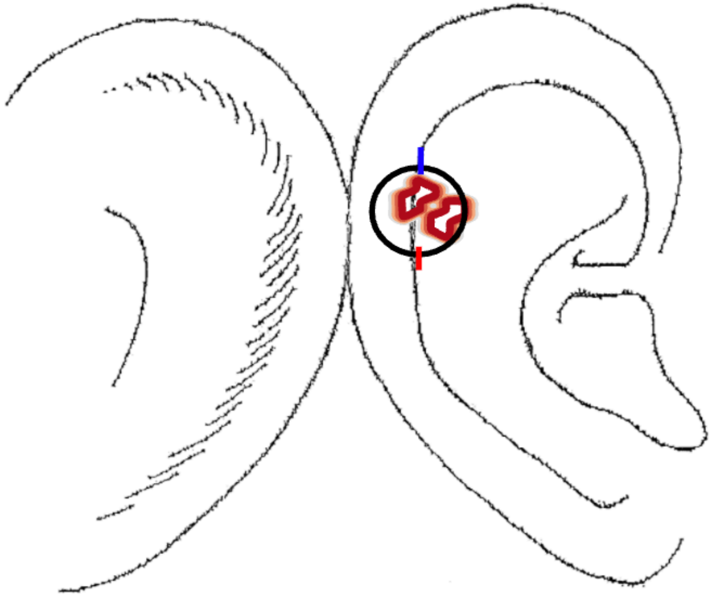

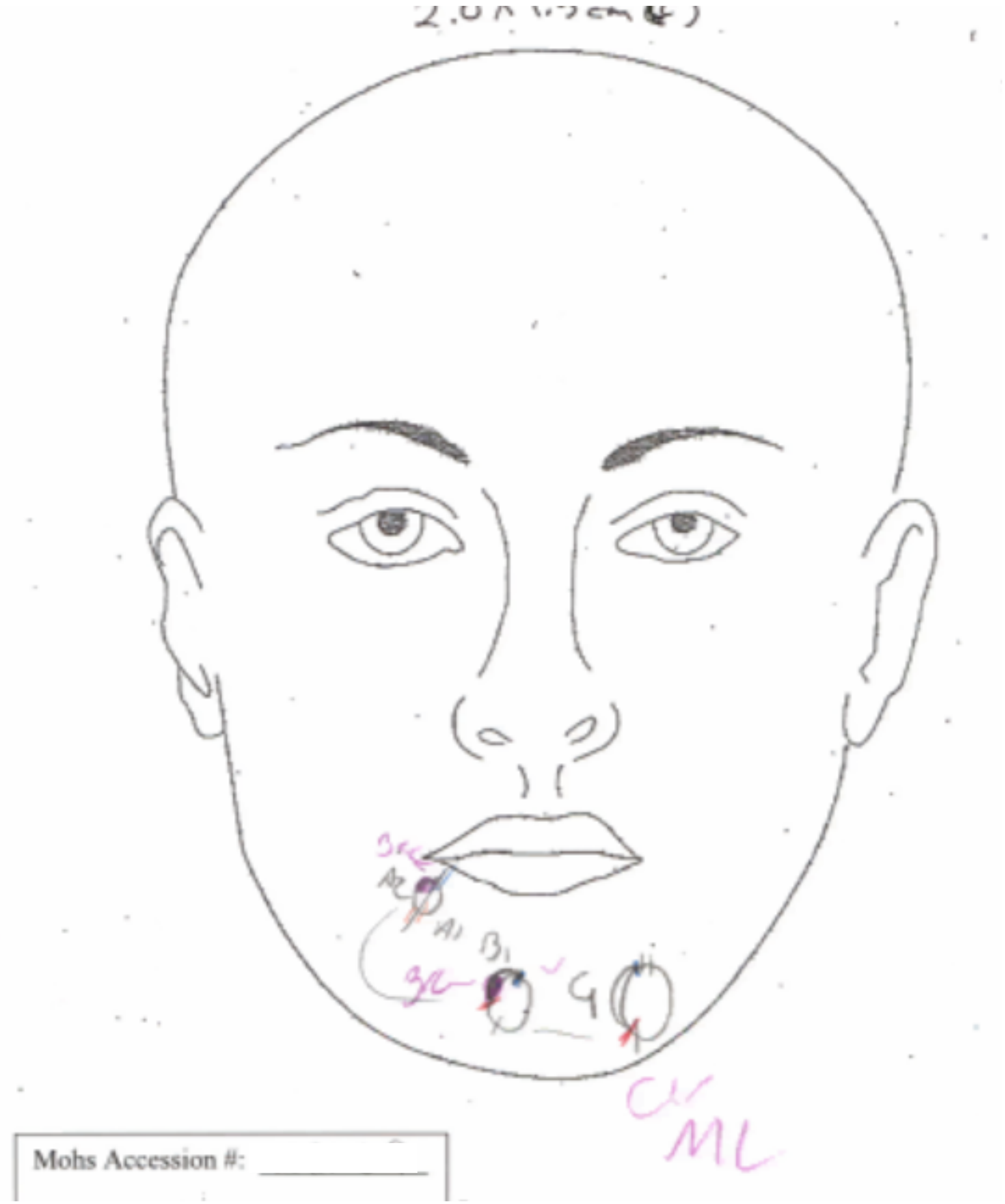


A

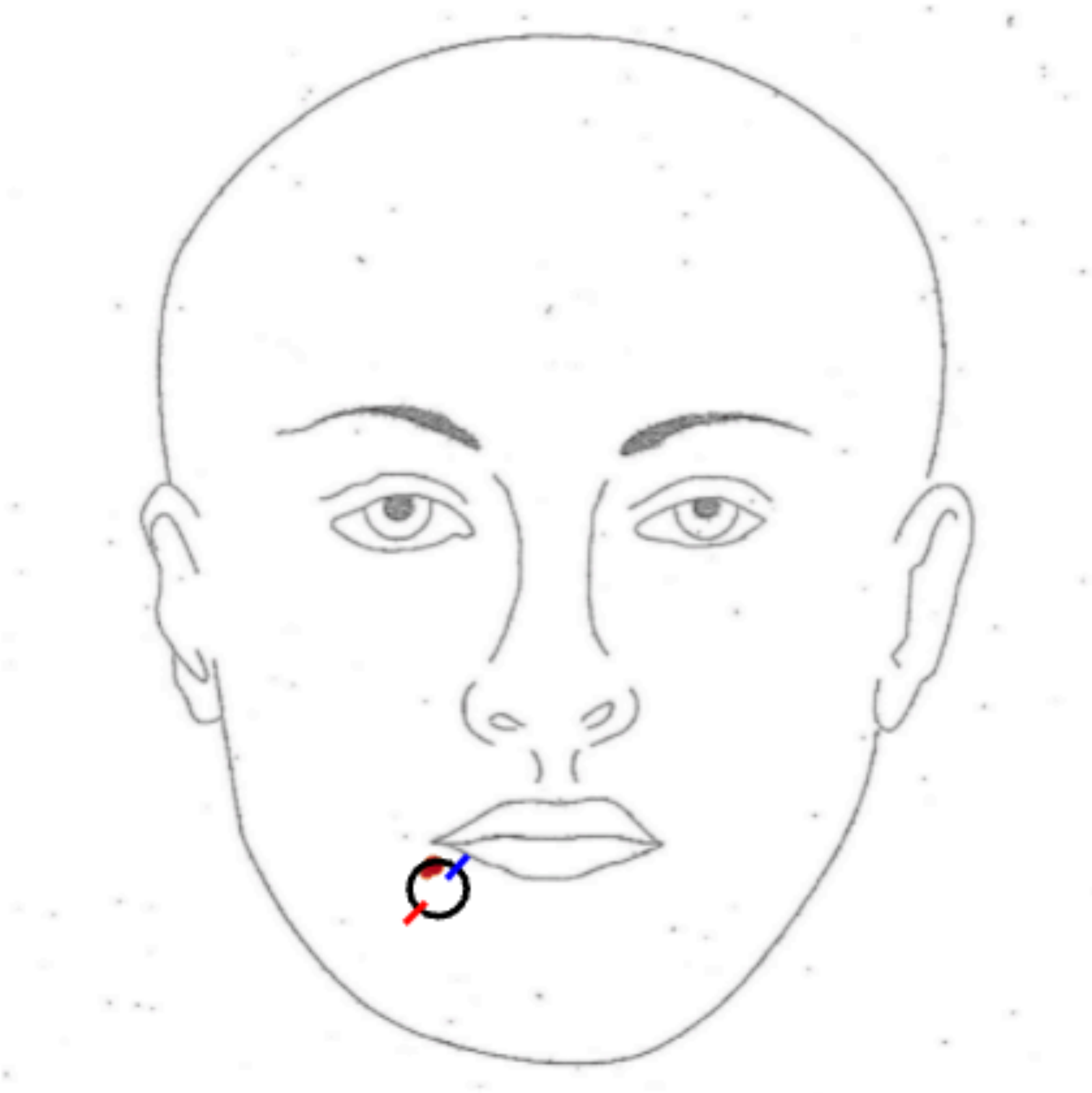

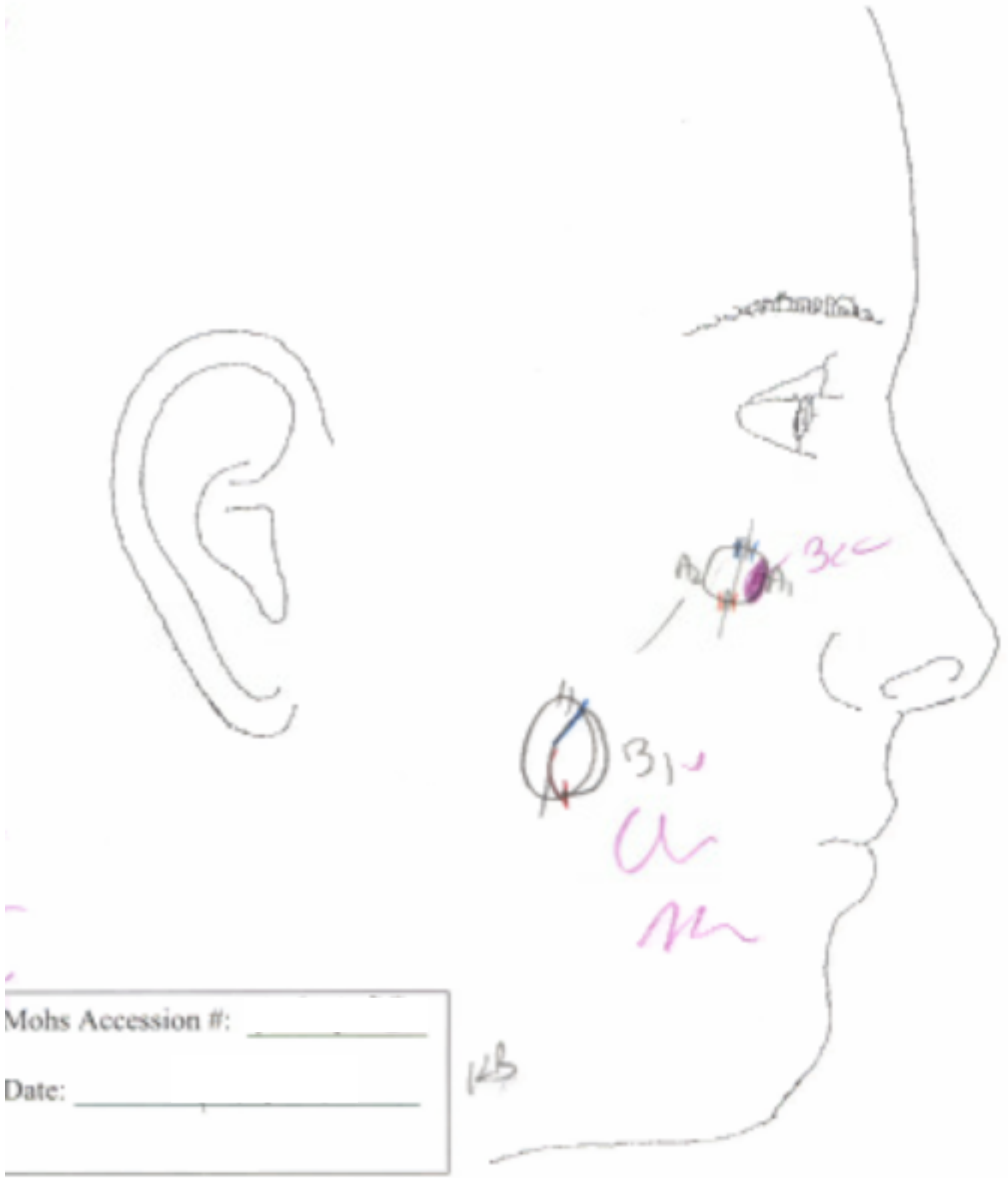
Mohs Accession #:
Date:

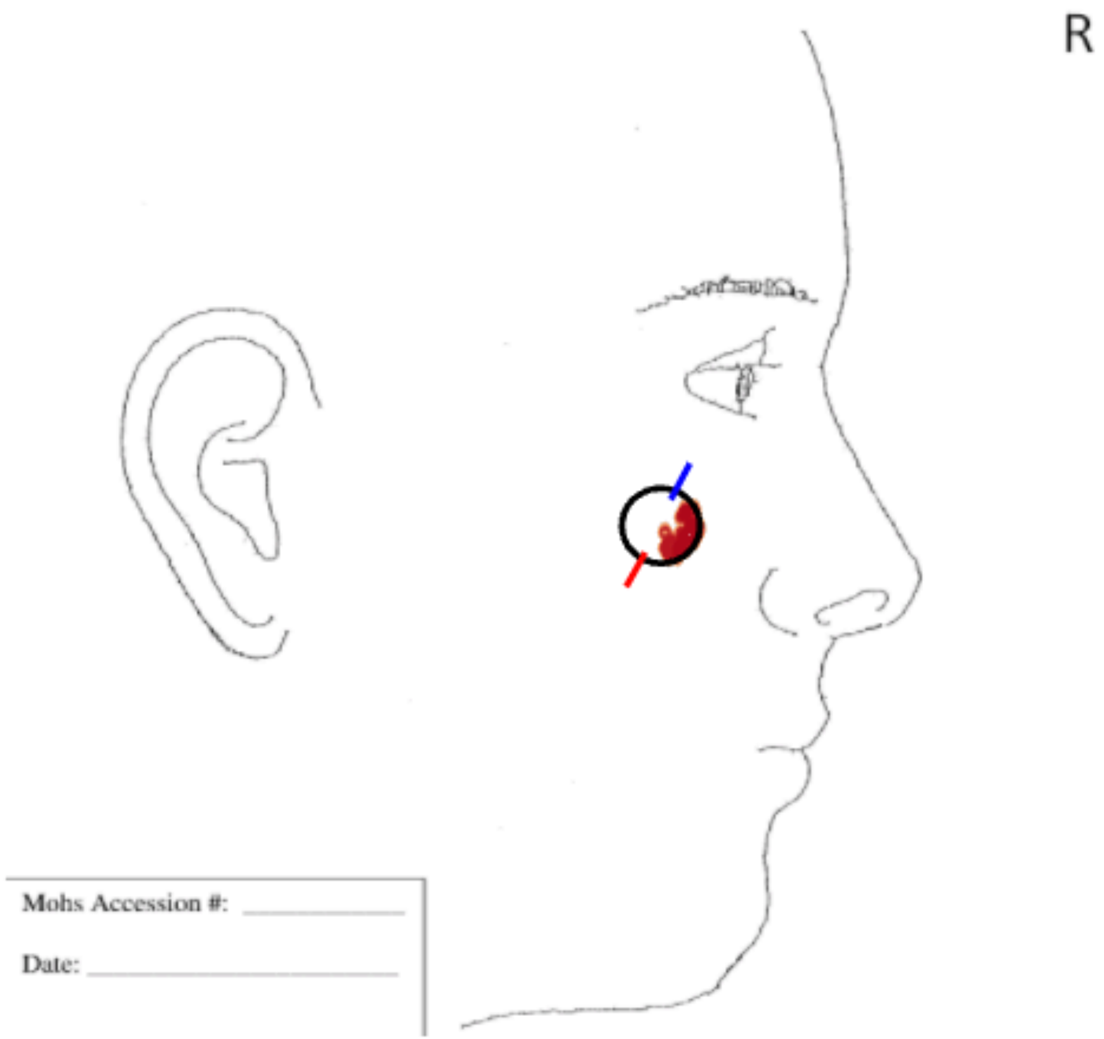
R
Mohs Accession #:
Date:

| Tumor Type | Location (per path report) | Anatomic Area |
|---|---|---|
| BCC - nodular | (L) neck | N fa[illegible] |
| Initial size (cm) | Final Defect (cm) | Room # |
| 1.1 x 0.9 cm | 1.4 x 1.3 cm | 3D[illegible] |

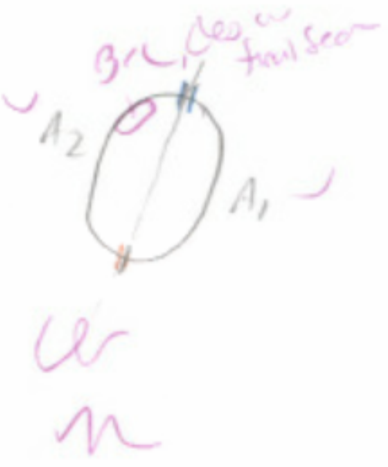


Mohs Accession #: ______

Date: [illegible]

A1

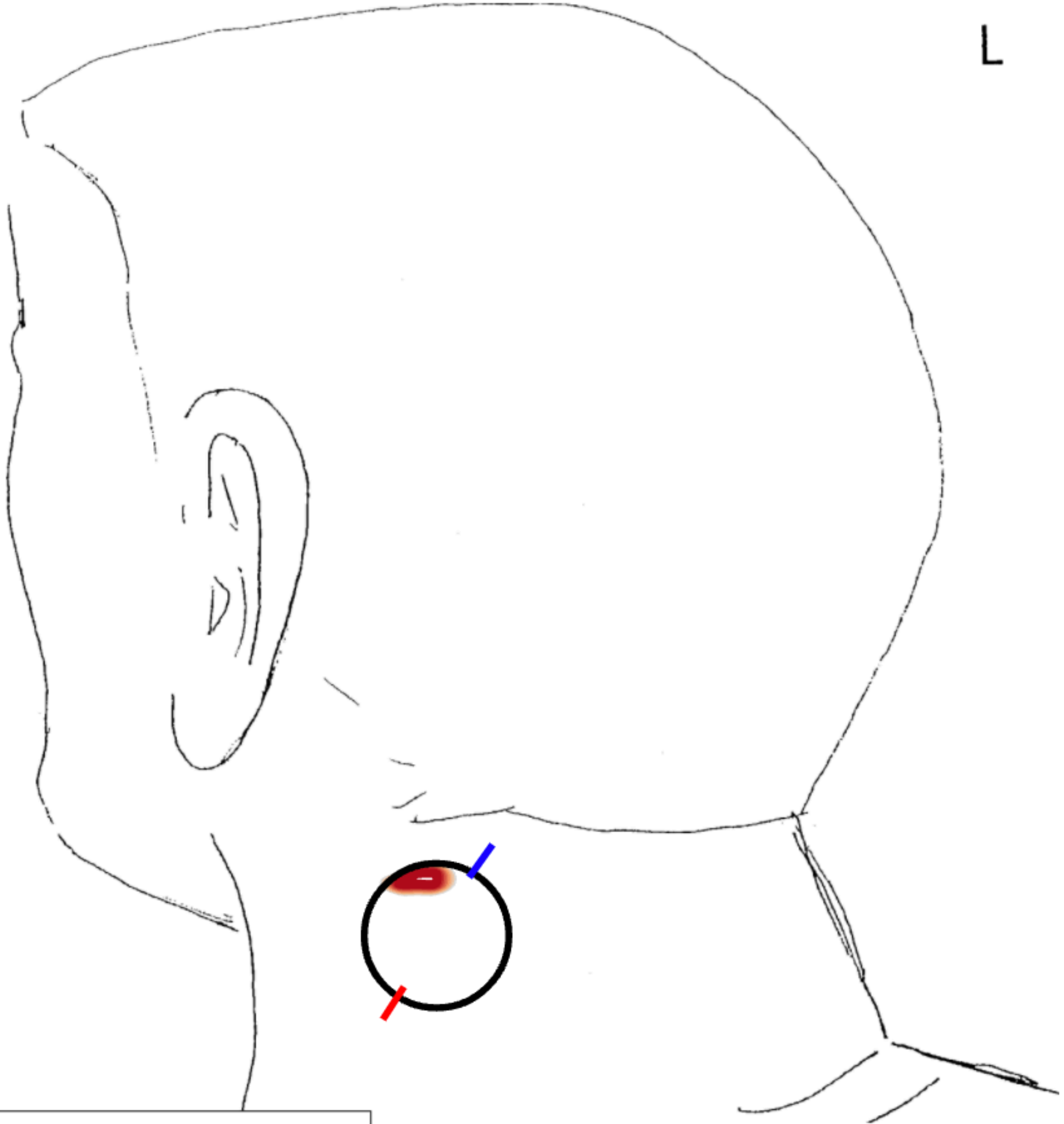


Mohs Accession #: ______

Date: ______
